# COVID-19: Data-Driven Mean-Field-Type Game Perspective

**DOI:** 10.1101/2020.07.23.20160853

**Authors:** Hamidou Tembine

## Abstract

In this article, a class of mean-field-type games with discrete-continuous state spaces is considered. We establish Bellman systems which provide sufficiency conditions for mean-field-type equilibria in state-and-mean-field-type feedback form. We then derive unnormalized master adjoint systems (MASS). The methodology is shown to be flexible enough to capture multi-class interaction in epidemic propagation in which multiple authorities are risk-aware atomic decision-makers and individuals are risk-aware non-atomic decision-makers. Based on MASS, we present a data-driven modelling and analytics for mitigating Coronavirus Disease 2019 (COVID-19). The model integrates untested cases, age-structure, decision-making, gender, pre-existing health conditions, location, testing capacity, hospital capacity, mobility map on local areas, in-city, inter-cities, and international. It shown that the data-driven model can capture most of the reported data on COVID-19 on confirmed cases, deaths, recovered, number of testing and number of active cases in 66+ countries. The model also reports non-Gaussianity and non-exponential properties in 15+ countries.

## I. Introduction

COronaVIrus Disease 2019 (COVID-19) is one of the severe acute respiratory syndrome-related coronavirus. COVID-19 is a disease caused by the virus SARS-CoV-2. SARS-CoV-2 is infecting people across several countries within few months and causing deaths, economic losses and food insecurity. As COVID-19 spreads around the globe, authorities are making decisions about local migration, confinement, isolation, quarantine and heath care system capabilities. Individuals, depending on the country, culture, social-economic context, age and season, are also making decisions concerning COVID-19. In this paper we propose an integrated mathematical model and analysis strategies for combatting the propagation of COVID-19 and for examining the economic consequences. For each infected person, one often defines the number of secondary infections that are generated by that patient. The probability distribution of that latter random variables is referred to as offspring distribution. The offspring distribution depends on its infectiousness which can be biological, environmental, behavioral or combination of these. The offspring distribution has been estimated from empirical data for outbreaks of several infectious diseases such as tuberculosis, cholera, plaque, smallpox, severe acute respiratory syndromes (SARS), MERS and Ebola. There are two important sectors that authorities are considering to mitigate COVID-19. The public health sector which tries to understand the dynamics of COVID-19 (via epidemiological models and efforts) and take measure to flatten the curve of active COVID-19 patients (mitigation and suppression). The economic policy sector has been used to alleviate negative consequences implied by COVID-19 and by the restrictive policy measures. The economic policies are also being used to prevent the amplification and the multiplication of the crisis. Unfortunately, these two sectors and approaches have not been coordinated enough. The effectiveness of the public health measures may depend on the perception of people in practice and their expectations because each individual also make its own decisions. Therefore both decisions, from economic policies by authorities and from country-specific individual preferences, should be integrated all together.

### Literature review on SARS-CoV-2

There is an important ongoing research on COVID-19. Below we review some closely related works. For SARS-CoV-2 (and hence COVID-19) the offspring distribution is not known and the dynamic COVID-19 data set which is an information that is updated on a daily basis or so, meaning it changes over time as new information becomes available.

- Effect of mobility on the propagation COVID-19: The movement and migration of people between locations add an additional difficulty in estimating the infectivity as each infected person may be moved, flown to another city, state, country or continent. Some estimations have been proposed by using contact-tracing or mobile tracking of the path of the each infected person and all its possible contacts using mobile networks and recognition. However, these estimations on COVID-19 remain limited at a global level [1]–[4]. The work [5] focuses on a Global Epidemic and Mobility model that integrates sociodemographic and population mobility data set in a spatially structured stochastic disease approach to simulate the spread of epidemics at the worldwide scale. In contrast to [5], we integrate a data-driven optimization setup for authorities and for individuals as well.
- Age and gender structure on COVID-19 death rate: Because the COVID-19 reported deaths depend on ages and gender, the role of demography on COVID-19 spread and fatality is examined in [6].
- The work in [7] explains the use of SARS-CoV-2 RT-PCR testing outcomes in Italy. The work in [8] provides some early estimates of clinical severity of COVID-19 by age.
- Epidemiological data set is needed during epidemics to best monitor and anticipate spread of infection. The work in [9] provides geo-positioned records of case of of COVID-19 outbreak. COVID-19-related deaths are not clearly defined in the international reports available so far, and differences in definitions of what is or is not a COVID-19-related death might explain some of the variations in the reported data.

Several basic compartmental models have been suggested in the literature to simplify the complexity of mathematical modelling of infectious diseases. These include

- SIR (susceptible-infected-removed),
- SEIR, that is extended SIR with exposed (E) agents
- SEIR with quarantine (Q)
- SEIR with age-structure
- SEIR with spatial location
- SEIRD that is *R* is divided into recovered (R) or deceased (D)
- SEIRD with data fitting via parameters.

These models need to be adapted to include important and key features of COVID-19 such as context-awareness, decision-making, co-morbidities per area, mobility, age, gender, healthcare status, testing capabilities to capture the evolving COVID-19 data set [10]–[14].

Table I displays some of the key differences and key features of the data-driven MFTG model.

**TABLE I:** Key features of the data-driven MFTG model.

| Feature | SIR | SEIR | SEIRD | SEIR-spatial | SEIR -age structured | MFTG |
| --- | --- | --- | --- | --- | --- | --- |
| locality | no | no | no | no | no | yes |
| city | no | no | no | no | no | yes |
| sector per city | no | no | no | no | no | yes |
| position | no | no | no | yes | no | yes |
| infection status | 3 states | 4 | 5 | 4 | 4 | 17 |
| age | no | no | no | no | yes | yes |
| gender | no | no | no | no | no | yes |
| height | no | no | no | no | no | yes |
| health | no | no | no | no | no | yes |
| family size | no | no | no | no | no | yes |
| economic status | no | no | no | no | no | yes |
| testing strategy | no | no | no | no | no | yes |
| decision-making | no | no | no | no | no | yes |
| untested positive | no | no | no | no | no | yes |
| data-driven | no | no | no | no | no | yes |

### Data-driven mean-field-type game theory

The term ‘mean-field’ has been referred to a physics concept that attempts to describe the effect of an infinite number of particles on the motion of a single particle. Researchers began to apply the concept to social sciences in the early 1960s to study how an infinite number of factors affect individual decisions. However, the key ingredient in a game-theoretic context is the influence of the distribution of states and or control actions into the payoffs of the decision-makers. There is no need to have a large population of decision-makers. A mean-field-type game (MFTG) is a game in which the (instantaneous) payoffs and/or the state dynamics coefficient functions involve not only the state and actions profiles but also the distributions of state-action process (or its marginal distributions).

In a mean-field-type game, the quantities-of-interest (such as instantaneous payoff, cost, performance functionals, state kernel/coefficients) depend not only on the type-state-actions of the decision-makers but also on the distribution of them. This allows us to consider higher order (non-linear) dependence in the distribution of state and or actions on the expected utility/cost functions. It also allows us to capture risk-awareness in the performance metric whenever one or more decision-makers are facing uncertainties. We refer the reader to [15]–[25] for more details on MFTG.

We build upon the MFTG model a data-driven methodology. The data-driven mean-field-type game theory (MFTG) model of the pandemic COVID-19 presented here is built upon Markov decision process model of epidemics. Despite the simplicity of this model, important features are extracted that can assist potential decisions on the strategy to combat the outbreak, essentially configuring a scenario ranging from short-term suppression to long-term mitigation depending on the achieved reduction in the contact number, exposed number, testing number, pre-existing health conditions, age, gender, location, mobility pattern and spatial contact rate. A coopetition and dedicated effort from individuals, governments, decision-makers and stakeholders will help in reducing the propagation but with a global high cost despite the increased rate of recovered people. Basically, data-driven MFTG is given as follows.

- Time is continuous
- Data is reported at discrete-time instants.
- Decision-makers: individuals, firms and authorities
- Control Actions: Individuals’ actions include contact/meeting rates and a movement path to go outside for food, pharmacies or other basic needs. Firms’ action include the production essential, moderate-essential and less-essential goods and total working hours. An authority at specific location controls the testing actions and migration rates within that area, between different areas of a city or between cities or the country level.
- Objective: an individual aims to reduce its risk of being infected and to limit its economic consequences. Each firm aims to maximize its revenue while keeping its employees from infections. Each authority aims to reduce the number of deaths under cost-benefit considerations (see Figure 1).

**Fig 1:**
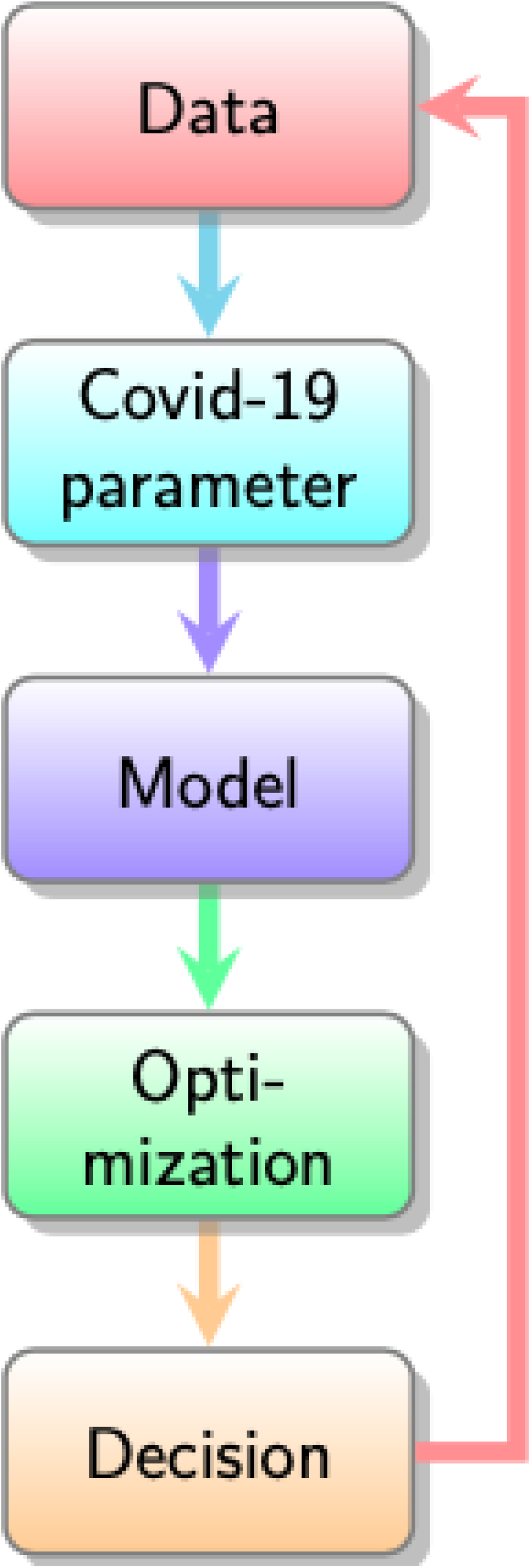
Data-Driven MFTG Model: Data, Covid-19 parameter, Model, Optimization, Decision and back to the data measurement. The verification-validation procedure used in this article.

### Contributions

The proposed data-driven MFTG model has the following key features

- context-awareness: captures some areas/city/country-specific features
- integrated decision-making and risk-aware cost-benefit analysis for authorities and individuals.
- the mobility map/pattern per areas/cities/countries is endogenous to the model
- the country-specific age-structure is endogenous to the model
- the pre-existing health conditions per city/country is endogenous to the model
- an estimation of the probability of being exposed (coughing, sneezing, surface contact)
- the hospital capacity is endogenous to the model. It is an aggregated data from number of hospital beds, intensive care facilities, ventilators, nurses, doctors, point-of-care etc.
- the testing capacity per country is endogenous to the model (the number of testing per week, number of testing facilities per areas/city/country)
- the infection status of the untested individuals is endogenous to the model

The goal of the article is to examine the data-driven MFTG modelling aspect of COVID-19. From the mean-field-type game theory perspective, one examines carefully each individual process at the microscopic level. The individual characteristics are age, gender, location, and infection status. As each infected individual may affect its evolving neighbors and path, its create an evolving mean-field term at the local, regional, national and international level. Thus, a macroscopic description of the proportion of infected per location will emerge as a global mean-field term. In addition, risk-awareness at the individual level as well as the global level is proposed. As several countries may not have the same season, culture or socio-economic context, we introduce temperature field, seasonal variation and local specificities across the countries and couple them with the decisions on the duration of the soft/medium/hard confinement. The age-structured dynamics is introduced to capture the dependence on age and on the prior medical conditions of the COVID-19 patient. For the pandemic COVID-19, many local hospitals cannot sustain a massive influx of infection and this will create a congestion and saturation. Saturation and the local capacities of the healthcare system are introduced into the model and de-congestion and curve flattening strategies are analyzed. We also examine the impact of interventions, control and risk-aware mitigation policies from the authorities. Both early control, late control and no control scenarios are examined from a network perspective using a risk-aware mean-field-type game theory. At the individual level we formulate a risk-aware decision-making problem that the probability of being infected and its possible consequences. We build upon prior optimization-based multi-agent models developed in [26]–[30]. The number of deaths in the infected population is estimated in two ways. The number of deaths at hospitals and the number of deaths who are not tested. While there is a reported data set for the number of deaths at hospitals that directly and indirectly related to COVID-19. Currently, there is no clear data set for the number of untested deaths. We use a model to estimate it.

The COVID-19 pandemic has already an enormous impact on economic, finance, agriculture, energy, and geopolitics. The decisions made by the authorities such as confinement, lockdown, isolation, curfew measures, may have severe side effects and consequences on other sectors such as employment, supply chain, agriculture and food security. Therefore a cost-benefit analytics need to be conducted for each policy per area/city/country.

The risk-aware mean-field-type game techniques presented here can be used to better understand the propagation of virus across geographical areas. As an integrated model, the impact of control strategies and switching between different locations in the propagation of the virus include

- international connectivity via flight, boat or ground transportation,
- inter-cities connectivity via flights, trains, buses, cars, motorcycles,
- within city connectivity with pedestrian mobility to pharmacies, local markets, open street air markets, supermarkets, workplaces, offices,

and isolation/confinement strategies and testing strategy based on the spreading characteristics. For finite networks, the mean-field term is stochastic and characterizes the evolution of the total number of infected/recovered/tested/untested agents per location. By introducing control strategies at both atomic and non-atomic levels, the number of deaths can be reduced. However, the control is costly and need to be carefully planned (Figure 11). Table II summarizes the heterogenous data sources.

**Fig 2:**
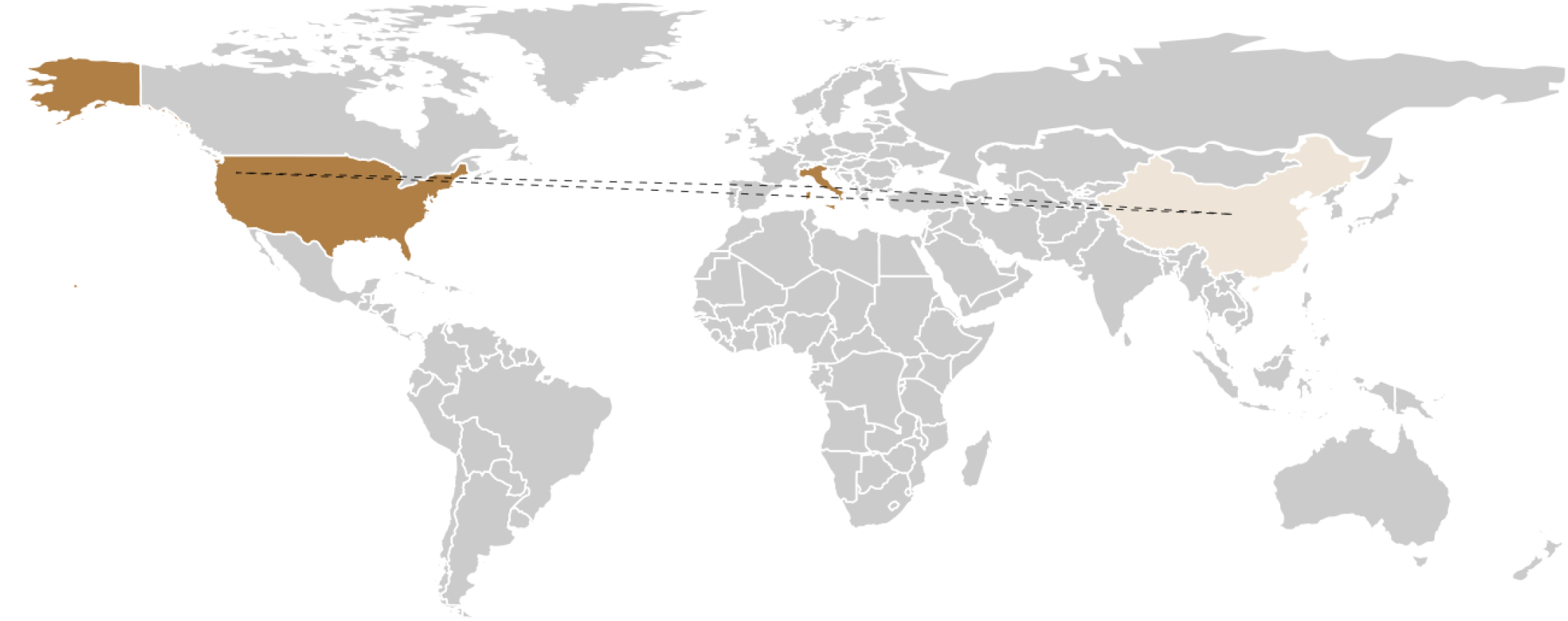
First flow of infection of COVID-19 from Asia to Europe to America and then to Africa.

**Fig 3:**
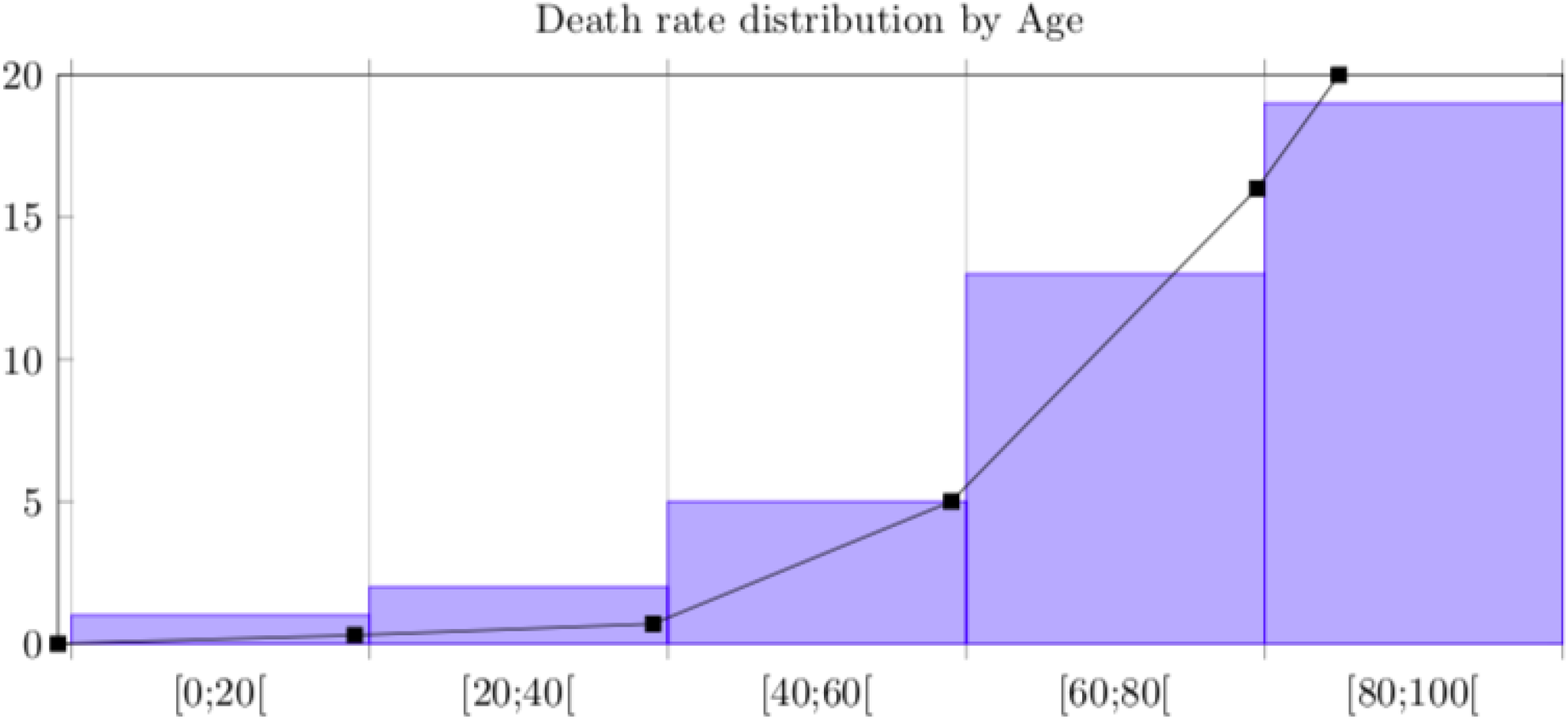
Death rate distribution by age.

**Fig 4:**
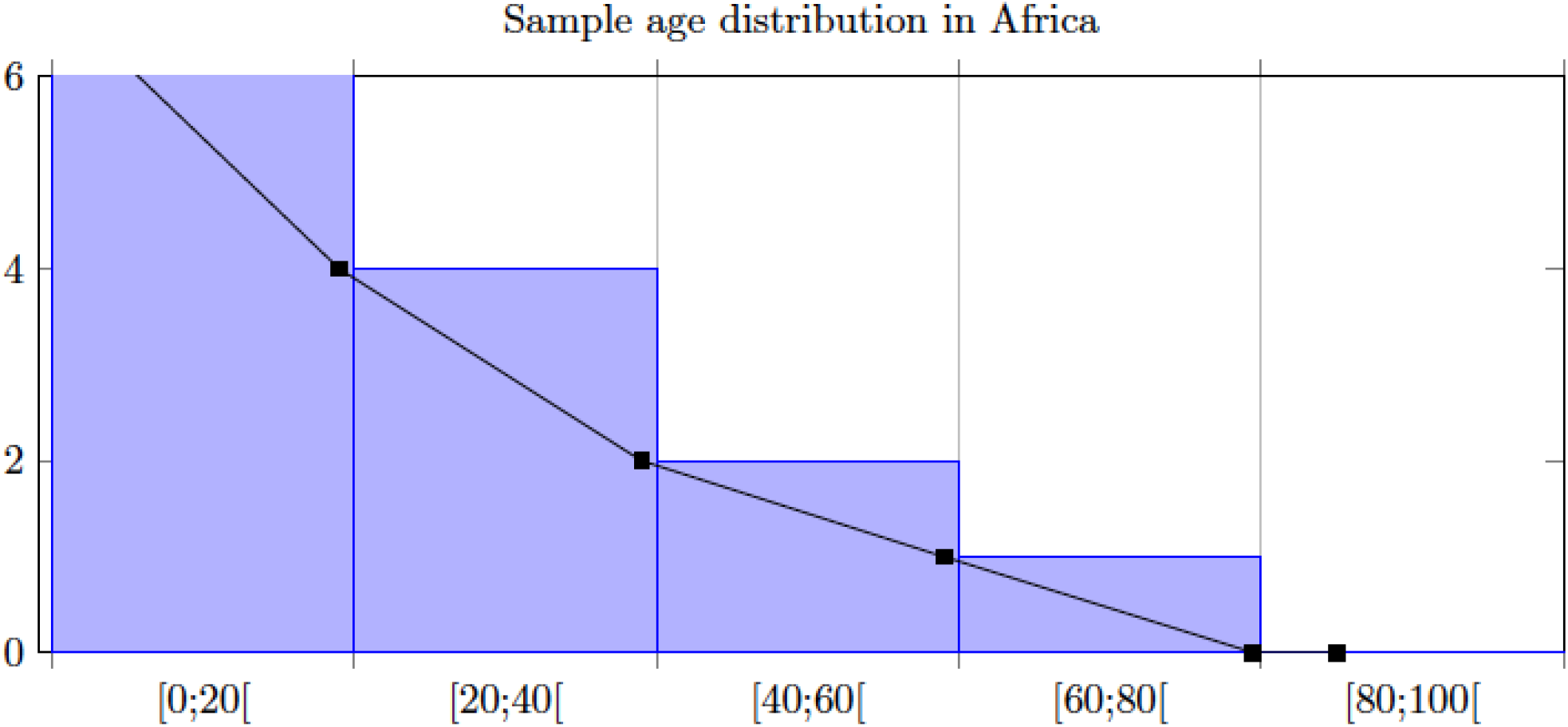
A sample age distribution (unnormalized) for a specific sector.

**Fig 5:**
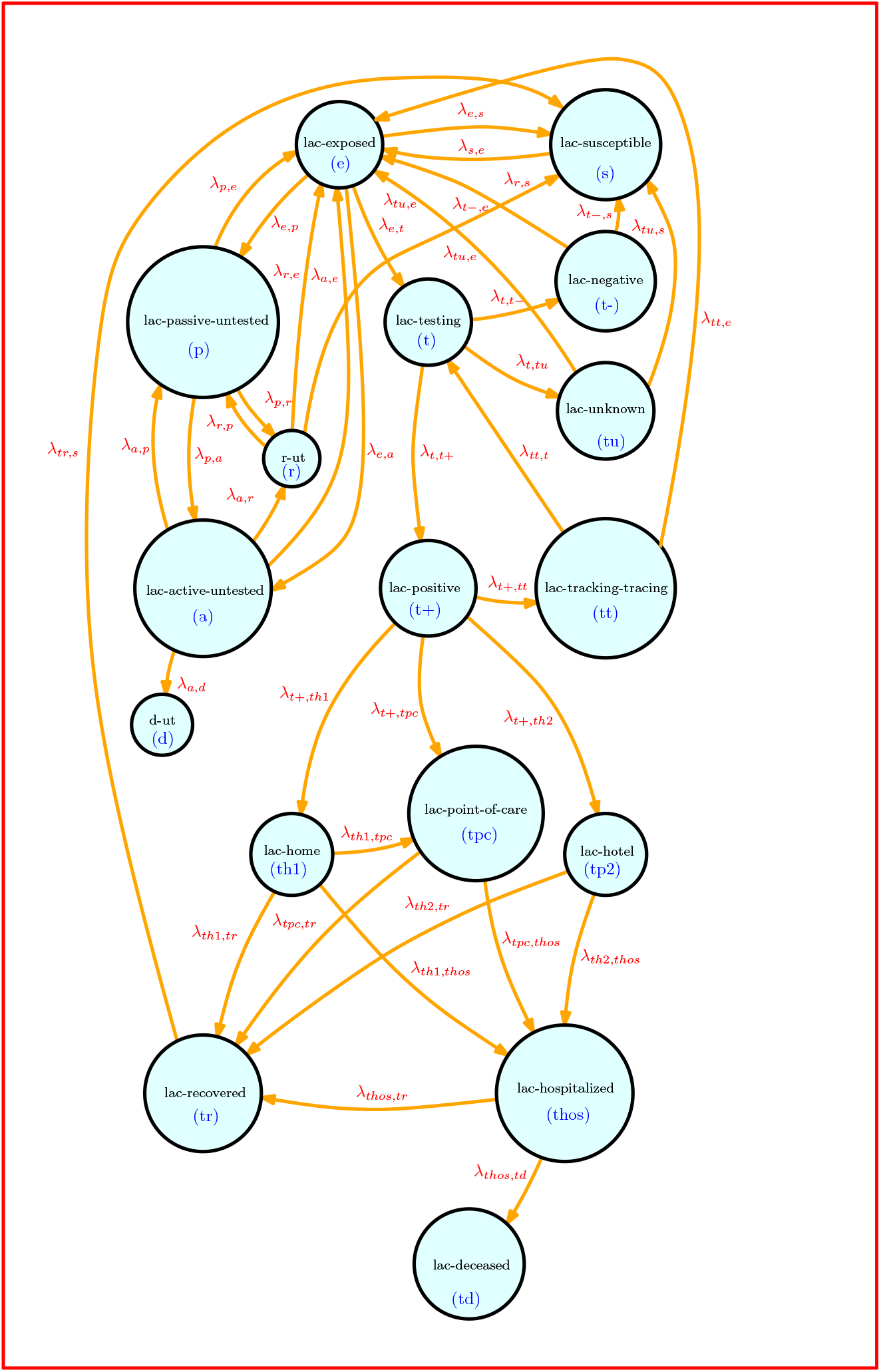
Intra-dynamics at a given location with untested and tested options. Home, hotel, point-of-care isolation. l.a.c=location(l)-age (a)-pre-existing condition (c), gender, height, family size, economic status

**Fig 6:**
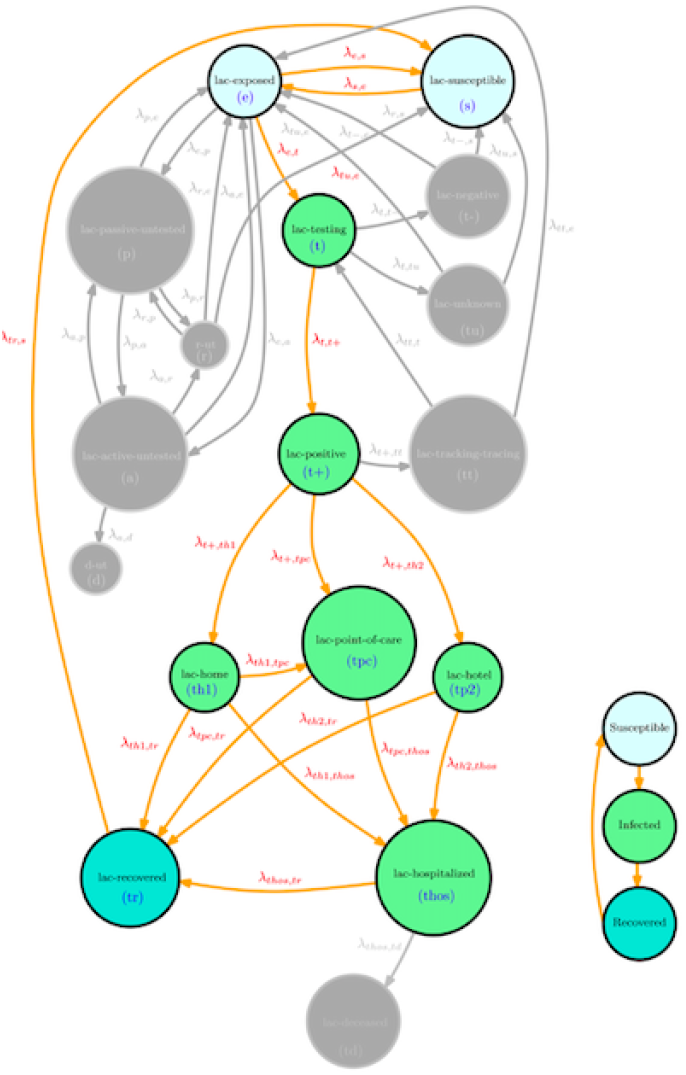
Connection of the dynamics of SIR

**Fig 7:**
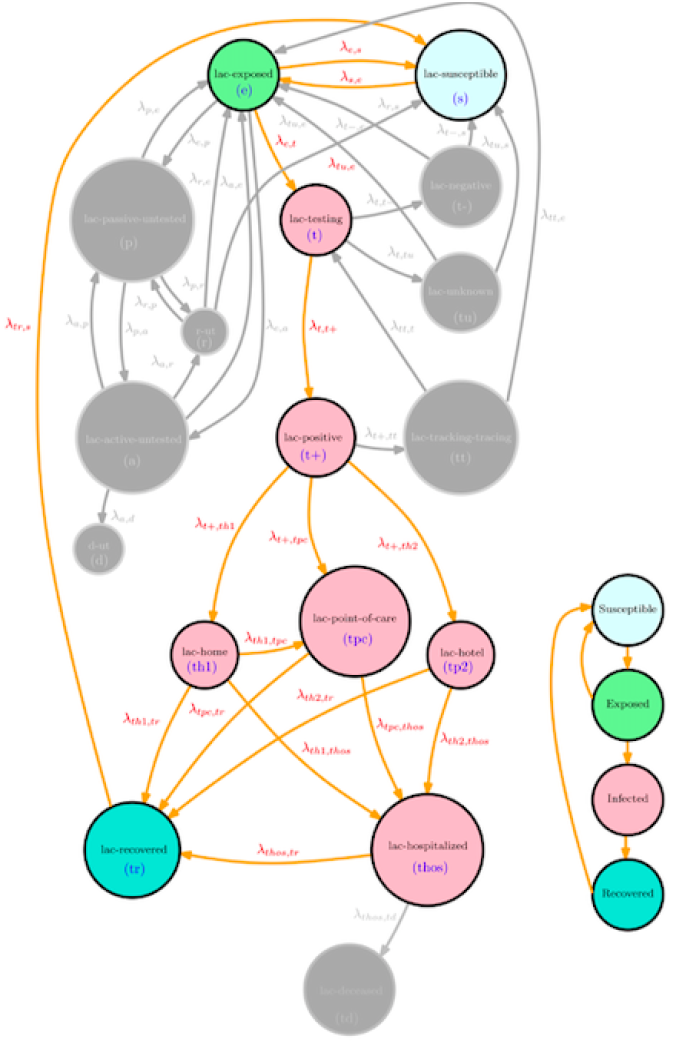
Connection of the dynamics of SEIR

**Fig 8:**
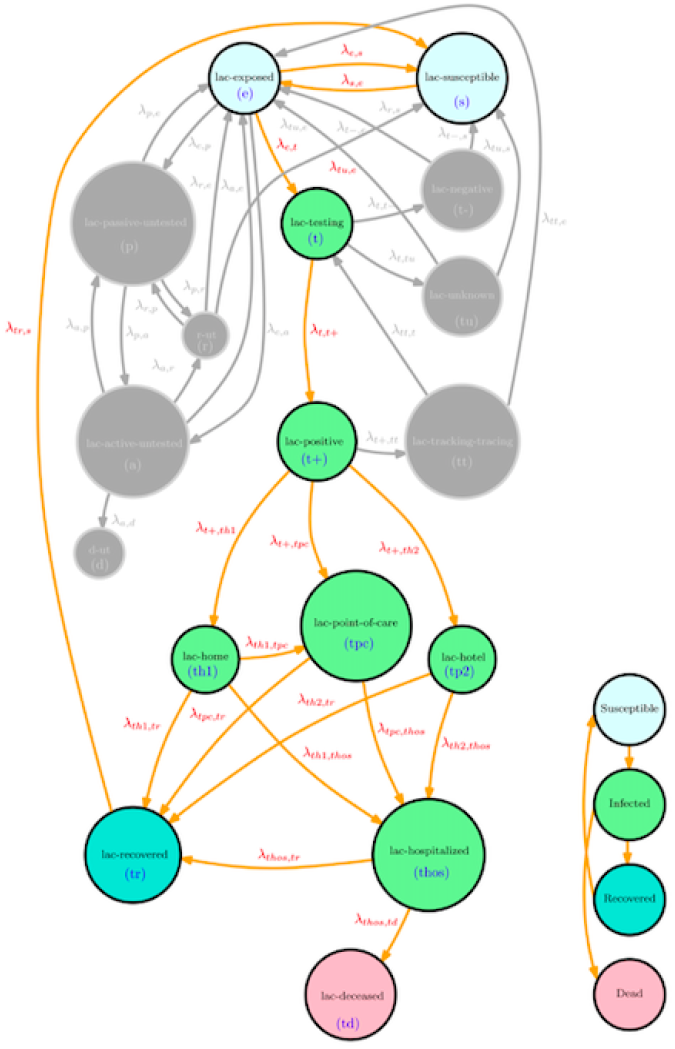
Connection of the dynamics of SIRD

**Fig 9:**
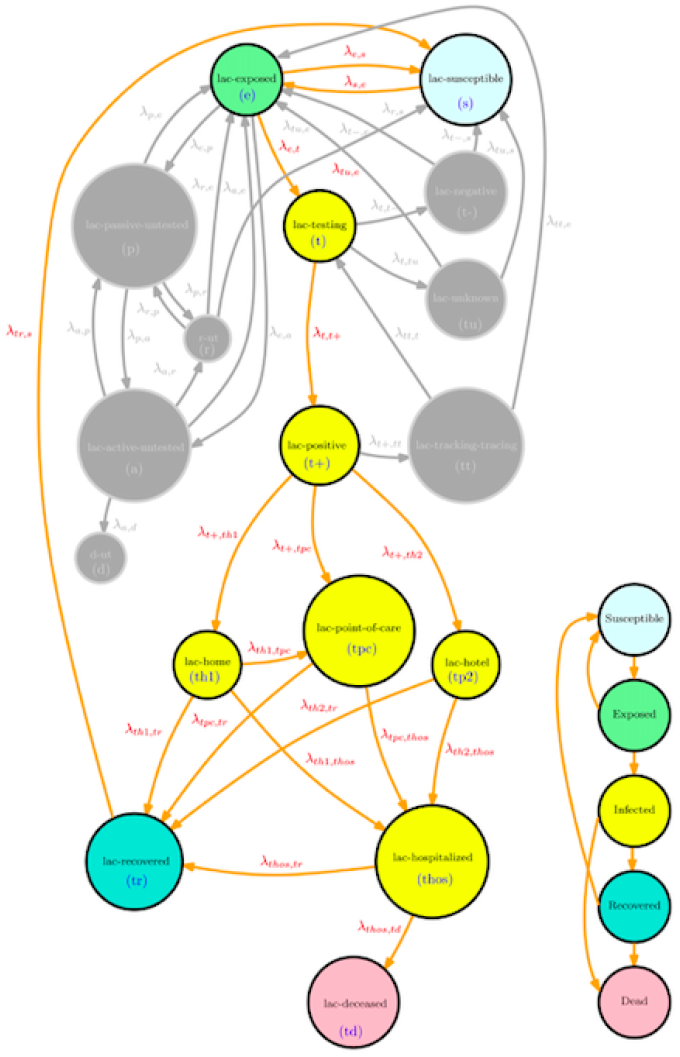
Connection of the dynamics of SEIRD

**Fig 10:**
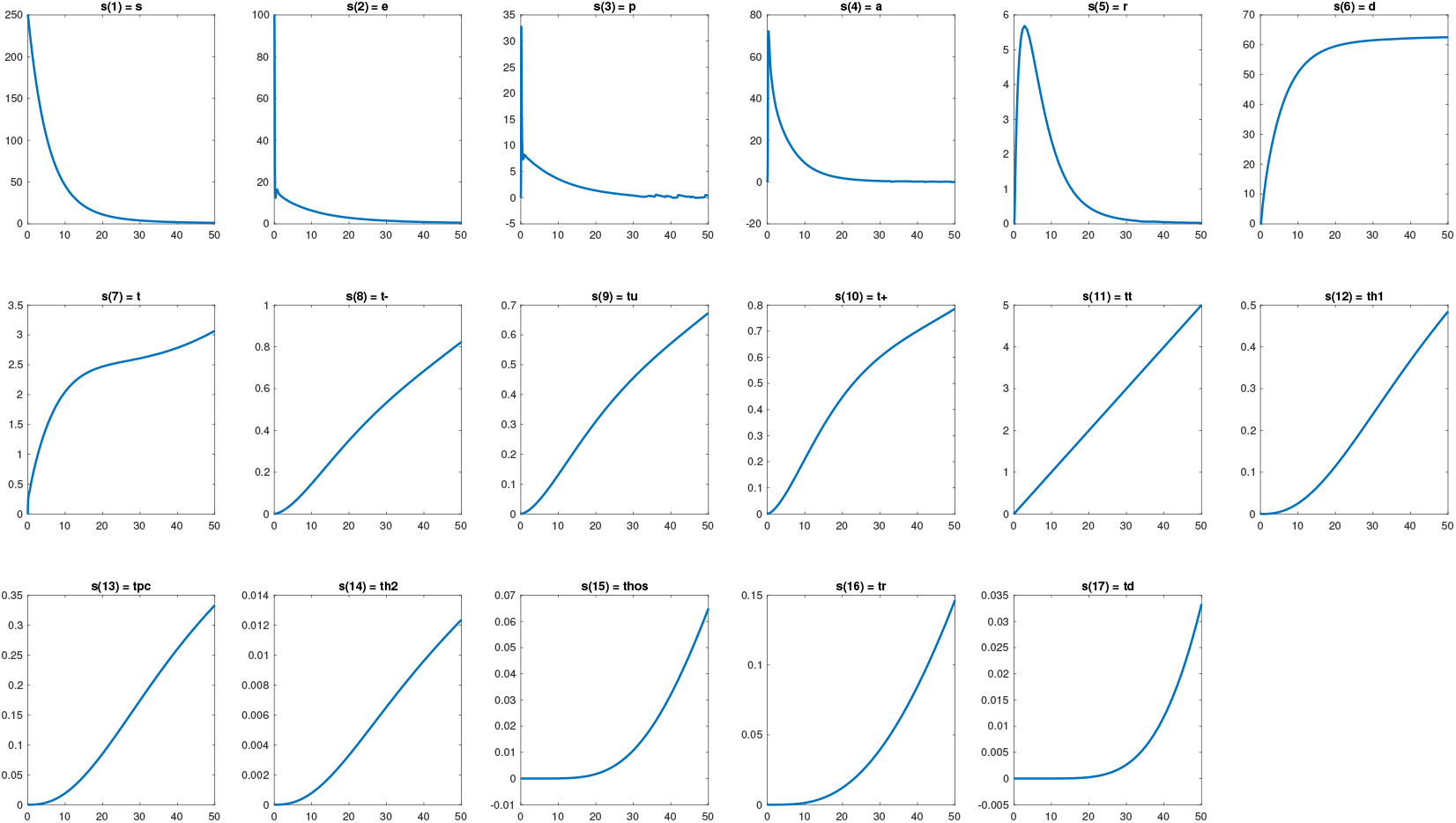
Evolution of the ODE with 17 infection states.

**Fig 11:**
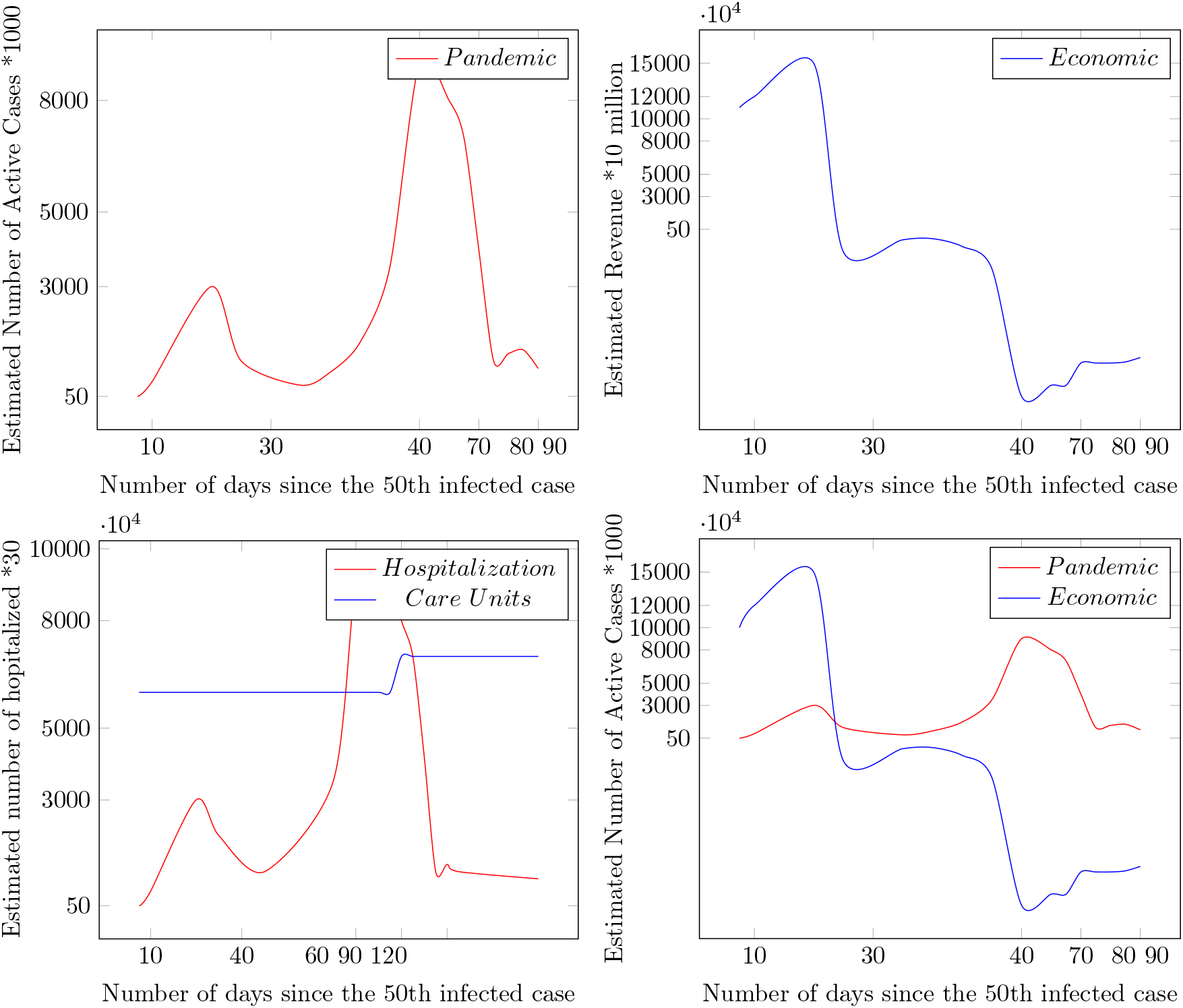
Pandemic vs Economic Consequences of COVID-19.

**TABLE II:** Heterogenous data sources.

| Data | Sources |
| --- | --- |
| International connectivity per country | Flightradar24 [31] |
| Age structure per country | [32] |
| COVID-19 data set | [10]–[13], [33] |
| Local mobility | Google Community<br>Mobility Reports [34] |
| Country-specific hospital capacity data | World Bank Group [35] |
| Geographical map | OpenStreet map |
| Economic policy | IMF |

### Structure

The rest of the article is organized as follows. In Section II we present some basic models. The data-driven MFTG setup is presented in Section III. Section IV presents the COVID-19 data set used. Section V concludes the article.

### Notation

Table III summarizes the notations used the proposed model.

**TABLE III:** Some notations used the proposed model.

| Symbols | Meaning |
| --- | --- |
| $l$ | locality |
| $l = (l_1, l_2, l_3)$ | country, city, area |
| $T(t, l)$ | temperature field |
| $\eta_1$ | migration between countries |
| $\eta_2$ | migration between cities |
| $\eta_3$ | local mobility between areas of a city |
| $z$ | age variable |
| $\rho(l, z, t)$ | initial age distribution |
| $\bar{t}_l(t)$ | testing capacity per week |
| $\delta_l$ | testing strategy at $l$ |
| $c = (c_1, c_2, c_3, c_4)$ | pre-existing health condition,<br>gender, height, family size, economic wealth |
| $x = (x_1, x_2, h)$ | position of an individual agent |
| $v$ | velocity of an individual agent |
| $s$ | individual epidemiological states |
| $n_l$ | subpopulation size at locality $l$ |
| $u$ | meeting/activity strategy |
| $m$ | mean-field |
| $f$ | Drift |
| $\beta$ | interaction kernel |
| $\epsilon$ | physical distancing of 2 m |
| $e$ | economic wealth |

## II. Basic models

SIR We start with a basic system of differential equations to estimate the propagation in an homogenous population with size *n* = *n*(*t*_0_) ≫ 1 at time *t*_0_ and three compartments (Susceptible “S”, Infectious “I”, Removed “R”) There are three different variables (S, I and R) representing the status of each compartment.

*β* represents the disease transmission rate and *γ* describes the “removed” rate.

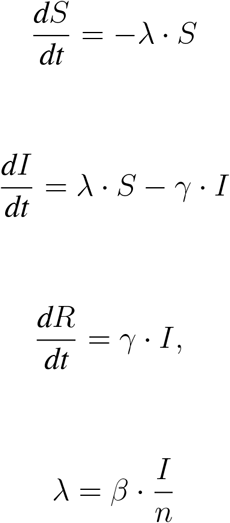

An important parameter, called basic reproduction number, is given by:

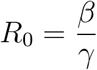

### A. Data-Driven SIR

We would like to have an idea of *I*(*t*_0_), *β, γ, R*_0_. We aim to estimate them from the dynamic data set that reported (publicly) by authorities.

- Given the data sets
  − infected cases *Î*(*t*_0_), *Î*(*t*_0_ + 1), …, *Î*(*t*_0_ + *k* − 1),
  − Removed cases 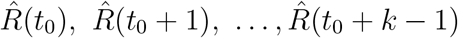 propose an estimate of 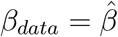 and 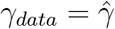
- Compute an estimate for *R*_0_ We estimate the rate parameters by solving a dynamic optimization problem.
- Given the data sets, solve the basic optimization problem

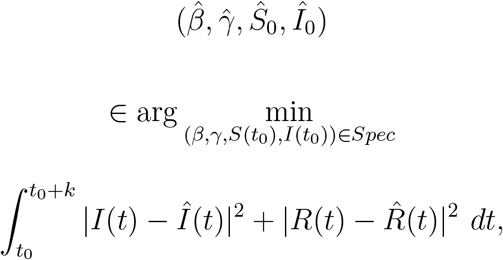
- This provides 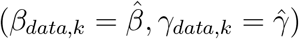

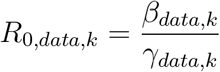

where *Spec* means the specification parameters and *k* ≥ 1.

This leads to a Data-Driven SIR:

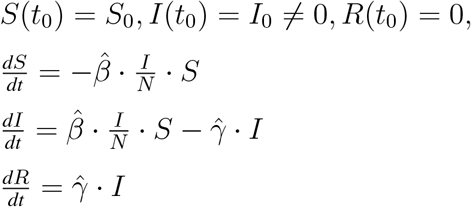

From the latter system one can obtain the trajectories of (*S, I, R*) over time. Then, several questions arise

- What if the data set changes regularly?
- Weekly estimates?
- Monthly estimates?
- Long-term estimates?

It is important to notice that for some countries, the data set provided need to be structured per week, two weeks, month time windows or a proper moving average to learn a meaningful trend of the data.

Data-Driven SIR

- Pros
  − we have a data-driven model of SIR
  − it is a simple model
  − it is compartment-based
  − it is based on the data set 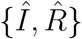 during the time window [*t*_0_, *t*_0_ + *k* − 1]
  − it is deterministic
- Contras:
  − *N* is fixed and global
  − not localized
  − it is homogenous
  − Does the shape of *I, R* capture the observed data 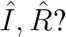

Next, we relax the restrictive assumptions made in the previous model and include context-awareness into the data-driven model.

### B. Basic SIR with location, age and economic wealth

We introduce the SIRD (susceptible-infected-recovered-deceased) dynamics with spatial location *x* over a map 𝒟_*l*_ at locality *l*, socio-economic status *e* and age-structure *z*.

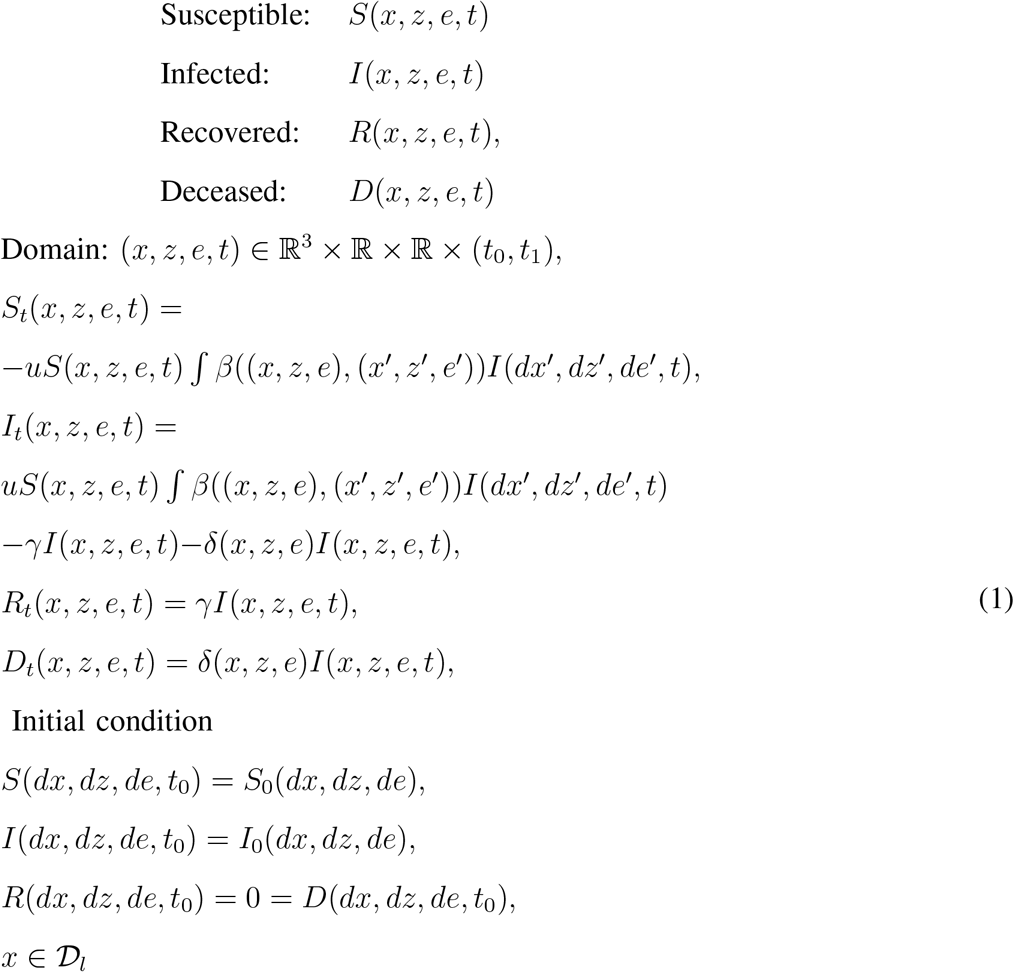

where *u* is the meeting rate, *γ* is the recovery rate, *δ* is the death rate. These are nonnegative real-valued.

From (1) we observe that the function

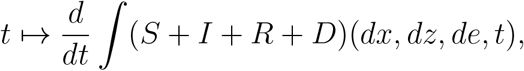

is identically zero. We set (*S* + *I* + *R* + *D*)(*dx, dz, de, t*_0_) = *n*_0_, at time *t* = *t*_0_. The condition for *I*_*t*_ to be negative yields *uS*(*βI*) < (*γ* + *δ*)*I*. This means that basic reproduction number *R*_0_ depends not only on 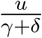 but also the kernel *β* which captures physical distancing and economic wealth interaction in the society. We choose *I*(*x, z, e, t*_0_) to be higher for very high *e* to capture the initial infection from (international) travelers and people who are in touch with international businesses. Then the contact rate kernel *β* is inversely proportional with *e*. The spread is transferred to areas with low income, the dynamics exhibits a higher infection of COVID-19 in some places with low income per family size. However the spread transfer from higher income population to a population with lower income may be slower in some countries due less interaction *β* with infectious people.

### C. Control of SIRD with spatial and age structure

We introduce the control of the SIRD (susceptible-infected-recovered-deceased) dynamics with spatial location *x*, socio-economic status *e* and age-structure *z*. Let *t*_0_ < *t*_1_ and (*x, z, e, t*) ∈ ℝ^3^ × ℝ × ℝ × (*t*_0_, *t*_1_).

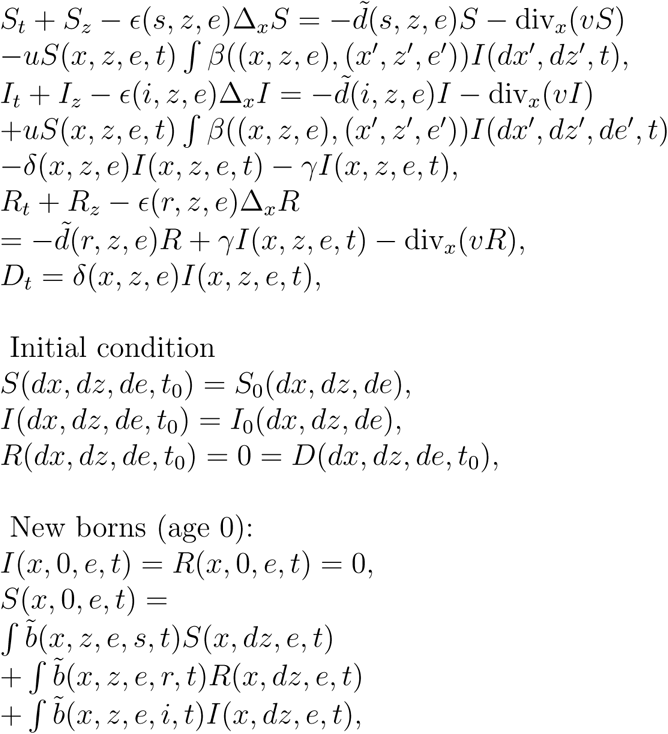

where the number of infectious people inside the ball ℬ_*ϵ*_(*x*), within two meters radius *ϵ* is given by

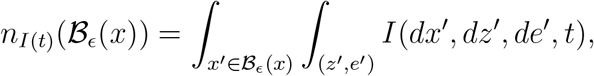

*u* is a control functional, *ϵ*(.) is a diffusion coefficient, ∆_*x*_ denotes the Laplacian operator, div_*x*_ denotes the divergence operator, *β*((*x, z*), (*x*′, *z*′)) is the interaction kernel by age and spatial position of the agents, 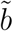 is the fertility rate of the population of Susceptible (S), Infected (I), recovered (R), 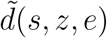 is the death rate by age and infection status, *v* is the velocity (three-dimensional), and *e* is the economic wealth. Then the optimal control of SIRD becomes a Mean-Field-Type Control Problem, and is given by

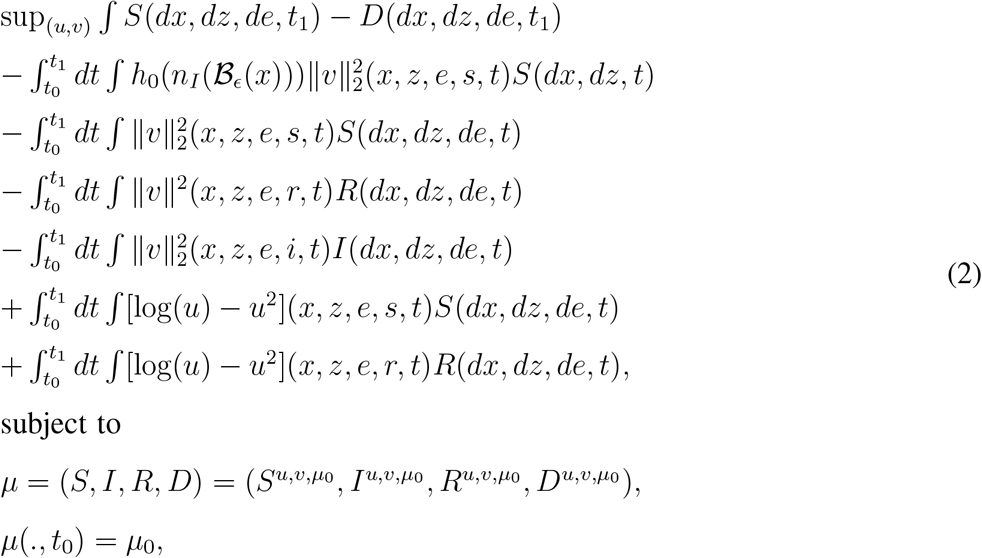

### D. A basic SIRD using Mean-Field Game

We formulate a Mean-Field Game Problem as follows

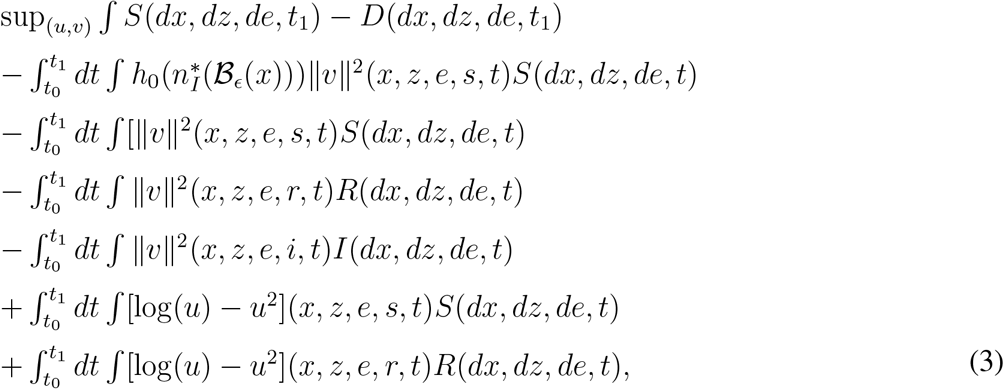

Note that Problem (3) is linear w.t.r the measure *µ* (but non-linear in *m*).

### E. A basic SIRD using Mean-Field-Type Game

We now introduce a basic Mean-Field-Type Game (MFTG) as follows

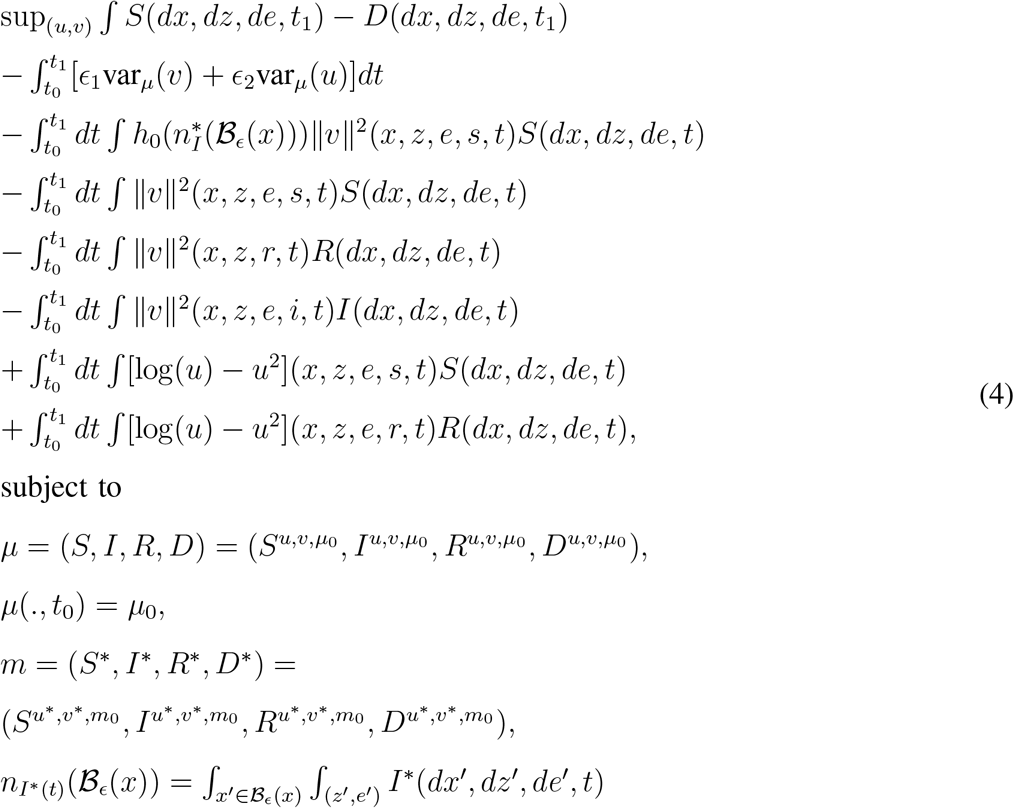

Note that Problem (4) is nonlinear w.t.r the measure *µ* (and non-linear in *m*).

## III. Data-Driven MFTG

There are *K* ≥ 1 countries distributed over a three-dimensional geographical area. A projected map is given in Figure 2. The set of countries is represented by a set of locations {1, …, *K*} with geographical coordinates (latitude, longitude) on the map.

### A. Authorities, Firms and People

- Authorities: In each country/location, there is an authority who can decide on the testing policy (when, how many), migration, confinement policies of the country and international connections via flights, boats and ground transportations. The authority in *l*, has a testing capacity, and the migration rate *η*_*ll*_*I, l* ∈ *N*_*l*_, where *N*_*l*_ is the set of connecting areas/cities/countries from *l*. The intra-city mobility in city *l*_3_ is represented by the mobility rate *η*_3_, the inter-city mobility from/to cities *l*_2_ is represented by the mobility rate *η*_2_, and the international connectivity from/to country *l*_1_ is represented by the mobility rate *η*_1_. The country specific seasons and temperature field *T* (*t, l*) are given. The temperature field will be taken into consideration when making a decision on the duration of a lockdown. In each country in {1, …, *K*} there is a subpopulation of people, called agents. The distribution of age of all agents in country *l* is given by *ρ*(*l, dz, t*) where the (chronological) age *z* ranges from 0 to 124 years. The function *ρ* is fitted from real data set making it heterogenous from one country to another. The age distribution data per country is available in [32]. An illustrative location-specific age distribution (unnormalized) function is displayed in Figure 4. Figure 3 illustrates a sample (unnormalized) death rate per age-structure. A spectrum of gender and height are also given. The authorities play the role of atomic decision-makers as their decisions can influence significantly the internal and external migration from that country, and the subpopulation at that location. The authority of country *l* aims to reduce infections, deaths and economic losses. The authorities strategic objectives are the following:
  − reduce the number of deaths
  − Interrupt agent-to-agent transmission including reducing secondary infections among close contacts and preventing further spread at a larger-scale;
  − Identify, isolate and care for patients early (testing-tracking-tracing);
  − Realizing a testing strategy at a given time window. The testing capacity 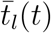 can be improved by the authority in *l* over time *t*.
  − Minimize social and economic impact through multisectoral partnerships between countries, geographical locations and airports hubs.
- Firms: Production goods are classified as essential, moderate-essential and less-essential. Each firm decides its total working hours and aims to reduce the number of infected employees.
- Agents per country: Each agent has an age *z* and a position located in one of the countries. An agent of any age can be infected by a virus (SARS-CoV-2 for the Coronavirus disease 2019). An agent with located in area *l*_3_ of city *l*_2_ of province state/country *l*_1_, age *z*,pre-existing health condition (c), gender(g), height (h), family size (fs), average revenue per family. We rename (*c, g, h, fs*) of the agent to be *c* = (*c*_1_, *c*_2_, *c*_3_, *c*_4_). and has an infection status described as follows.
  − For untested individuals, there are six states. Non-infected agents are susceptible (lac-S). Non-infected agents including recovered agents can be re-infected by another mutated version (if any) of the virus. An agent can be exposed (lac-E), for example, by an infected agent who is coughing, sneezing, from a surface contact or from a directional air conditioning (if any). Some of the exposed agents become passive (lac-P) which means dormant (asymptomatic infectious or mild symptom) and some others become active (lac-A). One absorbing state: deceased (lac-D). Some of the passive or active agents become recovered (lac-R).
  − Some of exposed people will be tested. There are ten additional states plus one specific state actioned by healthcare authorities to identify, isolate the potential exposed people for example by means of phone calls or contact-tracing and tracking. These states are Testing (lac-T), lac-testing negative, lac-testing positive, lac-testing unclassified/unknown, lac-contact tracing of the last two-weeks for each COVID-19 tested positive, lac-isolation at home, at hotel, at a point-of-care, lac-hospitalized (severely active case), lac-recovered, lac-deceased The 17 infection status are given by
  − lac-susceptible, lac-exposed
  − lac untested: lac-passive-untested, lac-active-untested, lac-recovered-untested, lac-deceased-untested,
  − lac-testing, lac-testing negative, lac-testing positive, lac-testing unclassified/unknown,
  − lac-contact-tracing-tracking: for each COVID-19 tested positive agent, a contact-tracing-tracking action is taken by the authorities to reach out and identify people who may have been potentially exposed by getting in the close contact of the COVID-19 positive agent in the last two weeks. This is not an individual but a collective action taken by the authority. A person who has been identified from the list of contract-tracing person may be considered as potentially exposed and a testing decision will be made after evaluation.
  − lac-isolation at home, at hotel, at a point-of-care
  − lac-hospitalized (severely active case), lac-recovered, lac-deceased

Figure 10 illustrates the evolution of the 17 states over time.

#### 1) Individual state

The internal state of individual *p* is (*l*_*p*_(*t*), *x*_*p*_(*t*), *s*_*p*_(*t*), *z*_*p*_(*t*), *c*_*p*_(*t*)) where

- *l*_*p*_(*t*) = (*l*_1,*p*_(*t*), *l*_2,*p*_(*t*), *l*_3,*p*_(*t*)) is an index representing latitude-longitude of the location of person *p* at time *t, l*_1,*p*_ for the country, *l*_2,*p*_ for the city/province, *l*_3,*p*_ for the area of the city. *l*_*p*_(*t*) ∈ *L* describes a discrete set of cities map of the countries in {1, …, *K*}.
- *x*_*p*_(*t*) = (*x*_1,*p*_(*t*), *x*_2,*p*_(*t*), *h*_*p*_(*t*)) represents the position of agent *p* at time *t*
- *s*_*p*_(*t*) ∈ {*S, E, P, A, R, D, s*_7_, …, *s*_17_} is the infection status of person *p* at time *t*,
- *z*_*p*_(*t*) ∈ [0, 124*y*] is the age of person *p* at time *t*, up to 124 years. The number *y* is a conversion from one year into a proper time units (miliseconds)
- *c*_*p*_(*t*) = (*c*_1,*p*_(*t*), *c*_2,*p*_(*t*), *c*_3,*p*_(*t*), *c*_4,*p*_(*t*)) with *c*_1,*p*_(*t*) ∈ *C*_1_, which is the set of all subset of 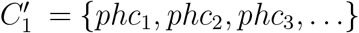 a finite list of pre-existing health conditions (phc) that are considered as relevant by health authorities. Notice that *C*_1_ contains the empty set ∅. If person *p* has no prior health issue then *c*_1,*p*_(*t*) is set the empty set *c*_1,*p*_(*t*) = ∅. The set *C*_1_ includes also some combinations of diabetes (type 2), severe asthma, pulmonary hypertension, cardiovascular disease, high blood pressure, cerebrovascular disease, chronic obstructive pulmonary disease, chronic kidney disease, among the others. Older people and people with pre-existing medical conditions are assumed to be more vulnerable to becoming severely ill with the virus.
- *c*_2,*p*_ ∈ *C*_2_ represents the gender of person *p. C*_2_ is a finite set representing the spectrum of gender, and *c*_3,*p*_(*t*) = *h*_*p*_(*t*) ∈ *C*_3_ is the height of person *p* at time *t. C*_3_ represents the spectrum of height and *C*_4_ represents the set of family size or number of roommates at residential areas.

The set of states of an agent in location *l* ∈ ℒ is 𝒳 × *C* × Status where the infection status of the agent can be {*S, E, P, A, R, D, s*_7_, …, *s*_17_} and *C* = *C*_1_ × *C*_2_ × *C*_3_ × *C*_4_.

#### 2) Global state

The population size is a large but finite number *n*. The global state of the system is the measure (*M* (*l, dx, s, dz, c, t*), *l* ∈ ℒ, *s* ∈ {*s*_1_, …, *s*_17_}, *z* ∈ [0, 124*y*], *c* ∈ 𝒞 where

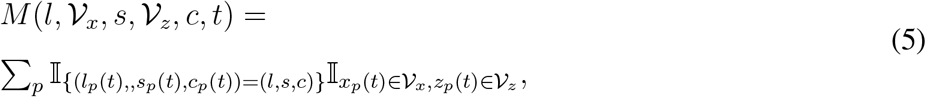

𝒱_*x*_ is a Lebesgue measurable set of positions at *l* and 𝒱_*z*_ is a measurable set of ages at *l*. The intra-state status of the system in location *l*, age *z* and characteristics (*c*_1_, *c*_2_, *c*_3_, *c*_4_) at time *t* is given by

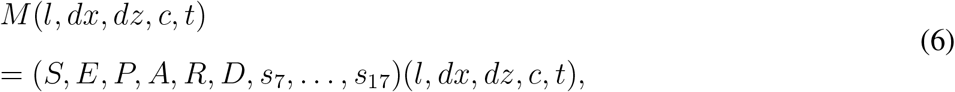

representing a measure of the total number of people located at *l* for each of the 17 status. Let *n*(*l, z, c, t*) be the population size at location *l* with age *z* and characteristics (*c*_1_, *c*_2_, *c*_3_, *c*_4_) at time *t*. Let *n*_*l*_(*t*) = ∑_*c*_ ∫ *n*(*l, dz, c, t*) be the population size at location *l* at time *t*. The occupancy measure in area *l*_3_ of city *l*_2_ of country *l*_1_ is

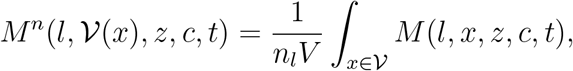

where *V* of the volume of the neighborhood 𝒱.

#### 3) Individual state transitions

We describe below the transitions between infection status.

- Susceptible agents can be exposed to the virus in different ways. (i) become exposed from a surface contact that has been infected recently. (ii) exposed from a coughing or sneezing of an infectious person (with or without symptoms) with rate *λ*_*se*_
- Exposed agents: Recent observations suggest that for the COVID-19, the risk of exposure can be affected by surface contact, coughing and sneezing (with a face mask vs without a face mask). We propose to examine the mathematics of coughing and sneezing in order to estimate the range and the risk of exposure of a person. A simple dynamics of coughing and sneezing is proposed in [36]. The trajectory of the puff is governed by the evolution of its momentum *I* and buoyancy *B*. The buoyancy force is vertical, causing the cloud to follow a curvilinear trajectory denoted by (*w, θ, h*) or equivalently (*x, y, h*)

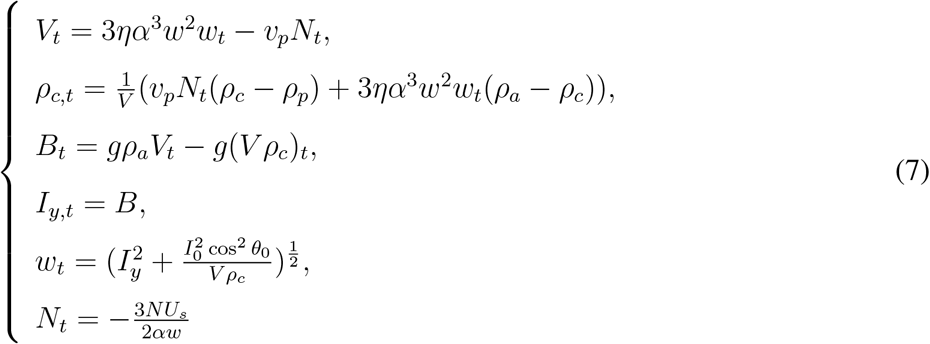

where *V* volume of the cloud, *η* = *α* = 0.1 constant, *s* distance from the source, *v*_*p*_ speed, *N* is the number of particles within the cloud, with particles concentration 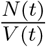 at time *t, ρ*_*c*_ (density of the cloud), *ρ*_*p*_ *ρ*_*a*_ (density of the ambient air), *B* cloud buoyancy, *g* is the acceleration due to gravity, *I* = (*I*_*x*_, *I*_*y*_), momentum, *I*_0_ initial momentum, *θ*_0_ initial angle. *U*_*w*_ = *w*_*t*_ is the speed of the suspended particles with a time scale Of 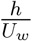 The quantities-of-interest include the fallout time, position, the horizontal range of deposition. These quantities affect the exposure to the range and possibly the transmission of the disease by emitted pathogens. We numerically compute the parameters *β, λ*_*ep,l*_ by looking at the distance over which the cough/sneeze travelled around 0.7 m, with an initial *v*_*p*_ of 4 m/s. An area of 0.17m^2^ is covered initially with an expansion rate equal to 1.3m^2^*/*s. We run the system for 45 seconds. A susceptible person present in this area will be considered as a directly exposed person with a probability proportional to the intersection of their two path areas [1].

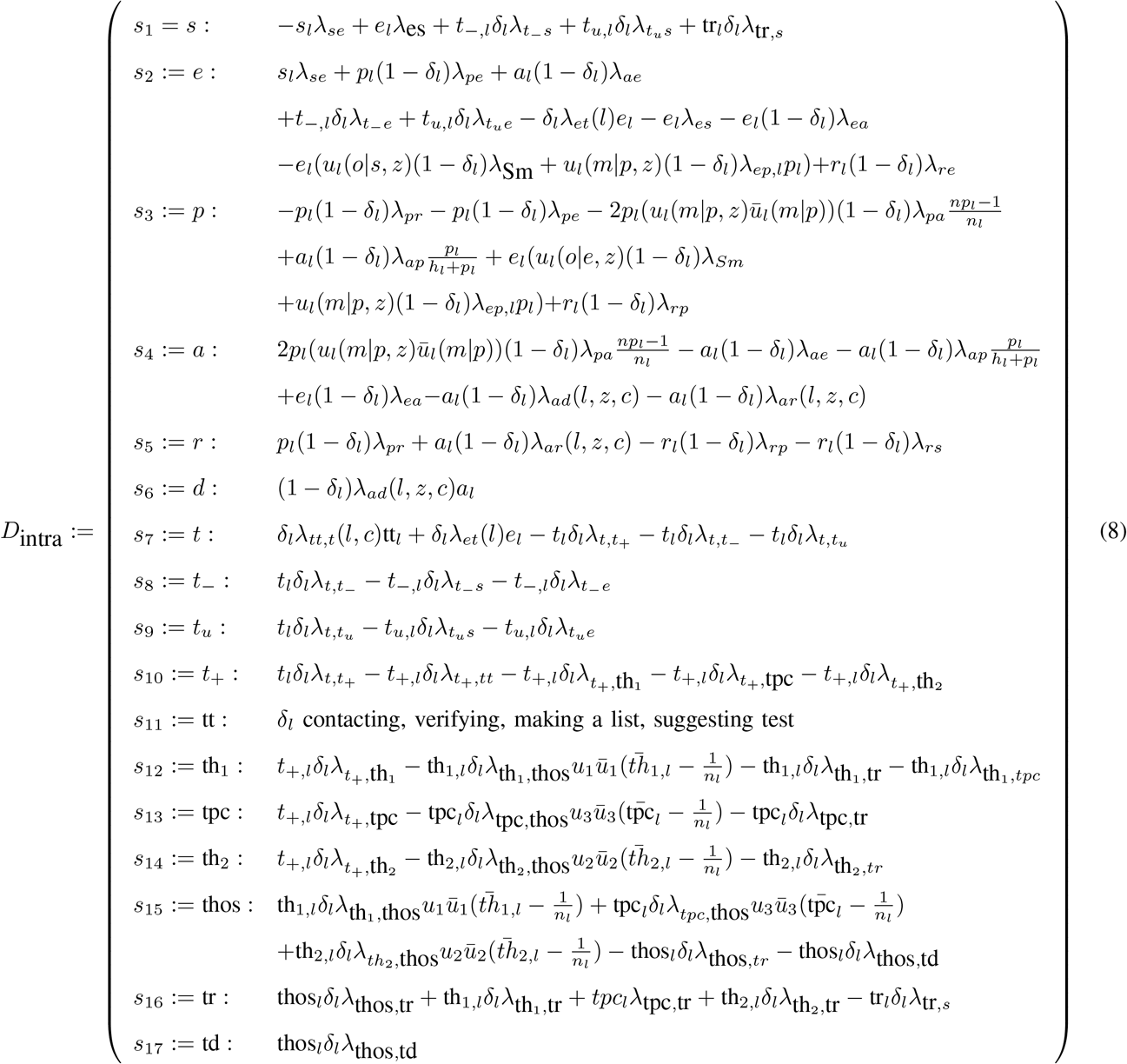 An exposed person
  − may become active with a probability *λ*_*ea*_(*l*)
  − may become passive via two ways. (i) *λ*_*Sm*_ is the probability of getting infected by a surface of contact or in close contact with an infected agent. The exposed agent can decide to receive or not an item or a visit to a workplace, grocery store, pharmacy, 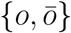. The contact probability is *u*_*l*_(*o*|*e, z, c*). In practice, the meeting or contact probability are estimated through the path trajectory of exposed people and the use of physical distancing to an exposed person or an exposed surface. (ii) 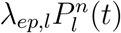 models the probability of encounter a passive agent where 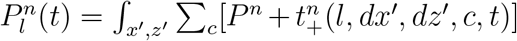. An example of interaction between an exposed agent and an active or passive infectious agent is given by

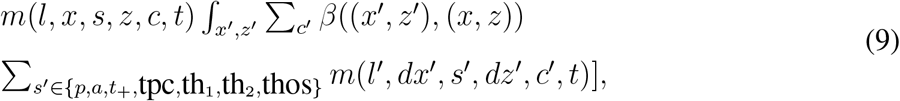

where *β* is the spatial exposure and cross-age kernel between positions *x* and *x* as in (7), and *s* ∈ {lac-S, lac-exposed}. For infection at homes and residential areas, we take the average family size per area/city/country. This is made endogenous to the model by setting *c*_4_ to the ‘family’ size (or number of roommates) of the individual.
- Untested Passive agents: In practice, the confirmed cases (asymptomatic and symptomatic) are done through a two-step process via testing for COVID-19 which can be positive, negative or underdetermined with some probabilities (*λ*_*po*_, *λ*_*ne*_). Since testing is still limited in many countries, we estimate the untested passive agents from a tracking of exposed agents obtained by phone calls, surveys, mobile tracking, contact-tracing, among the others. A large number of the population has not been tested yet. A person in country *l* is untested with probability 1 − *δ*_*l*_. where *δ*_*l*_ is the testing strategy adopted by the authority in *l*.
  − may become recovered with probability *λ*_*pr*_.
  − may become susceptible with probability *λ*_*ps*_.
  − may become active with a rate *λ*_*pa*,1_. may opportunistically encounter with another infectious agent or has comorbidities, and both become active. The rate is 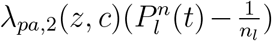. The passive agent can decide to be in physical contact or not with the other infected agent, so there are two possible actions: 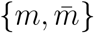. The meeting probability is *u*_*l*_(*m*|*p, z, c*).
  − may cause death with age.
  − newborns with passive status with age 0. This will be considered as a very small number as in the (publicly) reported data set.
- Untested Active agents:
  − may become susceptible with probability *λ*_*as*_(*l, z, c, ρ*) and depends on age.
  − may become passive with probability 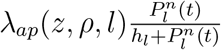. This will be considered as a very small number as in the (publicly) reported data set.
  − may cause death with age. The death rate 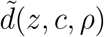, due to the virus, increases with the age *z* of the agent. The distribution of age in location *l* solves

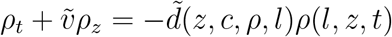

where 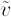 is the aging speed and 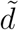 is the death rate. The initial distribution of age in location *l* is *ρ*(*l, dz*, 0). The birth rate is *ρ*(*l*, 0, *t*) = *fert*(*l, z, ρ*)*ρ*(*l, dz, t*) where fert(*l, z, ρ*) is the fertility rate at age *z* in country *l*. Older people and people with pre-existing medical conditions such as severe asthma and pulmonary hypertension, high blood pressure, diabetes, are assumed to be more vulnerable to becoming severely ill with the virus.
  − newborns with active status with age 0. The newborn part of the population contributes as *ρ*(*l*, 0, *t*).
- Untested Recovered agents: Depending on the type of virus mutation and immunization of the agent, a recovered agent can be re-infected by a mutated version of the virus and hence,
  − may become susceptible with *λ*_*rs*_(*z, c*) but it is assumed to be small as suggested by the publicly reported data set.
  − may become passive with *λ*_*rp*_(*z, c*) but it is assumed to be small as suggested by the publicly reported data set.
- Untested Death: this is an absorbing state. No transition to any other state once in. The death rate depends not only on the age of the patient but also on the temperature field or season at location *l*. It also depends the capacity of the Healthcare system at location *l*. The constraint is 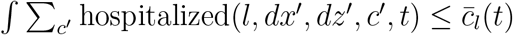, where 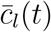 is the capacity of the healthcare system at location *l* at time *t*. When the capacity 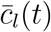 is exceeded, the death rate 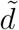 increases drastically. Authorities anticipate the saturation and find alternatives such as installations of equipments at stadiums and new hospitals. An aggregated hospital capacity data set 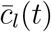 is available from World Health Organization and World Bank Group [35]. Depending on the country, there is also a certain rate *λ*_*co*_ of undetected COVID-19 patients (not reported at hospitals or from their homes) that lead to death. Figure 3 illustrates a data-driven death rate by age.
- lac-testing: During a period of asymptomatic infection, the testing can reveal infection depending on the outcome. The testing result can lead to different outcomes and the isolation can be at home, hotel or point-of-care as displayed in Figure 5. Testing occurs in country *l* with probability *δ*_*l*_.The result of the testing is often classified as positive, negative or underdetermined (unknown). Quarantine at home, hotel or point-of-care policy is case-dependent. It can be dependent on whether a case is unknown, known positive, known negative, or recovered. Testing therefore makes possible the identification and quarantine of infected individuals and release of non-infected individuals. Each country has its own testing capacity 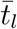 and testing rule *δ*_*l*_ (deciding by the authority at *l*) and the constraint is captured by

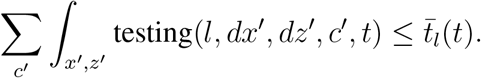
- The testing state moves to one of the three states: lac-testing negative, lac-testing positive, lac-testing unclassified/unknown.
- lac-contact tracing of the last two-weeks for each COVID-19 tested positive agent.
- lac-isolation at home, at hotel, at a point-of-care:
  − may become hospitalized with probabilities

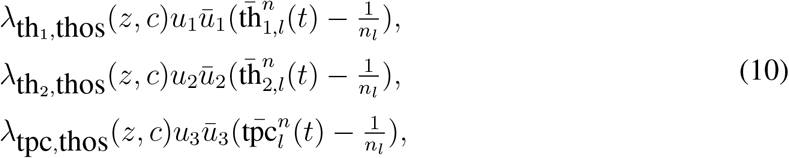

respectively.
  − may become recovered. The rates are 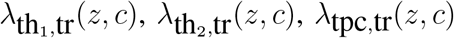,
  − There may be also a transition from home to one of the point-of-care with probability 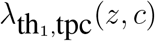.
- lac-hospitalized (severely active case),
  − may become recovered with rate 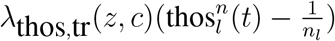.
  − may become dead with rate 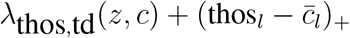.
- lac-recovered
  − may become susceptible with rate *λ*_tr,*s*_(*z, c*) but it is assumed to be very small.
  − may become exposed with rate *λ*_tr,*e*_(*z, c*) but it is assumed to be very small.
- lac-deceased: this is an absorbing state. There is no transition to any other state once in.

When *δ*_*l*_ = 0, the model (without testing at locality *l*) reduces to six basic individual states. Figure 5 represents some of the intra-dynamics for both tested and untested individual at a given location wit pre-existing health condition *c*, gender, height, family size, age *z*, and untested options. Connection with SIR, SEIR, SIRD, SEIRD can be found in Figures 6, 7, 8 and 9.

#### 4) Mobility per area/city/country

- The mobility pattern of the agents through ground transportation within the same street/area of the city, between areas of the cities, between cities via ground and air transportation and international connectivity are given by a mobility map or graph with rate functions *η*_*ll*_*I* (*T, C, c, λ, m, ρ, u, v, δ, n*) between locations *l* and *l*′. We decompose *η* into three layers: *η*_1_ connecting countries, *η*_2_ for connecting cities, and *η*_3_ for connecting areas of the same city.

The mobility pattern by area per city can be formulated as follows. Let *l* = (*l*_1_, *l*_2_, *l*_3_) be a location of a specific area *l*_3_ of the city *l*_2_ of region/country *l*_1_ and *x* = (*x*_1_, *x*_2_, *h*) ∈ ℝ^3^ a geographical location of an individual agent inside the area *l*_3_. Let *m*(*l, x, s, z, c, t*) be a measure of the agents satisfying the Kolmogorov equation (18 in Table **??**). The limiting measure *m* can be written as

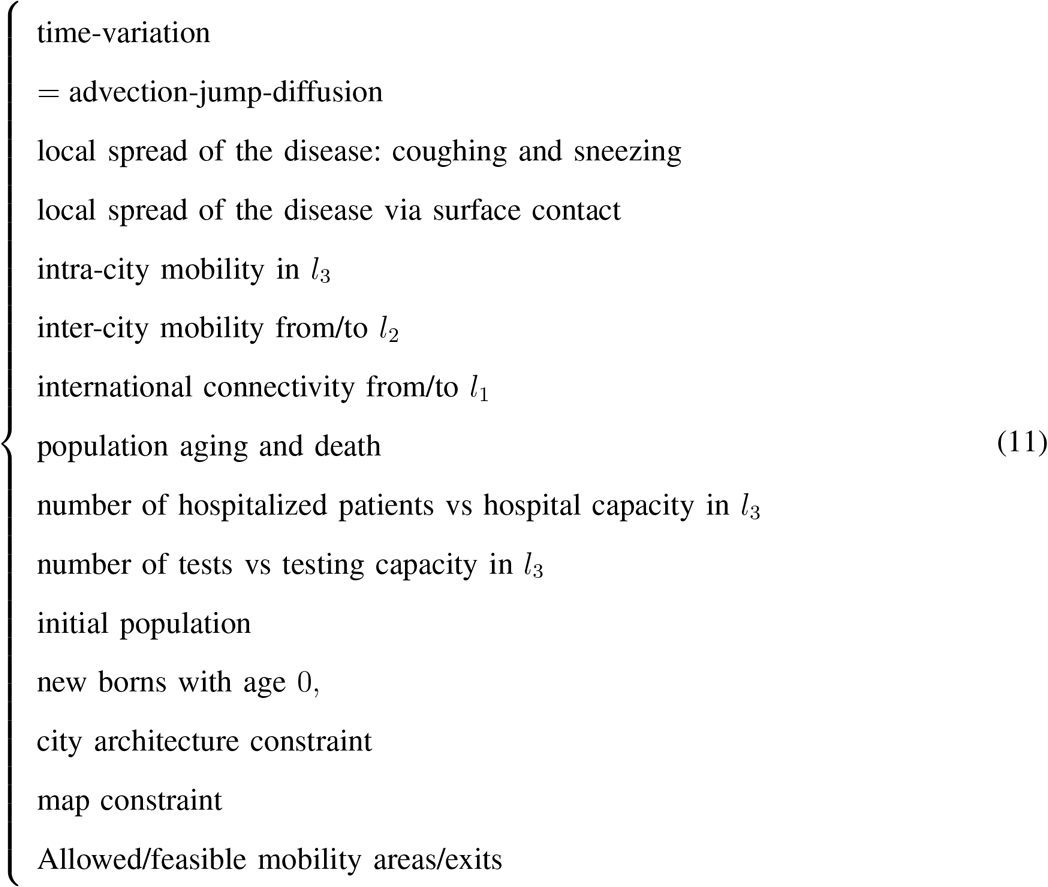

The evolution of fraction of agents per state is given by

- The number of agents for which there is a transition in one time slot is always less than 2.
- Payoff contribution from switching : 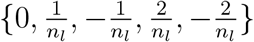.
- The intensity of interaction is in the order of 1*/n*_*l*_.
- The drift (the expected change of *M*^*n*^ in one time-step given the current state) of the system is: 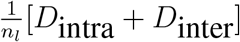, with *D*intra given in (8) and

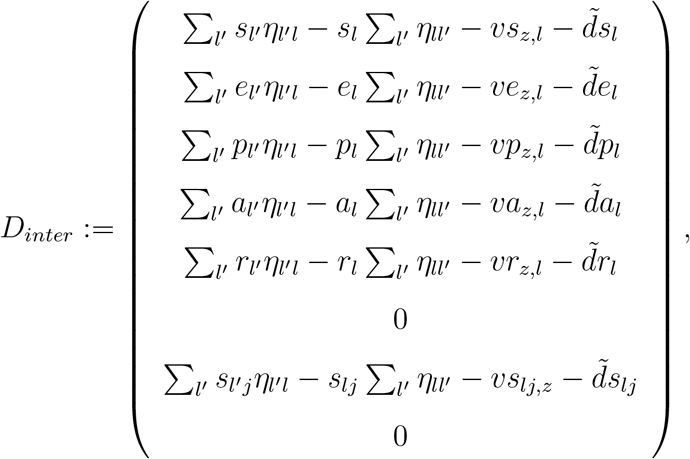

where *j* ∈ {7, …, 16}, and *m* is the actual population mean-field profile as a function of (*l, x, s, z, c, t*) given in (18) where *yq* denotes a quarantined agent with status *y*. Agents with non-empty pre-existing conditions *c* ≠ ∅ are handled separately from the above dynamics with a certain quarantine rate at the early stage. Here 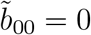 for *z* > 0 and The new borns are counted in the calculations of *m* from the functional 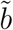with age *z* = 0. The transition rate 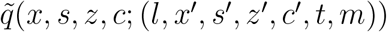 is obtained from the functions *λ* following the diagram in (8).

##### Proposition 1.

*As n*_*l*_ → +∞ *the occupancy measure M*^*n*^(*t*) *converges in probability to the solution of the* (*controlled*) *differential equation given by*

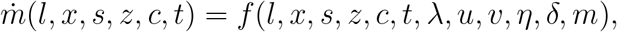

*Starting from m*(., *t*_0_) = *m*_0_(.).

The convergence is in distribution. It is, in general, weaker than the convergence in probability. However, here, the limiting object *m* is a deterministic (parametrized) object, and hence, the convergence is also in probability. The proof follows from [27], [28] by using a martingale approach with an infinitesimal generator driven by the drift

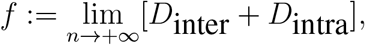

where the limit is taken in (8).

#### 5) Data-driven mean-field trajectories

We build upon the works on mean-field-type filters developed in [37]–[39] to incorporate the economic data and the pandemic data together in the model. We tune the parameters of the model such that the solution of the dynamical system becomes as close as possible to the dynamically changing data set. We rewrite the model with the parameters *λ* to be tuned from the reported data set using the mean-field dynamics in Proposition 1.

- The geographical coordinates are taken from the world map data set.
- The sequences of reported data set *ŷ*_*l*_(*t*_*k*_), *k* ∈ {0, 1, …,}, *t*_*k*_ ≤ *t* is given. The reported data set is observed at discrete time instants *t*_0_ < *t*_1_ < *t*_2_ < …. *t*_*k*_ captures the time instant at which the *k*-th data point is officially reported by the corresponding state and healthcare authorities at location *l*. The reported data set is a sequence of aggregated data set: *ŷ*_*l*_(*s, t*_*k*_) where *s* ∈ {tr,td,testing, *t*_+_}. The conditions of untested agents are not in the reported data set. The reported data set is assumed to be noisy and inaccurate.
- The reported data set is used to construct a filtered trajectory minimizing the global error within [*t*_0_, *t*_1_] by choosing the tensor *λ* and *h*_*l*_. Then the resulting dynamics is used to forecast the mean-field within an uncertainty region. The sequence of optimization problems up to time *t*_*k*_, is given by

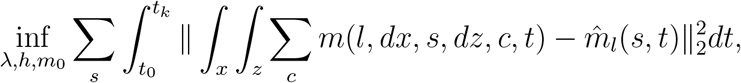

Where 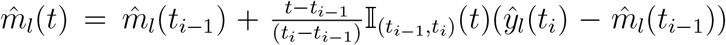. As a new data point 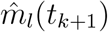 for *t* ∈ (*t*_*i*−1_, *t*_*i*_) is observed, we need to re-update the estimation over the integral [0, *t*_*k*+1_] by solving the new minimization problem 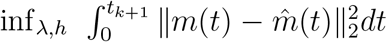, and so on. The continuation of the mean-field starts from *m*(*t*_*k*_) at time *t*_*k*_.

The above optimization provides *λ*data, *h*data that are specific to COVID-19 country-specific spread. Moreover, *λ*_*data*_ and *h*_*data*_ are context-aware as they depend on country-specific, age, gender, testing and pre-existing health conditions. The data-driven dynamics yields

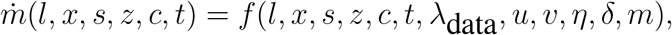

with *m*(., *t*_0_) = *m*_0_(.). Instead of 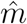 above, the estimation is replaced by the distribution of 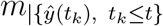 leading to a mean-field-type filtering problem [37]–[40]. The diagram of Figure 1 represents the main steps of the algorithm of the data-driven MFTG model.

#### 6) Beyond pairwise interaction

We have detailed above the pairwise interaction case with a kernel *β* in (9). A 3-wise, 4-wise,… *j*-wise interaction case, depending on the local markets, supermarkets, pharmacies, family members at home, and the number of infected people at the neighborhood *V*(*x*) can be formulated as

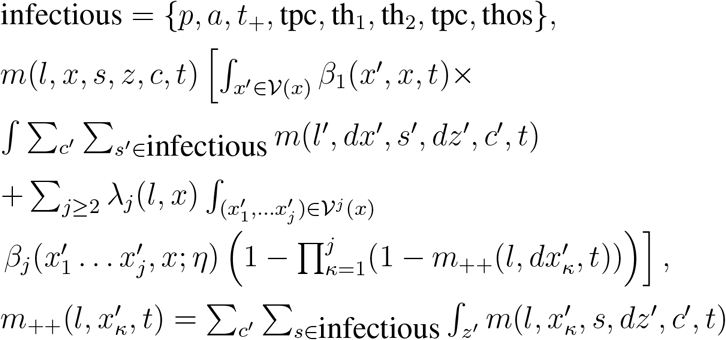

where *λ*_*j*_(*l, x*) denotes the probability of having *j* agents around *x*.

### B. Risk-aware MFTG

Introduce 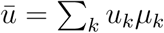 and 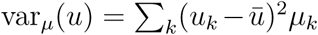. The benefit-cost functional is a risk-aware function as it depends explicitly on the variance. Each individual agent chooses *u* and solves the following best response problem given (*m, η*)

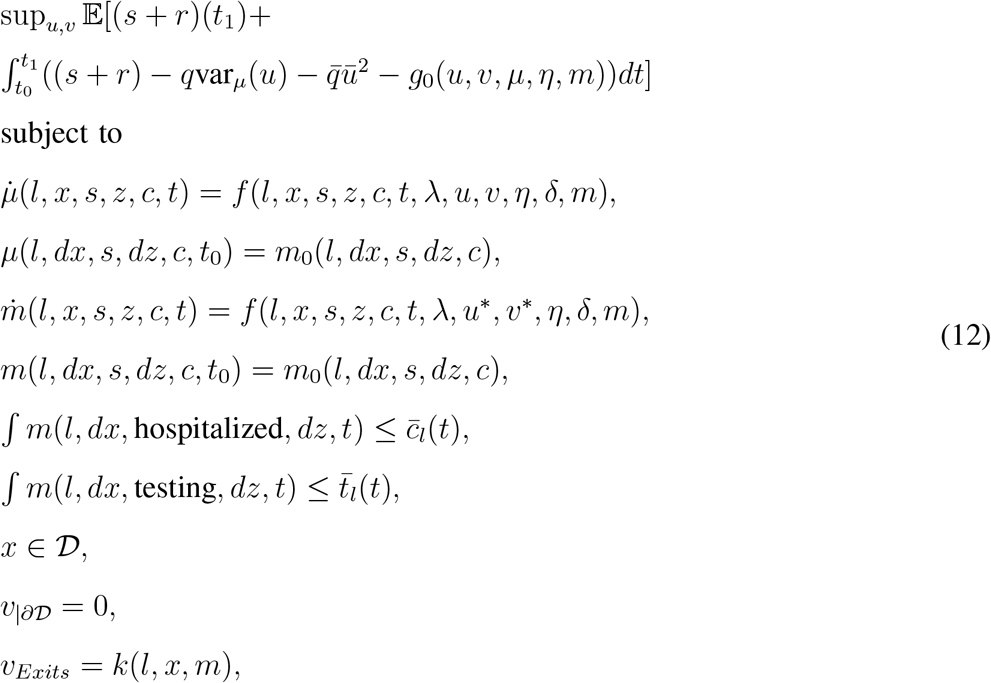

with *∂*𝒟 being the border of the map 𝒟,

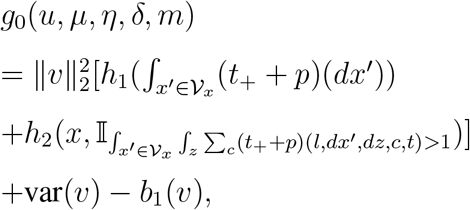

and *b*_1_ is the benefit of the agent, *h*_1_, *h* _2_ are positive and non-decreasing functions (in the second variable for *h*_2_.)

The *l*-th authority chooses a finite dimensional vector *η* and solves the following best-response problem given *m* and given the migration restrictions imposed by the other authorities.

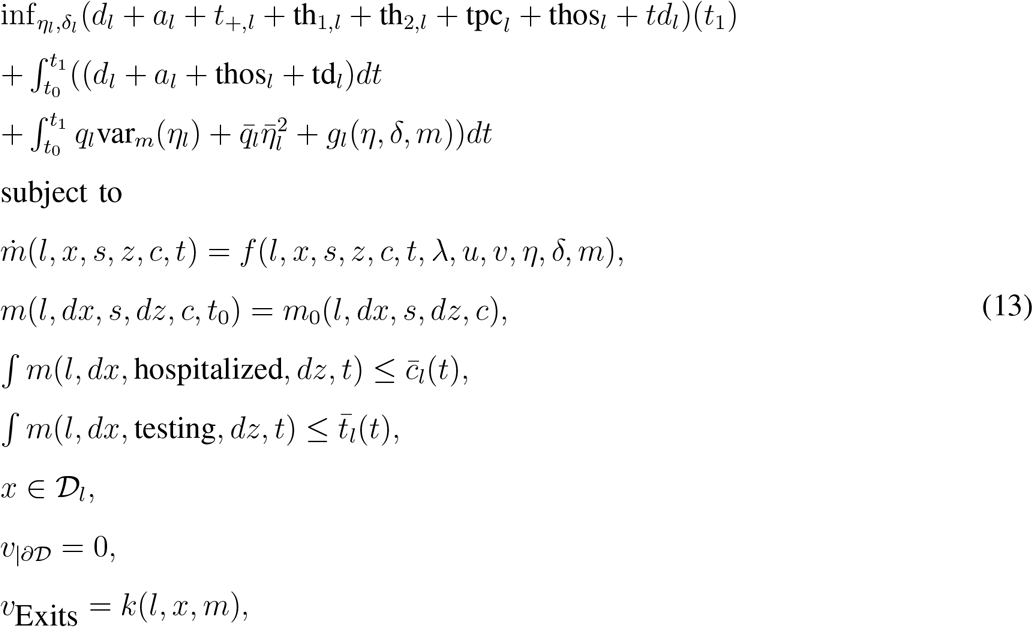

Where 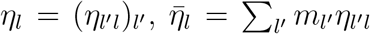 and 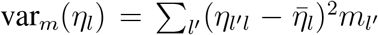 and *g*_*l*_(*η, δ, m*) is the economic cost-benefit at *l*.

#### 1) Mean-field-type equilibrium

The MFTG problem is then obtained by combining the two interdependent problems (13) and (12). A solution (*u*^∗^, *v*^∗^, *η*^∗^, *δ*^∗^) to this MFTG problem is a mean-field-type Nash equilibrium.

##### Proposition 2.

*If there is a solution to the following Hamilton-Jacobi-Bellman system in the space of*

*Measure* 𝒫_2_(ℝ^2^ × *S* × [0, 124*y*] × *C*) *given by*

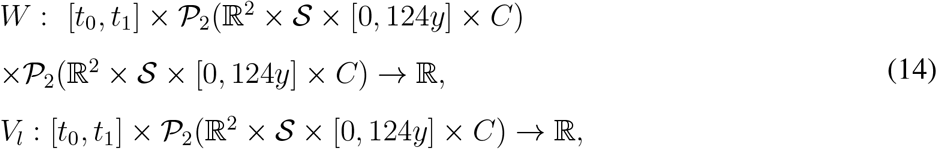

*where* 𝒫_2_ *is the set of square* (*in space and age*) *integrable* (*positive*) *measures and*

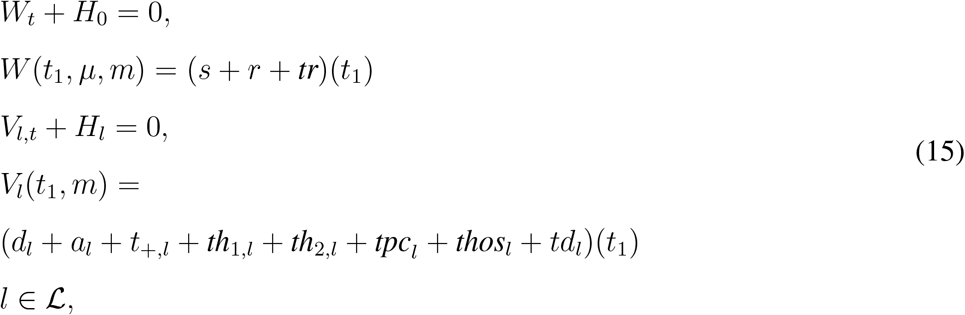

*where the Hamiltonians are given by*

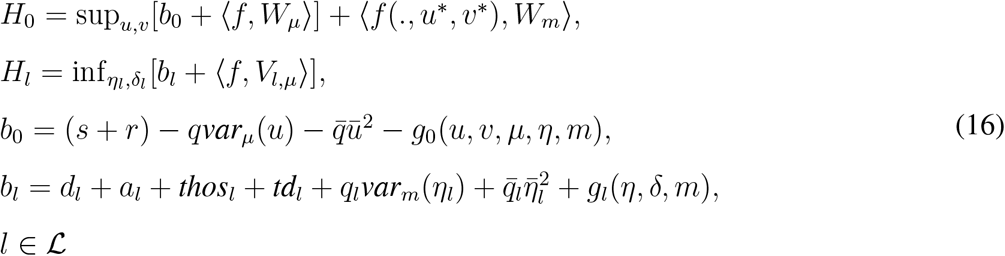

Then, (*u*^∗^, *v*^∗^, *η*^∗^, *δ*^∗^) is a mean-field-type Nash equilibrium.

The proof of Proposition 2 follows similar steps as in [20]. From the formulation of the minimization problem, the number of infected people and the number deaths are significantly reduced at the mean-field equilibrium. However, the cost-benefit analysis suggests that it depends on the functionals *g*_0_, *g*_*l*_, *h*_1_, *h*_2_.

The master adjoint system (MASS) is given by

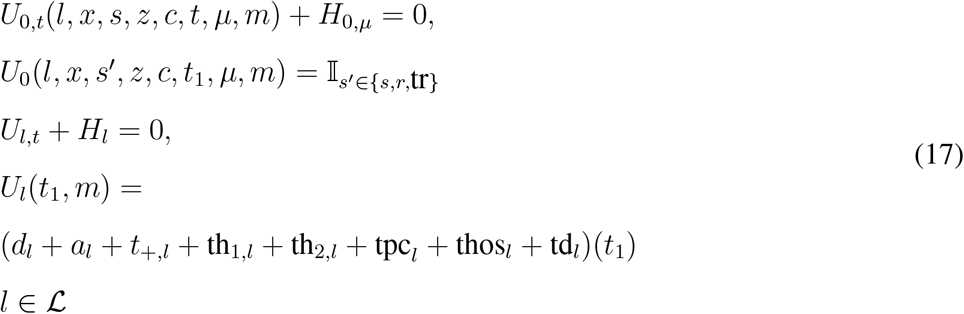

The MASS system provides the equilibrium strategies but not necessarily the equilibrium cost.

### Other relevant payoffs

- Authorities The authority in locality *l* decides on
  − Migration rules (lockdown, confinement, isolation, curfew) (*η*_1,*l*_, *η*_2,*l*_, *η*_3,*l*_), testing strategy *δ*_*l*_
  − Budget allocation and incentives *ls*_*l*_, *tr*_*l*_ Multiple objectives for authority in locality *l*:
  − reduce the number of deaths, infected,
  − reduce economic losses
  − maximize the number of recovered-deaths

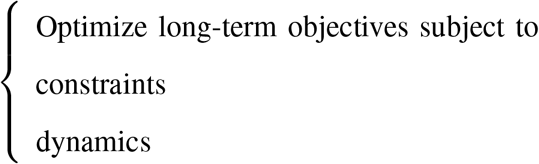
- Consumption goods firms Firms (producing essential, moderate-essential, less-essential goods) in locality *l* decide on
  − Production, Total hours worked *A*_1_ = *a*_1_*m*
  − Budget constraint Multiple objectives for *j* :
  − Reduce the number of infected employees,
  − Maximize profit

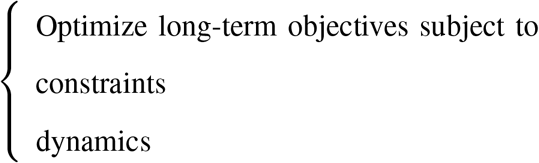
- Each individual’s risk-awareness An individual in locality *l* decides on
  − Meeting rate *a*_2_, local movement *x, v, a*_3_, consumption *a*_1_ Multiple objectives for an individual
  − reduce the risk of being infected,
  − reduce the risk of exposing the others (if co-opetitive),
  − reduce economic losses

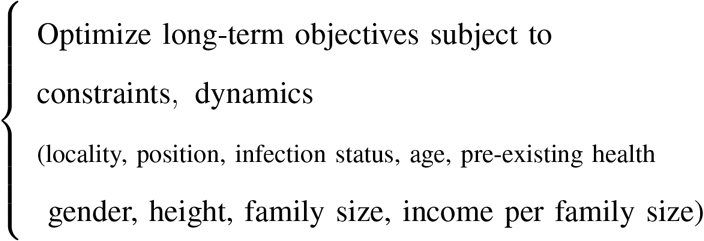
  for susceptible: economic: 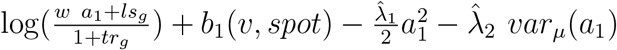 pandemic: 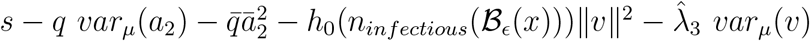

## IV. Model outputs and Data visualization

### A. Data used and constraints

We present some illustrative examples with *K* = 195 countries and more detailed analysis for 66+ countries. The COVID-19 data set is taken from [10]–[13], [33]. The age-structure data per country is provided in [32]. For the international connectivity graph, *η*_1_ we have used Flightradar24 [31], which is a global flight tracking service that provides information about flights from 1200 airlines and 4000 airports around the world. The data set does not capture all international connections but it provides a sample. As the initial propagation of COVID-19 started from flights we have modulated the flight data set to capture it. After a certain waiting time, the soft/partial/hard flight restriction decision has been made between some countries. Those countries we made *η*_1,*l*_′_*l*_ to be zero or a very small number (0.001) if travels from *l*′ to *l* are restricted. For the geographical mobility and co-location map of people in country *l*_1_, we have used OpenStreetMap before and after confinement decisions (if any) per country. The Google Community Mobility Reports can be found in [34].

The country-specific hospital capacity data set is obtained from World Bank Group [35] and is used to capture

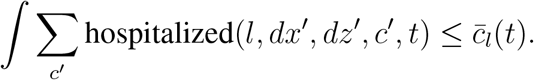

However, the data set is limited to the country level and it is not per area/city/county. The country-specific testing capacity data set is not available but the number of tests per country is provided in [12]. The set 𝒟 = *∪*_*l*∈ℒ_𝒟_*l*_ where 𝒟_*l*_ is a constrained non-empty set. Given two countries *l*_1_ and 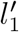, the dynamics *m*(*l, x, s, z, c, t*) and *m*(*l, x, s, z, c, t*) will be related to each other via the switching rates *η*_1_. Table II summarizes the heterogenous data sources. Figure 12 represents the proportion of infected people

**Fig 12:**
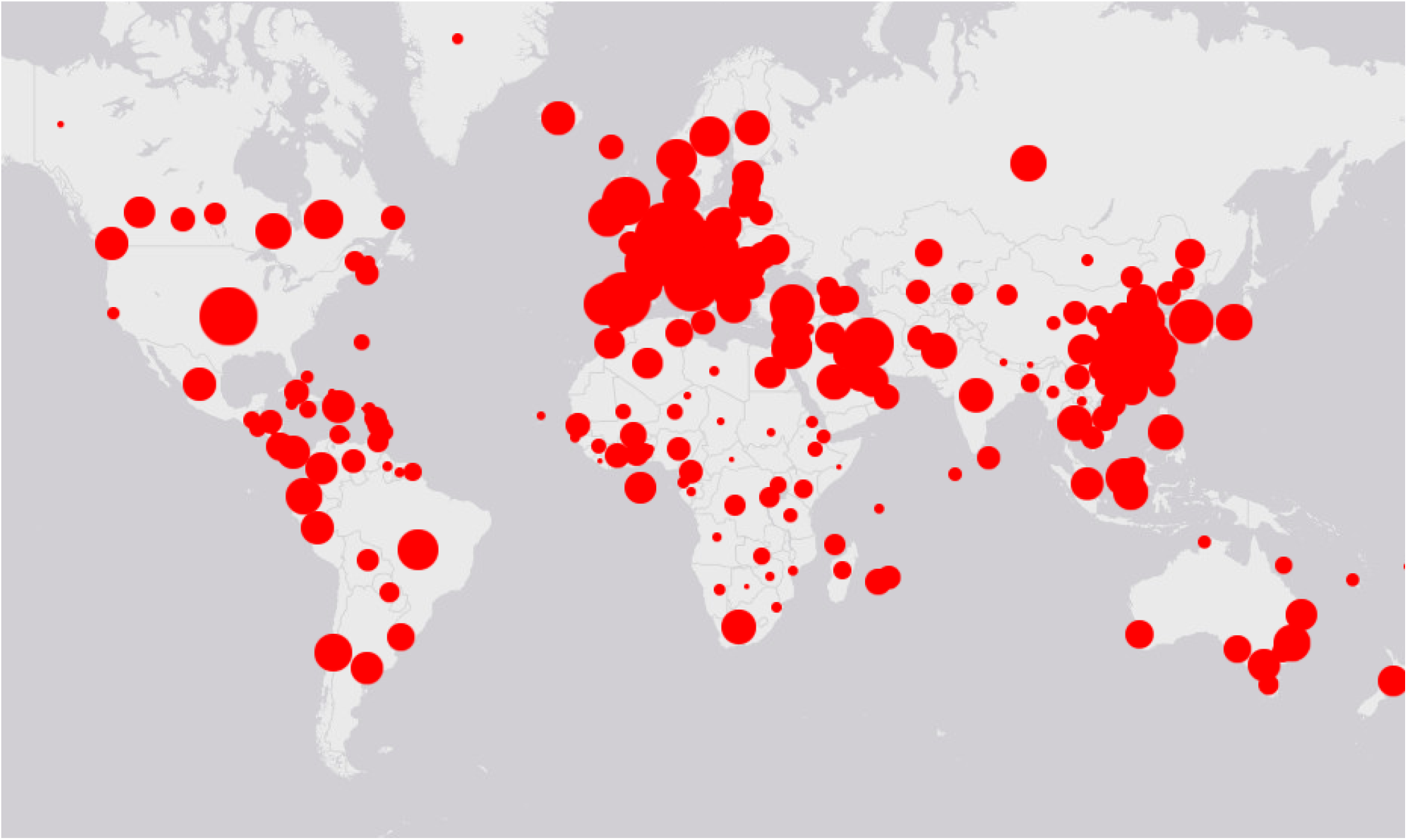
World map: reported infectious as of March 30, 2020

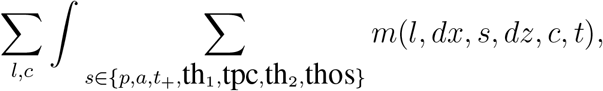

at the 195 countries as of March 30, 2020. Figure 13 illustrates the status of the fifteen most (reported) infected states as of April 1, 2020. Figure 14 samples G7 countries. Detailed testing results are displayed in Figure 15 for the cities of New York and Los Angeles.

**Fig 13:**
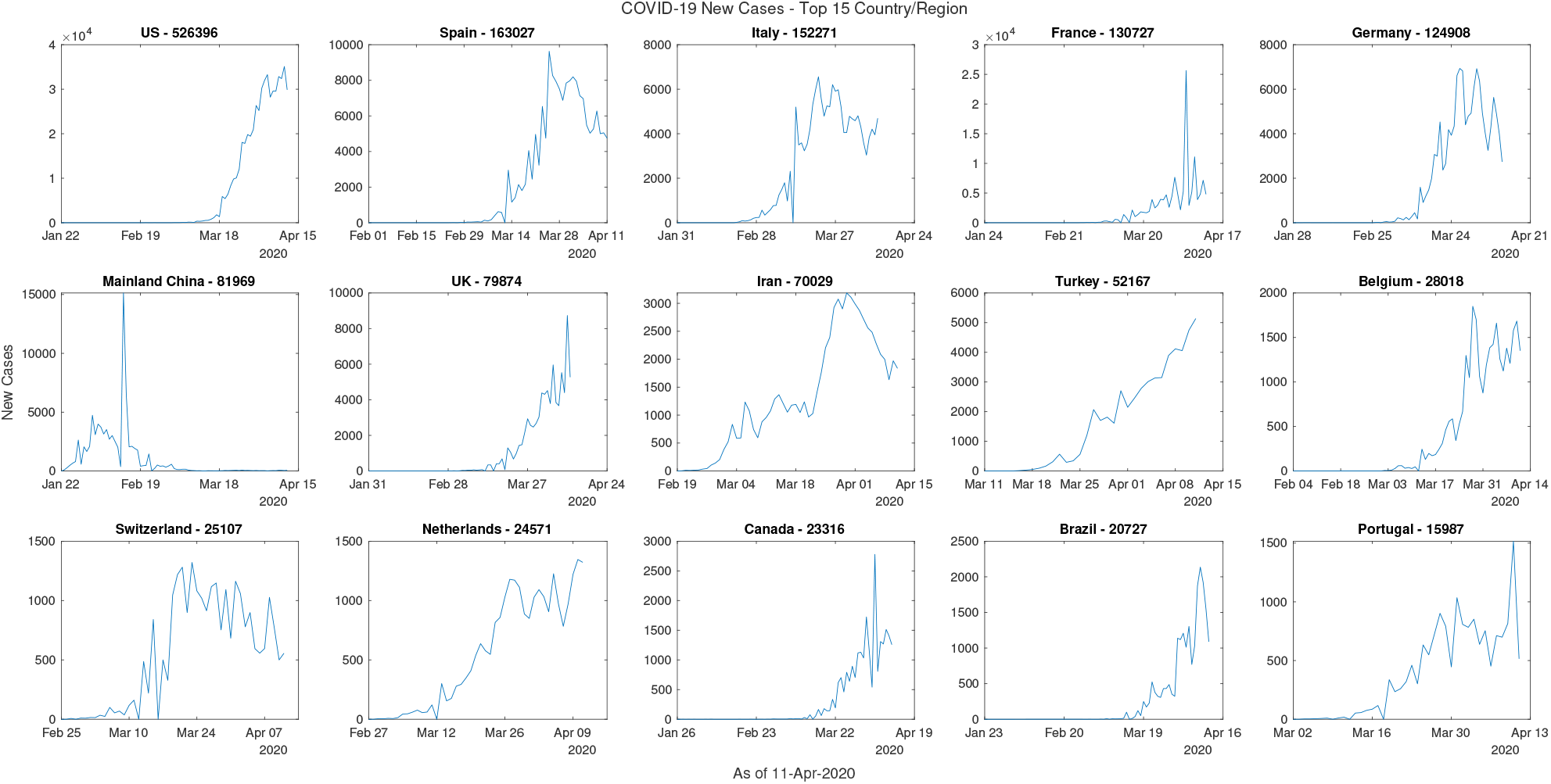
Propagation of the virus in 15 most reported active cases as of April 11, 2020.

**Fig 14:**
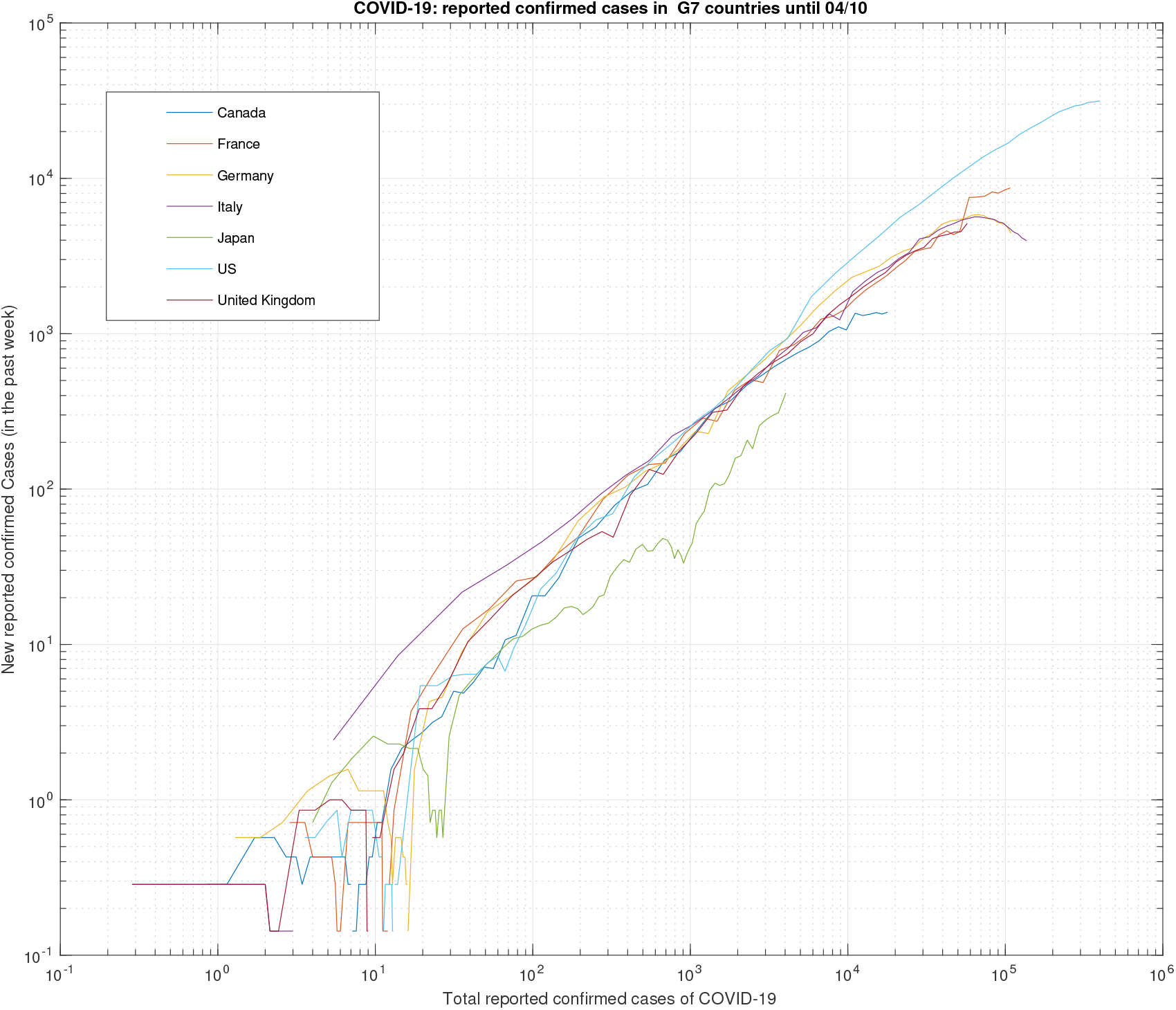
COVID-19 samples in G7 countries

**Fig 15:**
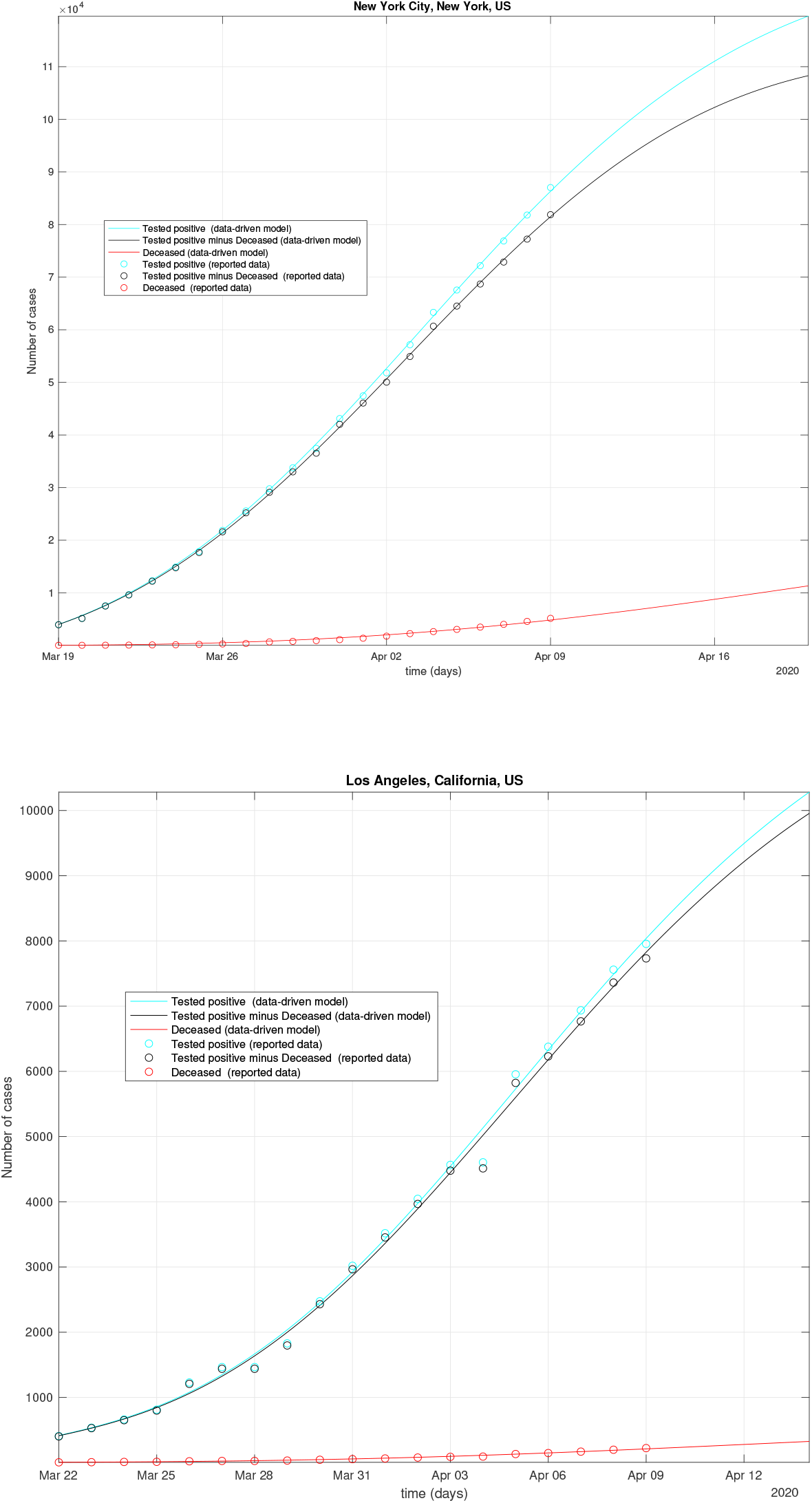
New York and Los Angeles testing results: data vs model.

We make the following observations form the model.

- the model is flexible enough to capture some local context-awareness including local mobility and age structure per country. The provided model can be filtered to track the data set provided in [12] even for some of the countries with few number of cases.
- The connectivity of the local graph *η*_3_ plays a key role in the propagation of the virus across different areas of the city
- The connectivity of the local graph *η*_2_ plays a key role in the propagation of the virus across different cities
- The connectivity of the local graph *η*_1_ plays a key role in the propagation of the virus across states/countries.
- The current case fatality rate (data-CFR) per country data set is the ratio between the number of (reported) confirmed cases and the total number of (reported) deaths from COVID-19 outbreak data set [33] for that country. We observe that there are some significant differences in terms of data-CFR per country even for whose who have made similar decisions at similar infection time. Some of the differences comes from the implementation in practice and the response of the people to the measures. The proposed MFTG model captures the data-CFR per country as illustrated in 66+ countries.

### B. Visualization

The optimization algorithm is implemented in C++ using [41]. The plots on the geographical-map (see Figure 12) are done using MATLAB. We combine several layers: the transportation layer (air and ground), economics layer, population mobility map after the first infection of COVID-19 in that city/country, and the epidemic data-driven model. We display below the outputs (tested positive, tested-recovered, tested-dead, tested-active) of the epidemic model in 66+ countries. We choose the constraints 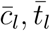 to be high enough so that the interior solution can be found. This means extending the capacity by means of new resources. The COVID-19 initial death rate is chosen as in Figure 3 for the countries where the death rate due to COVID-19 is unknown. We estimate the death rate from the model using the reported data [12]. We choose *k* = 0.7*m*(.) for the numerics at the exits on the grid over OpenStreet map. The rate *λ*_*ap*_ is set 10^−7^ or zero depending on the country/locality. The other *λ* functions are obtained from the minimization with the smallest norm allowed to derive *λ*data. As new data comes, *λ*data can be recalculated. The newborns are taken from the last year growth scale. From *λ*data we plot the behavior of the dynamics and integrates it out. The initial *t*_0_ is set to January 22, 2020 for most countries.

We observe that the proposed model captures the behavior in most countries. The data-driven MFTG model reveals that the number active COVID-19 positive patients in countries such as Venezuela, Germany, Italy, Malaysia, Senegal, Zambia, Togo have several local peaks and oscillating behaviors. Some of them cannot be fitted with a Gaussian or bi-Gaussian. It cannot be fitted with a single exponential. By changing *h*_*l*_ from 0.1 to 0.9 we observe a limit cycle of the dynamics. Figures 89 and 90 represents a COVID-19 sample of Non-Gaussianity and non-exponential. Also, Non-Gausianity of the reported active cases and hospitalized cases are observed in 15+ countries. It includes Albania, Azerbaijan, Bahamas, Bosnia, Costa Rica, Djibouti, Georgia, Irak, Iran, Jordan, Lebanon, Lituana, Malta, San Marino, Uzbekistan, etc. Tables IV, V, VI summarize lists of figures and illustrations.

**Fig 16:**
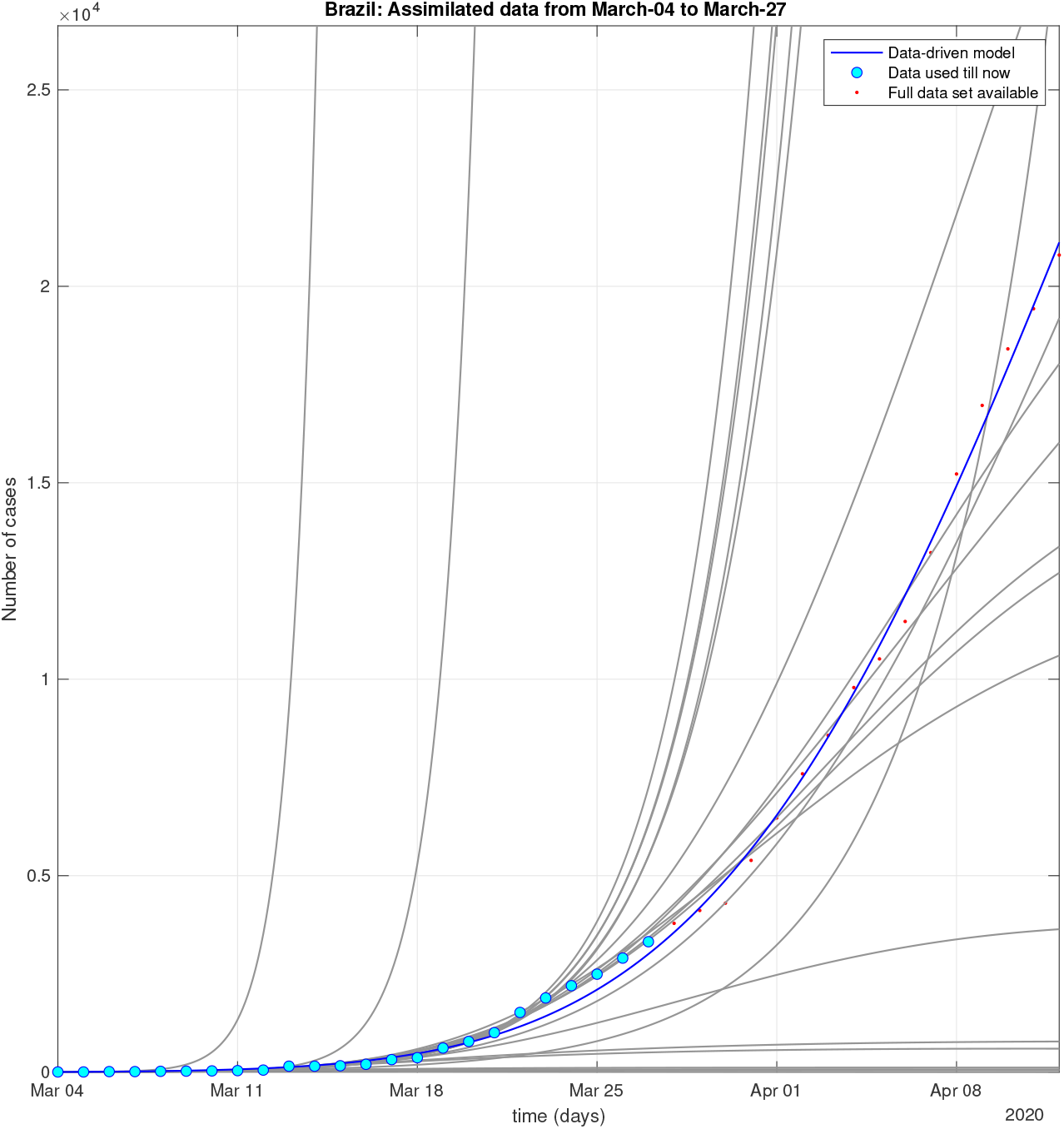
Brazil: Sequential data-driven model.

**Fig 17:**
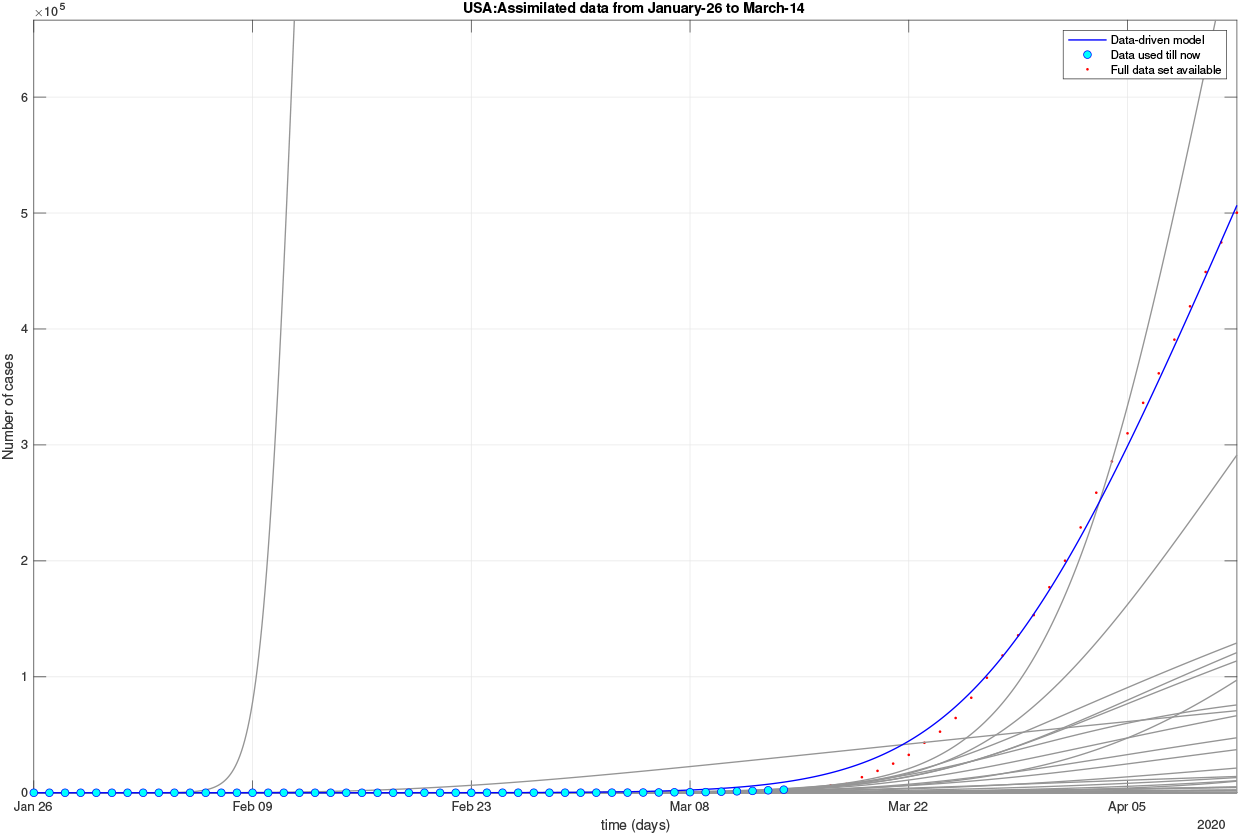
USA: Sequential data-driven model.

**Fig 18:**
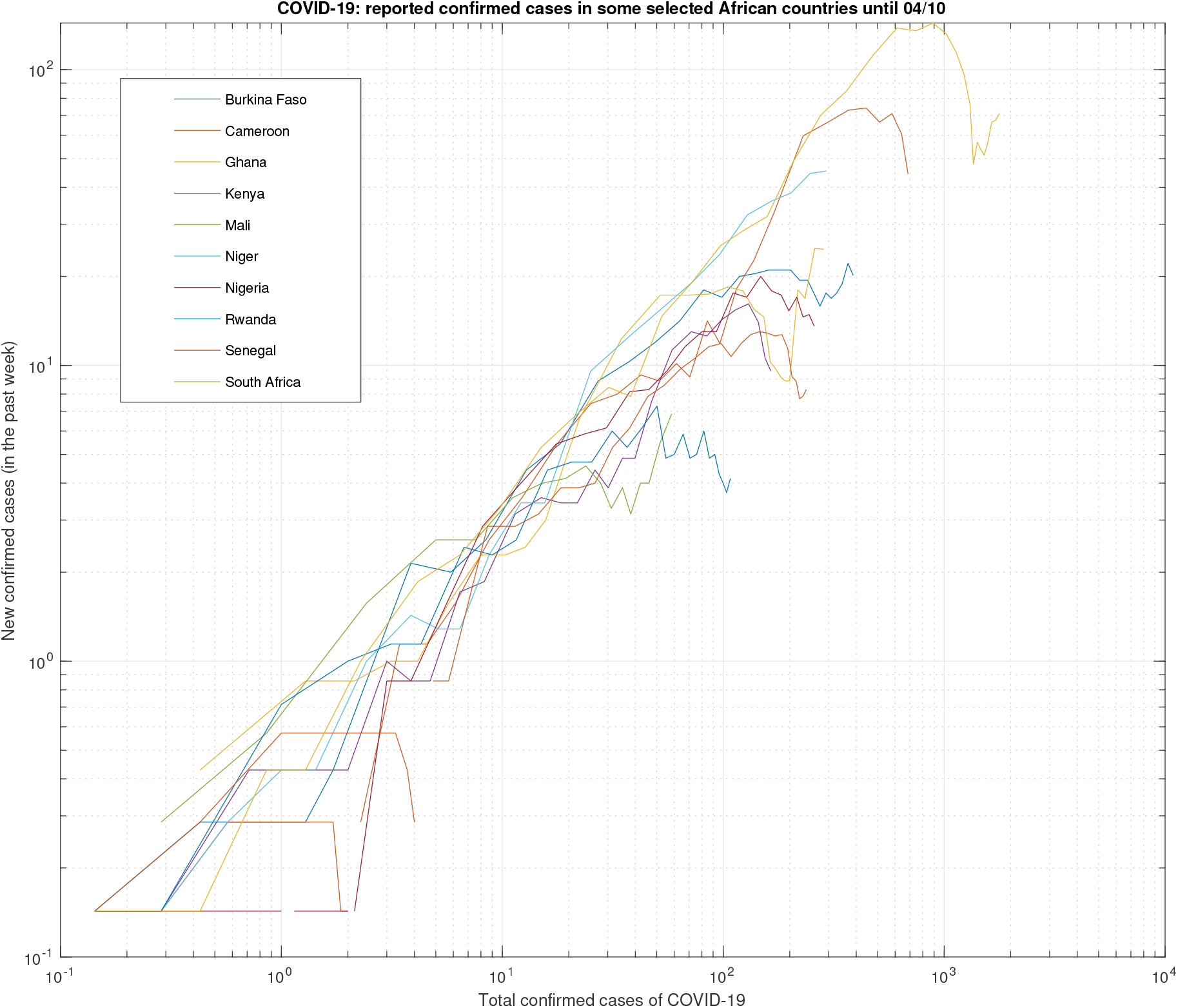
Covid-19 samples in selected countries in Africa

**Fig 19:**
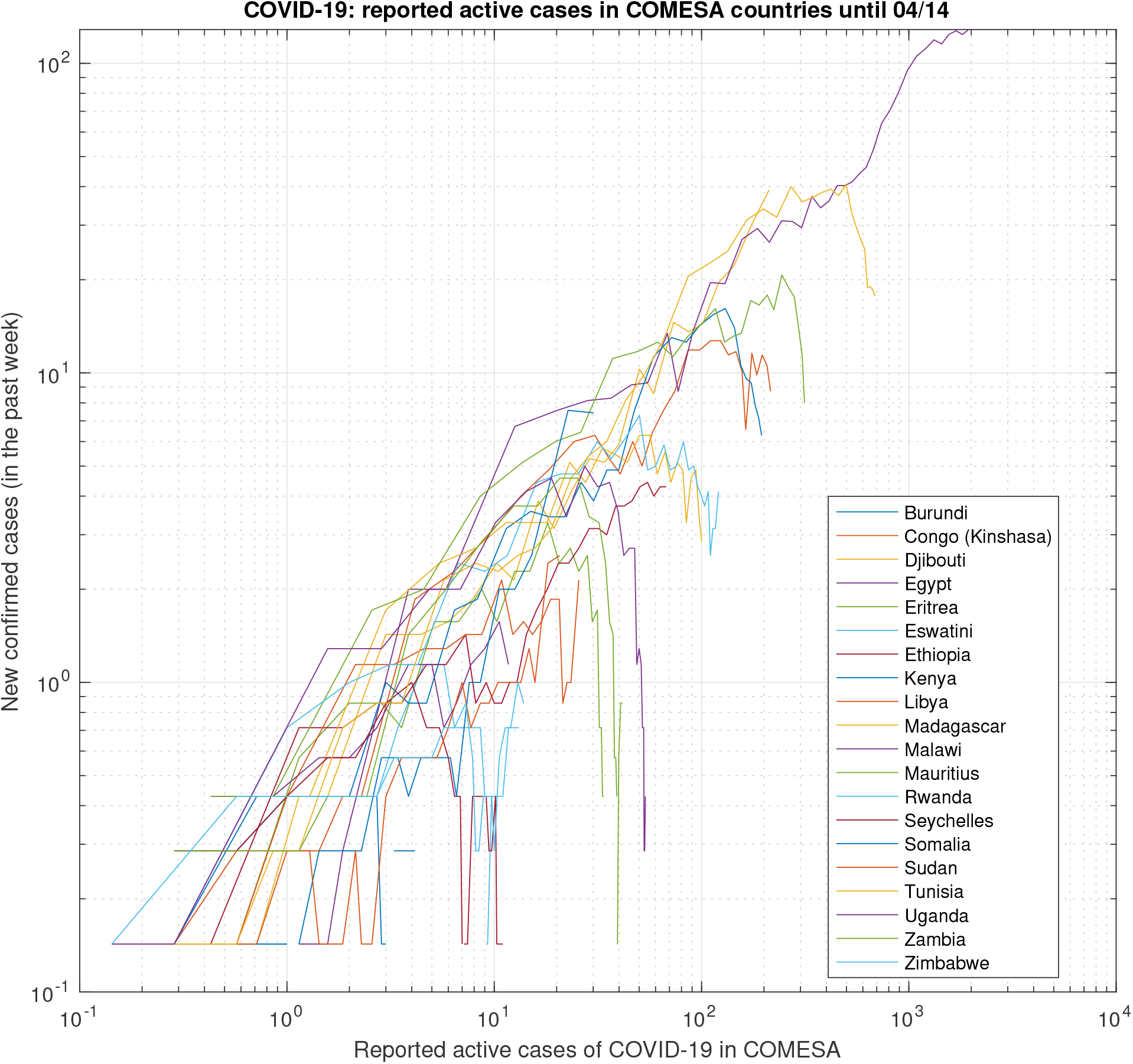
Selected COMESA countries

**Fig 20:**
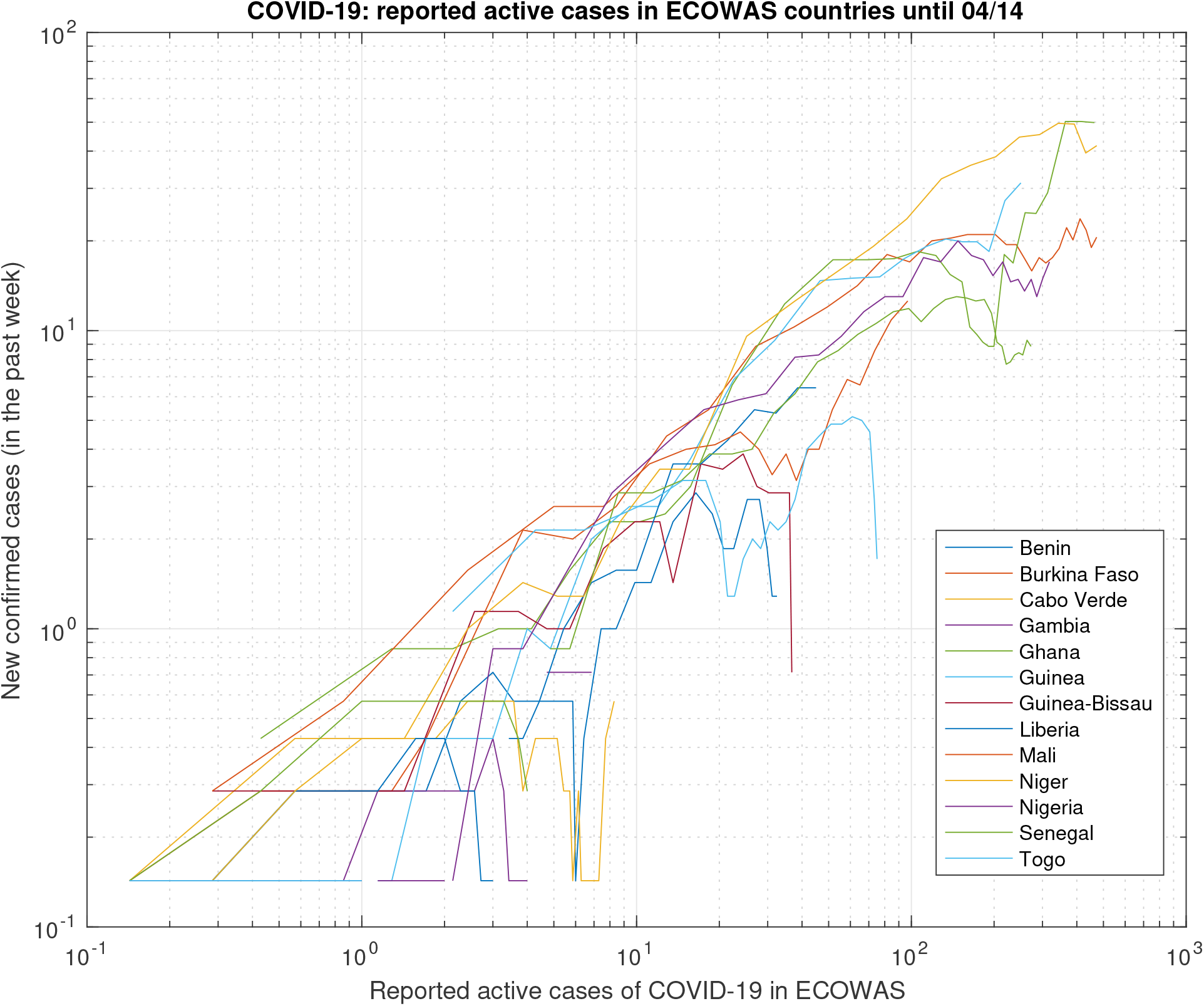
Selected ECOWAS countries

**Fig 21:**
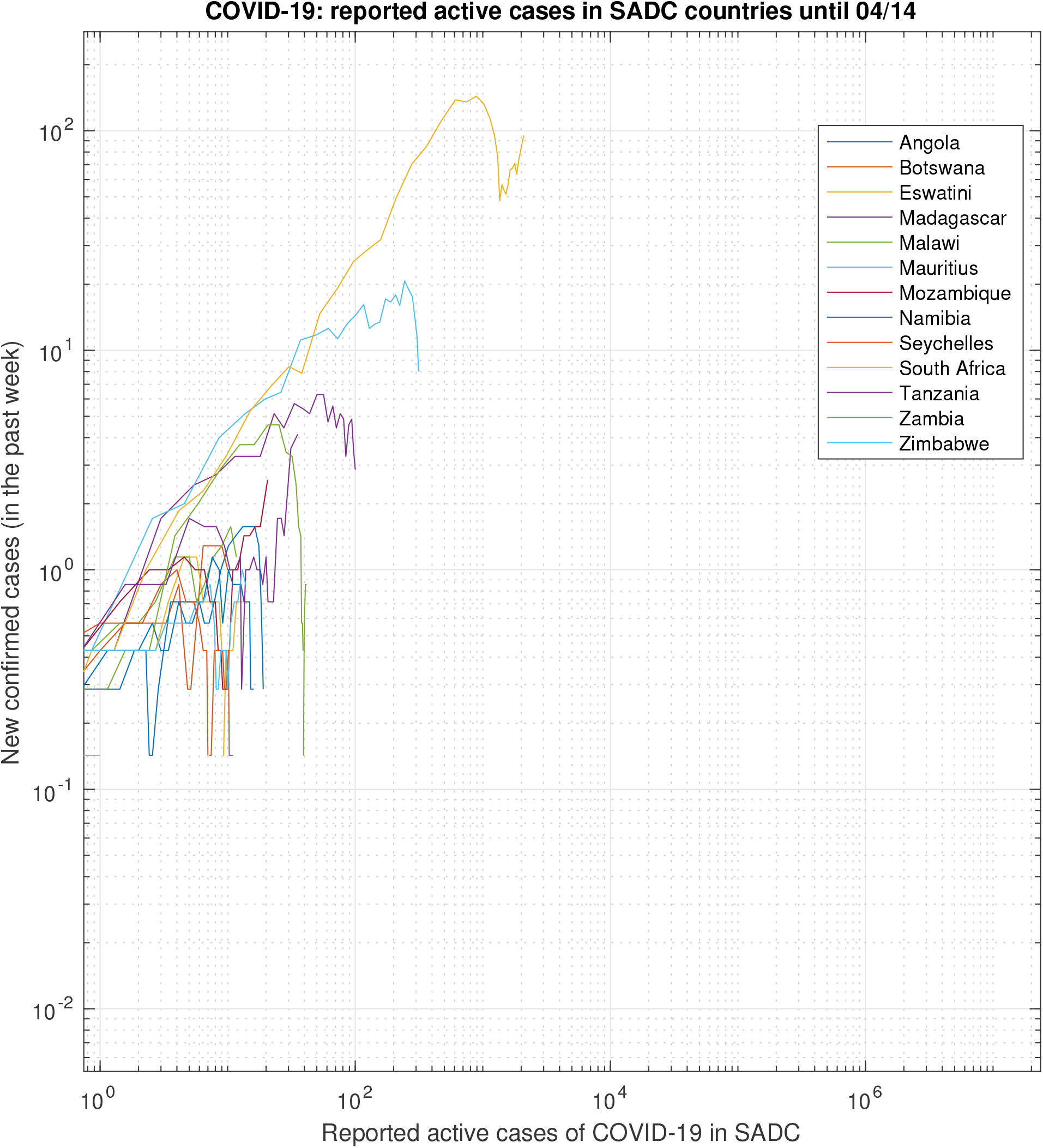
Selected SADC countries

**Fig 22:**
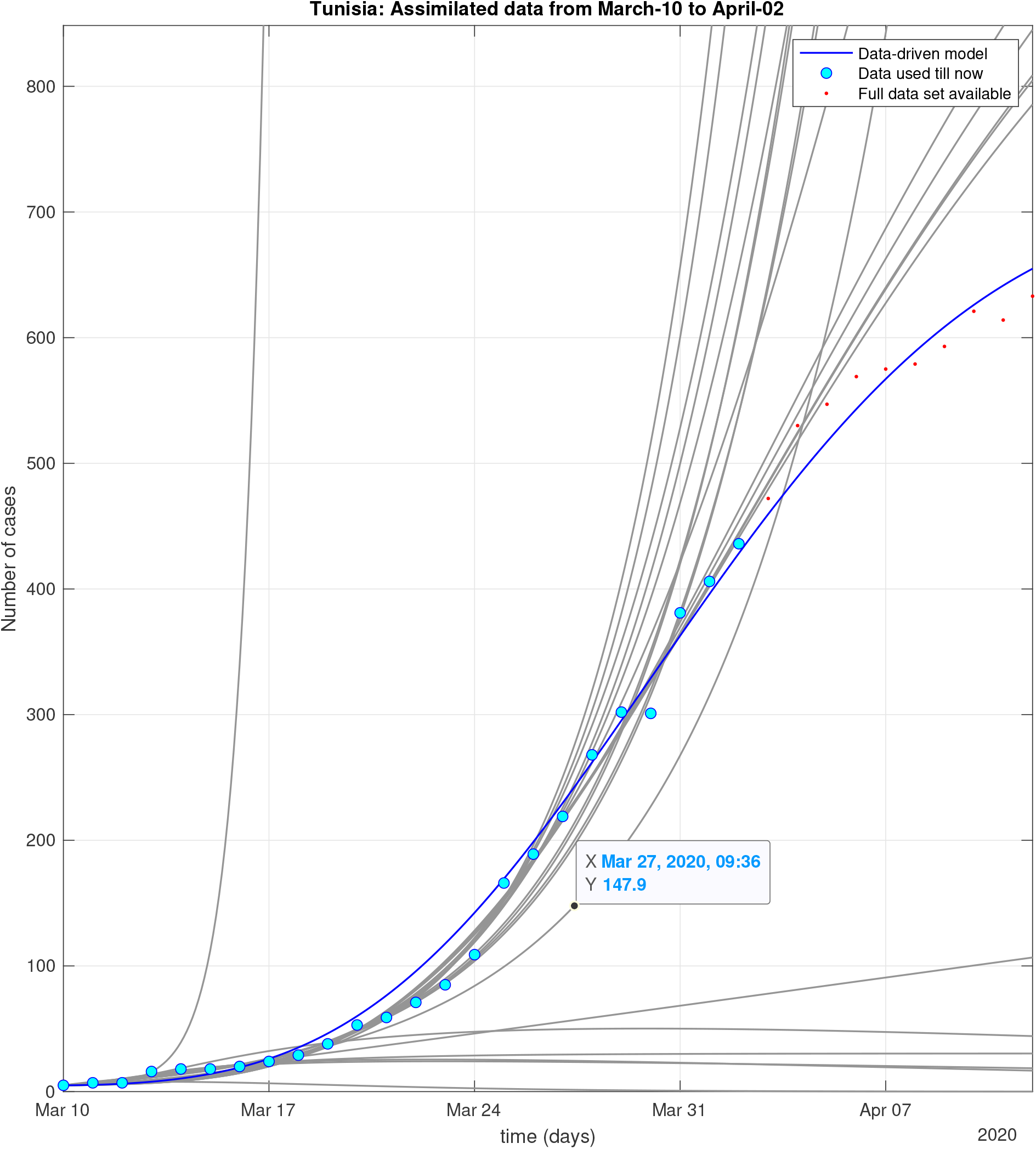
Tunisia: Sequential data-driven model

**Fig 23:**
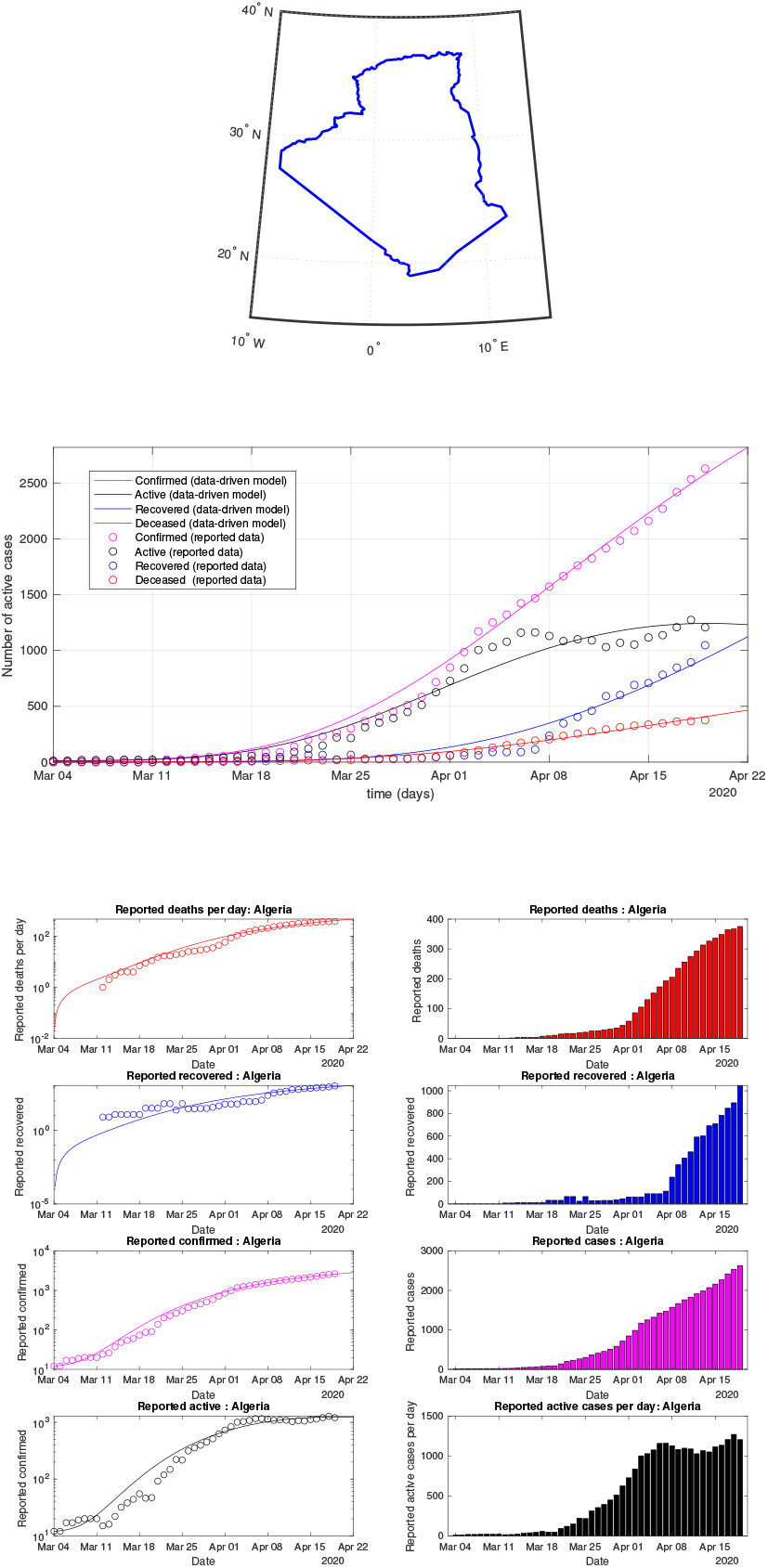
Algeria: data of active, recovered, deceased (in circle or bar) vs model of active, recovered, deceased over time. Tracking the number of active cases. Predictive analytics for the next couple days

**Fig 24:**
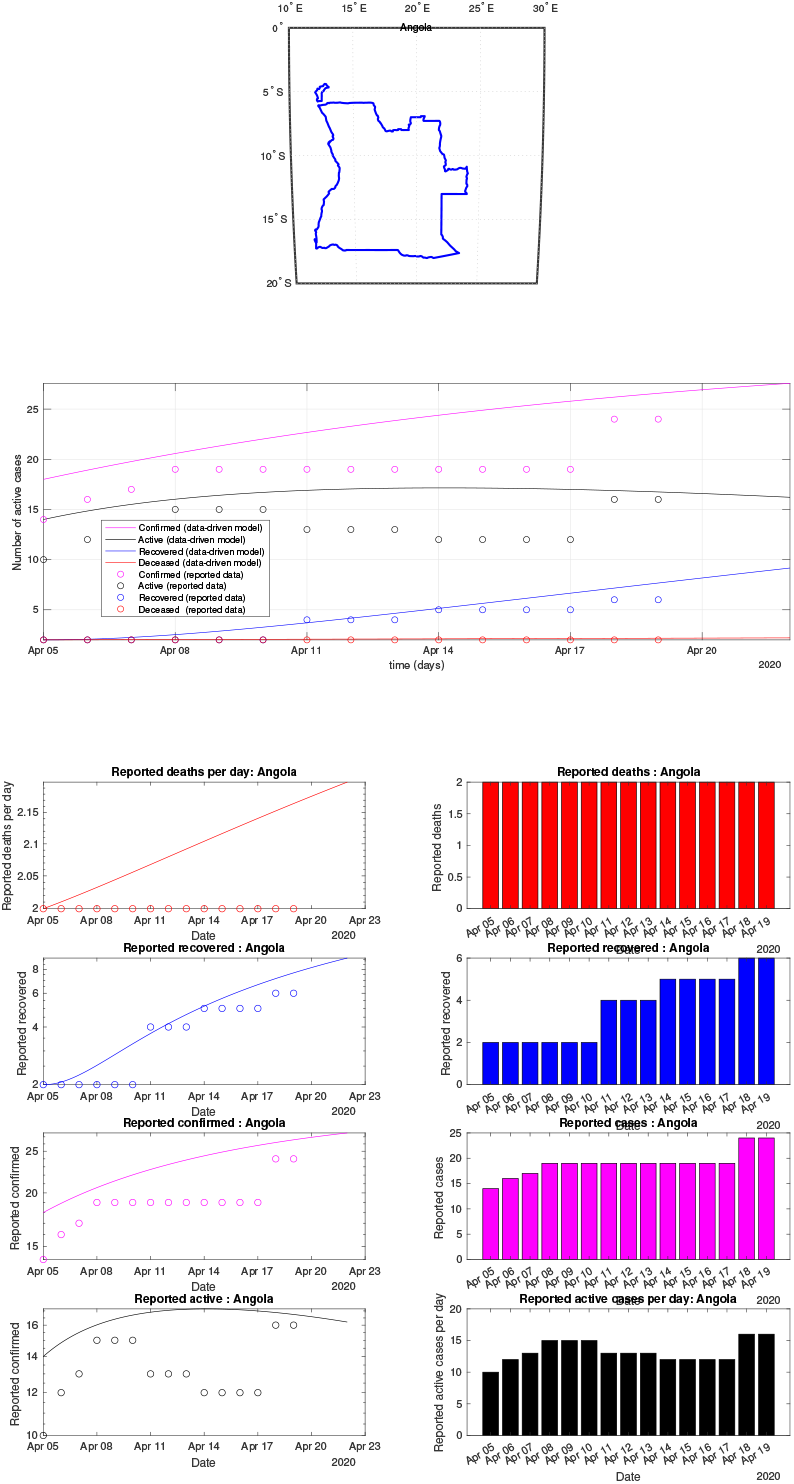
Angola: data of active, recovered, deceased (in circle or bar) vs model of active, recovered, deceased over time. Tracking the number of active cases. Predictive analytics for the next couple days

**Fig 25:**
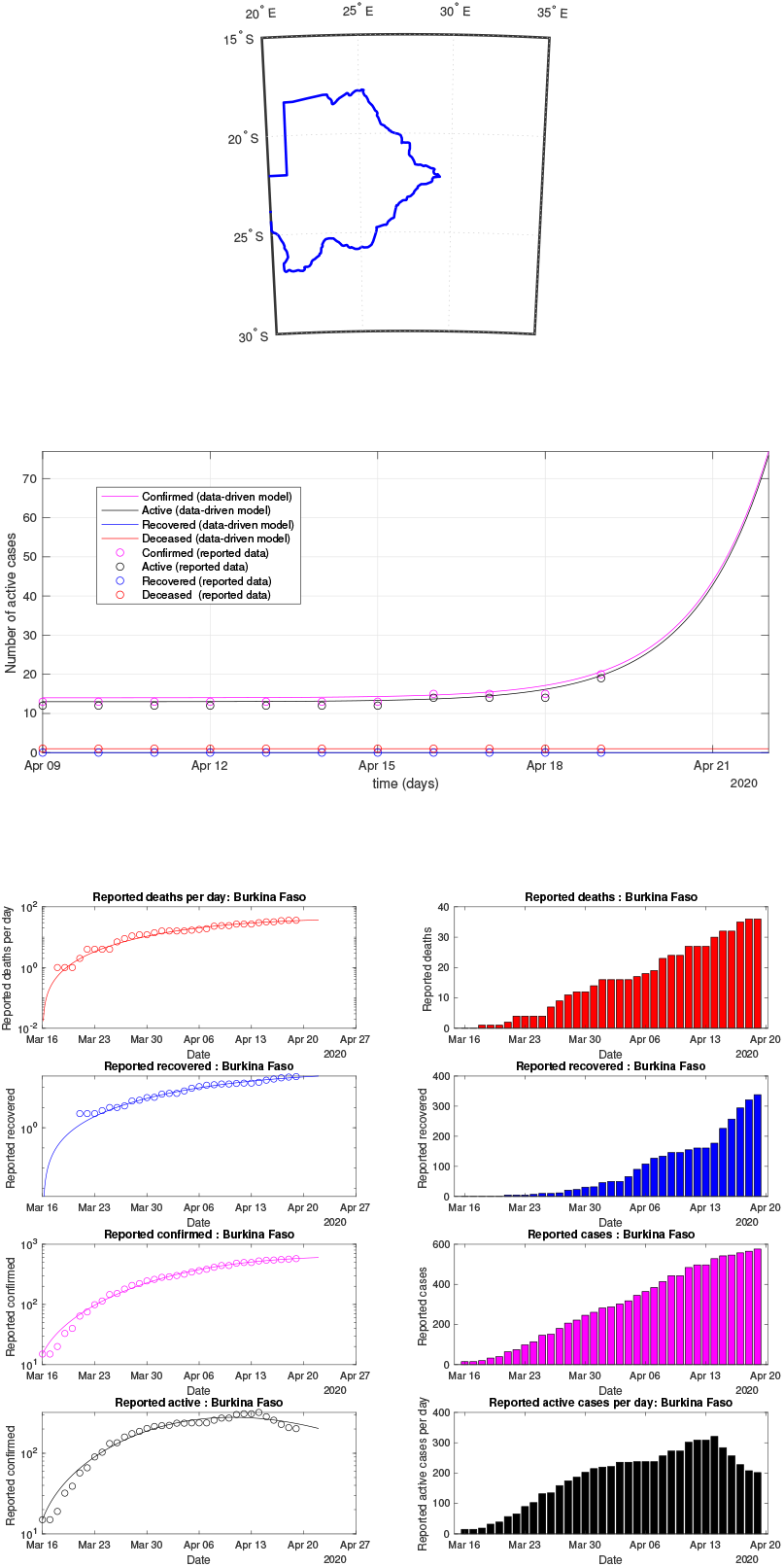
Botswana: data of active, recovered, deceased (in circle or bar) vs model of active, recovered, deceased over time. Tracking the number of active cases. Predictive analytics for the next couple days

**Fig 26:**
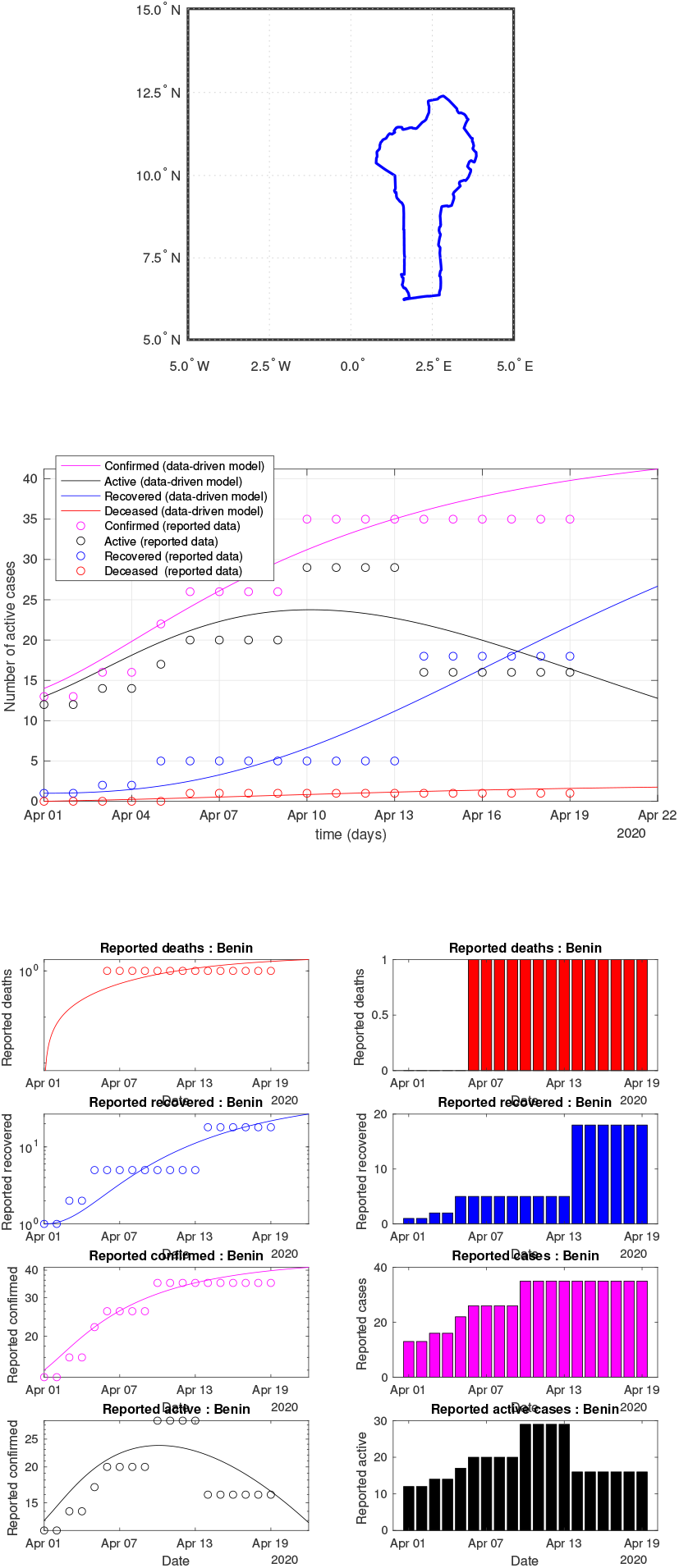
Benin: data of active, recovered, deceased (in circle or bar) vs model of active, recovered, deceased over time. Tracking the number of active cases. Predictive analytics for the next couple days

**Fig 27:**
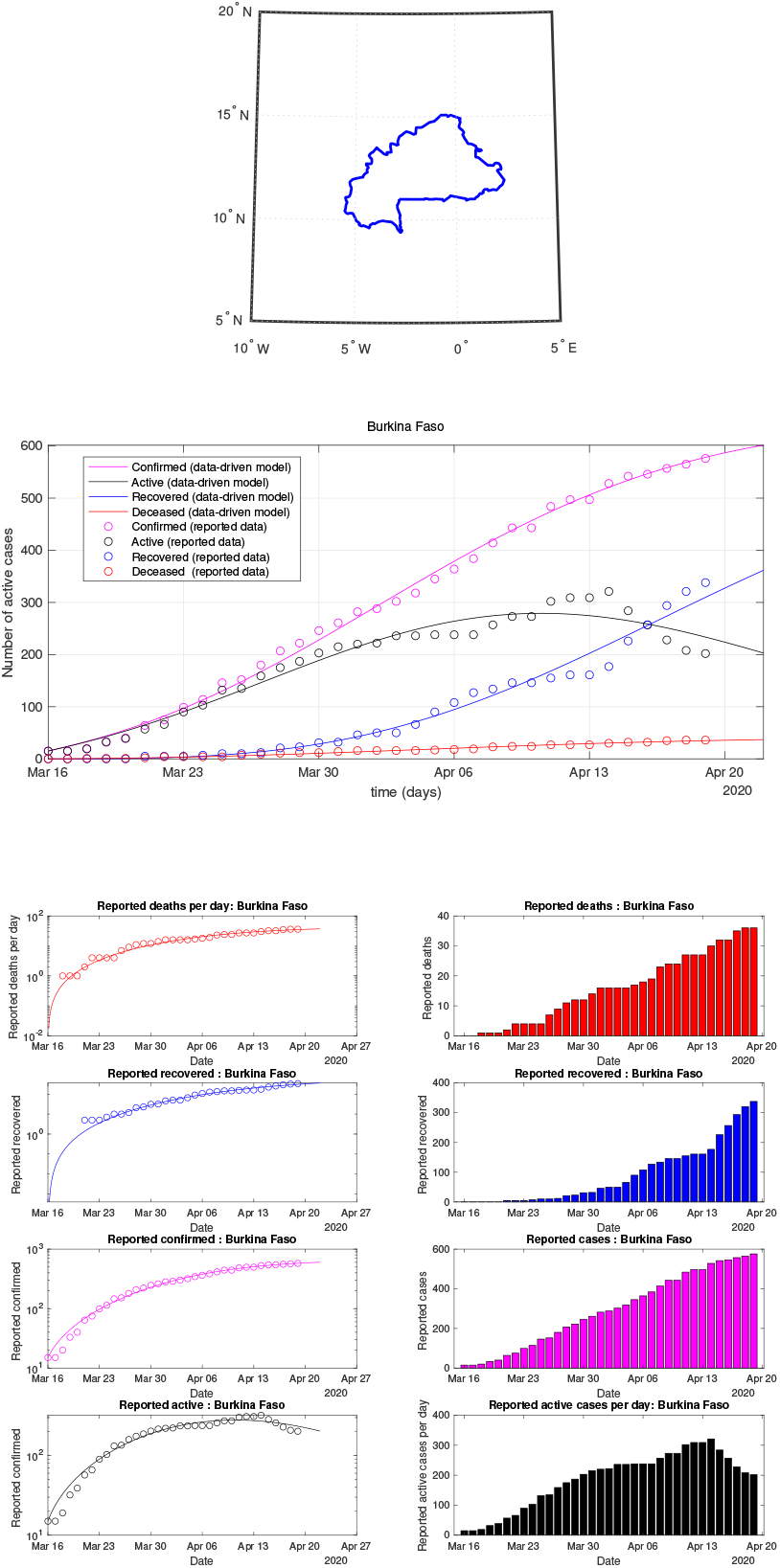
Burkina Faso: data of active, recovered, deceased (in circle or bar) vs model of active, recovered, deceased over time. Tracking the number of active cases. Predictive analytics for the next couple days

**Fig 28:**
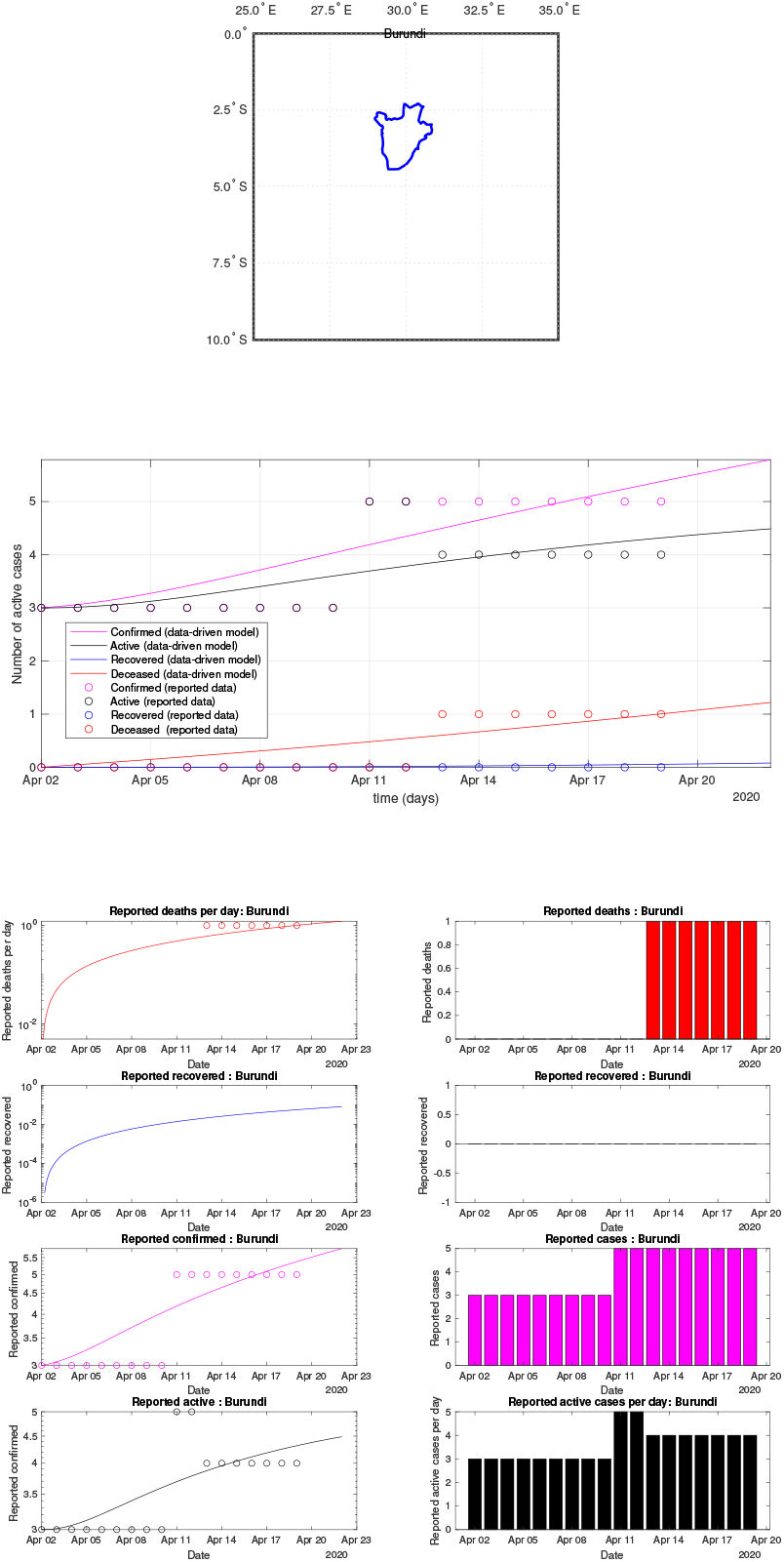
Burundi: data of active, recovered, deceased (in circle or bar) vs model of active, recovered, deceased over time. Tracking the number of active cases. Predictive analytics for the next couple days

**Fig 29:**
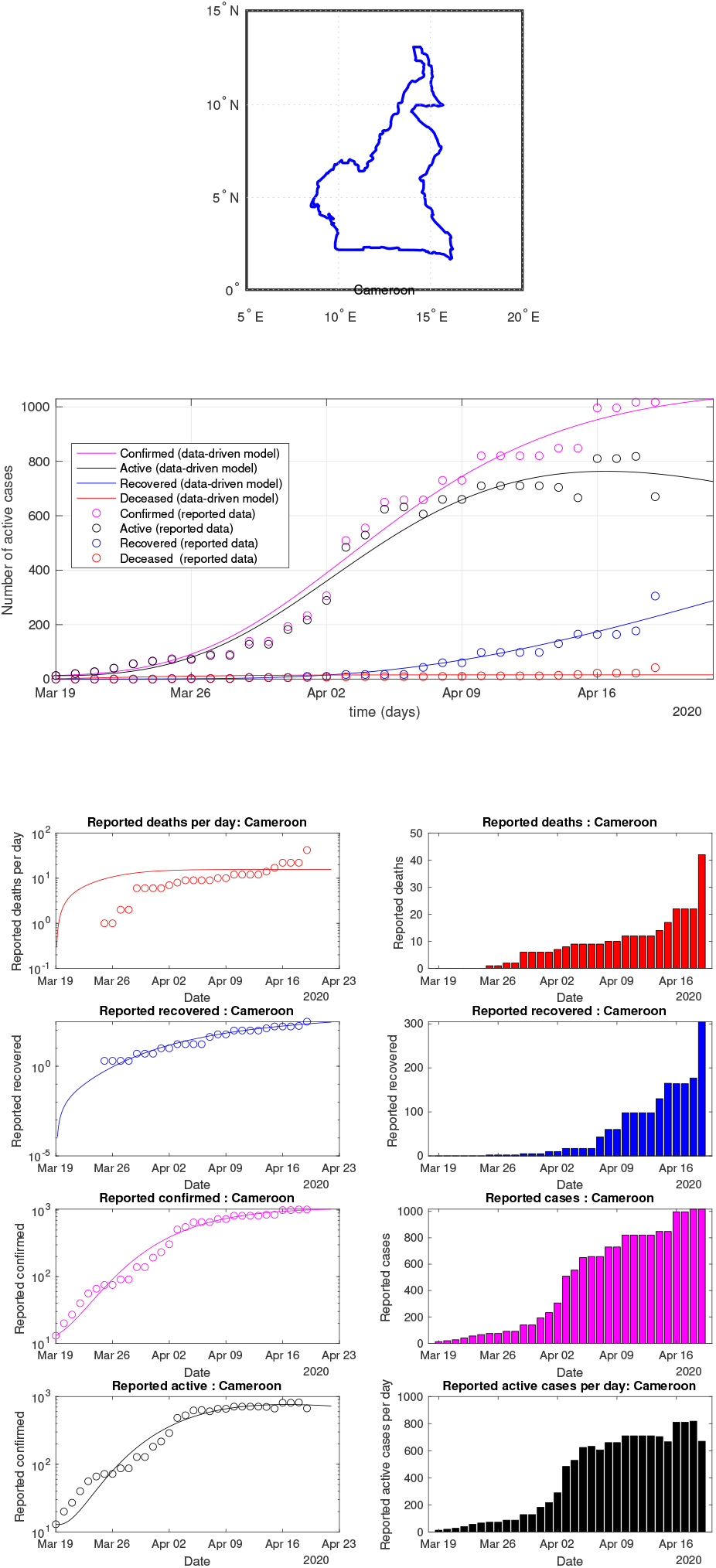
Cameroon: data vs model

**Fig 30:**
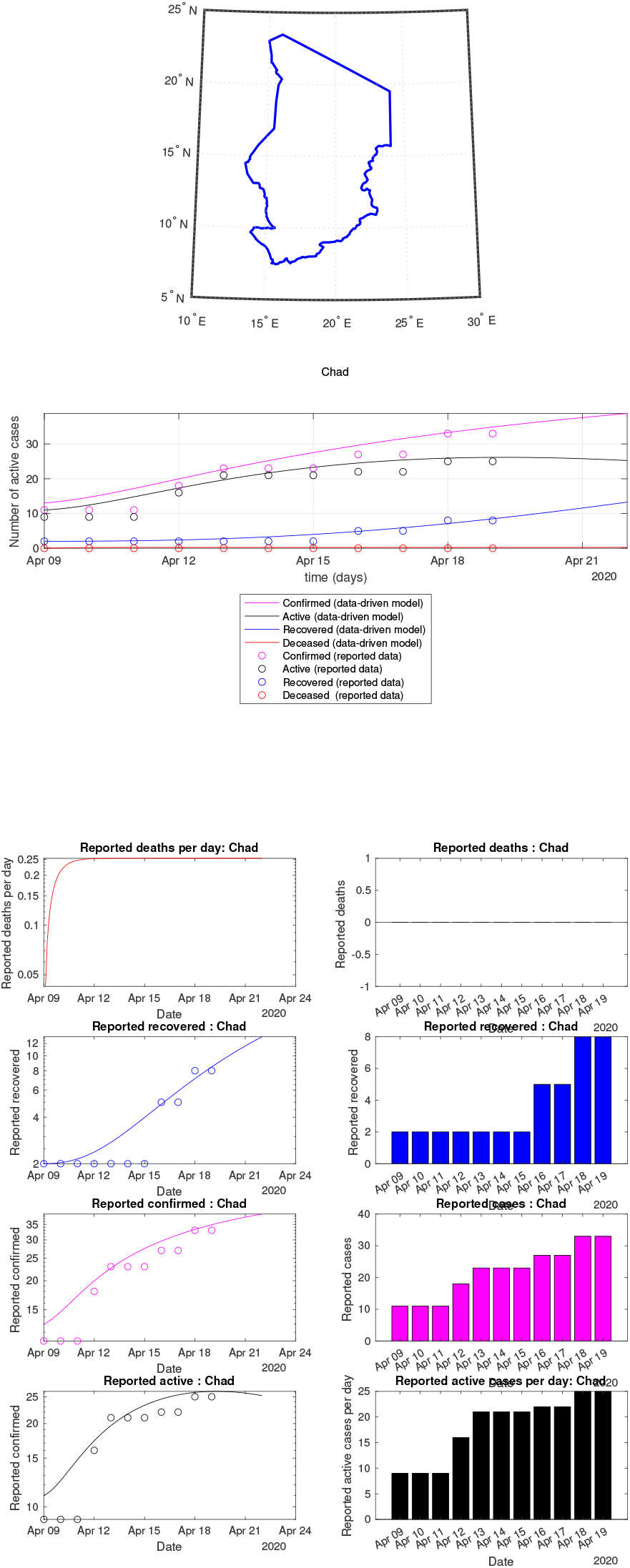
Chad: data vs model

**Fig 31:**
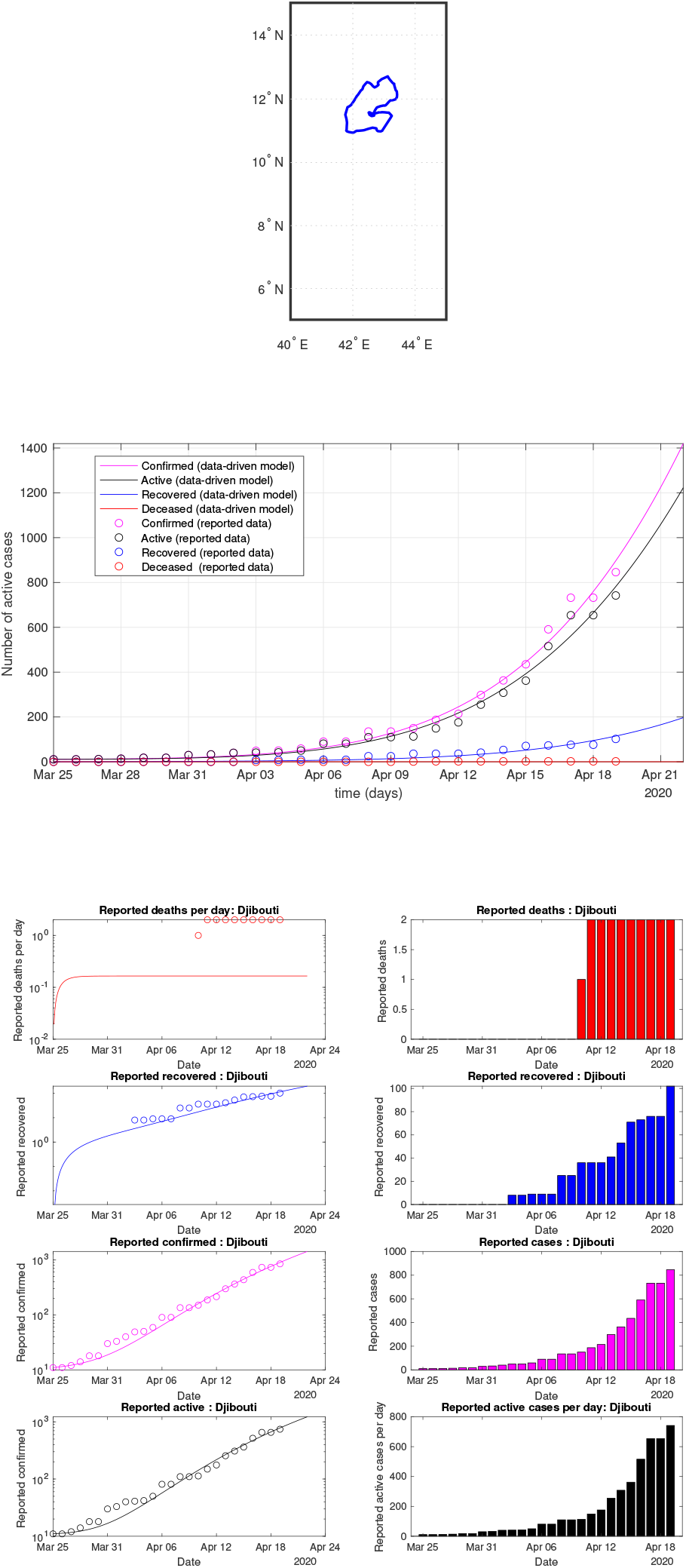
Djibouti: data vs model

**Fig 32:**
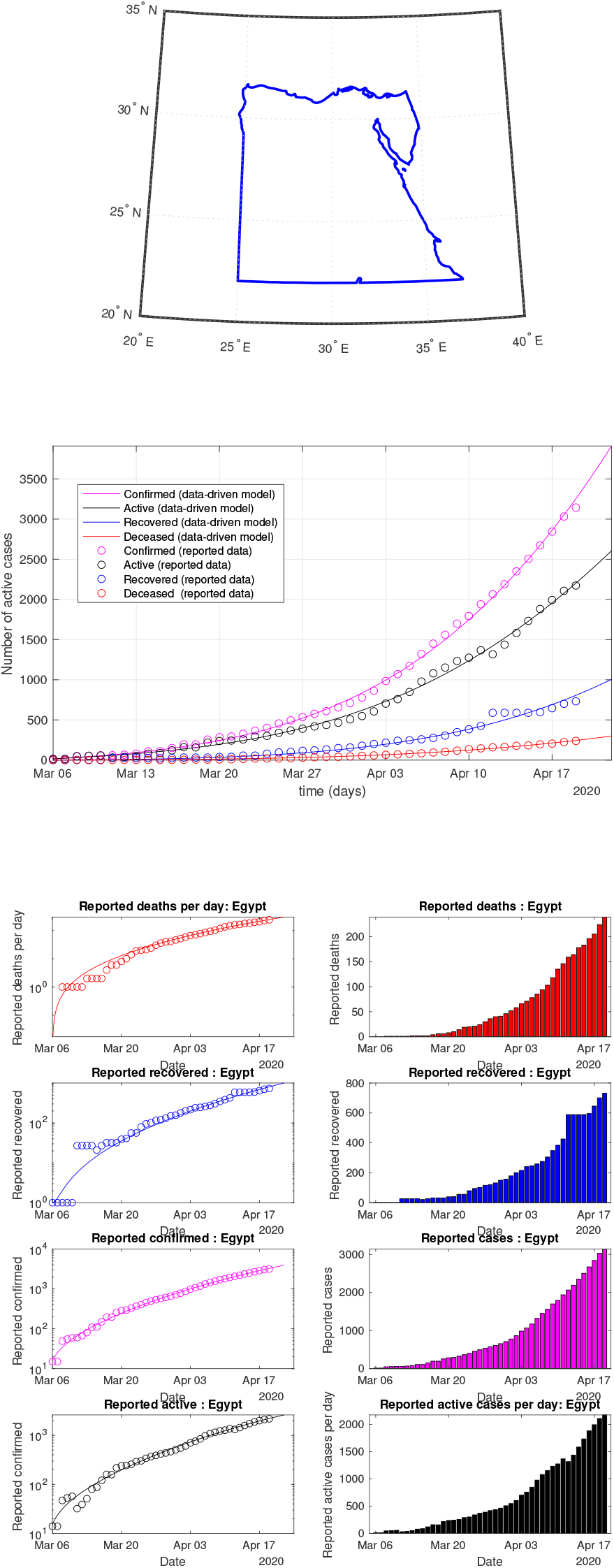
Egypt: data vs model

**Fig 33:**
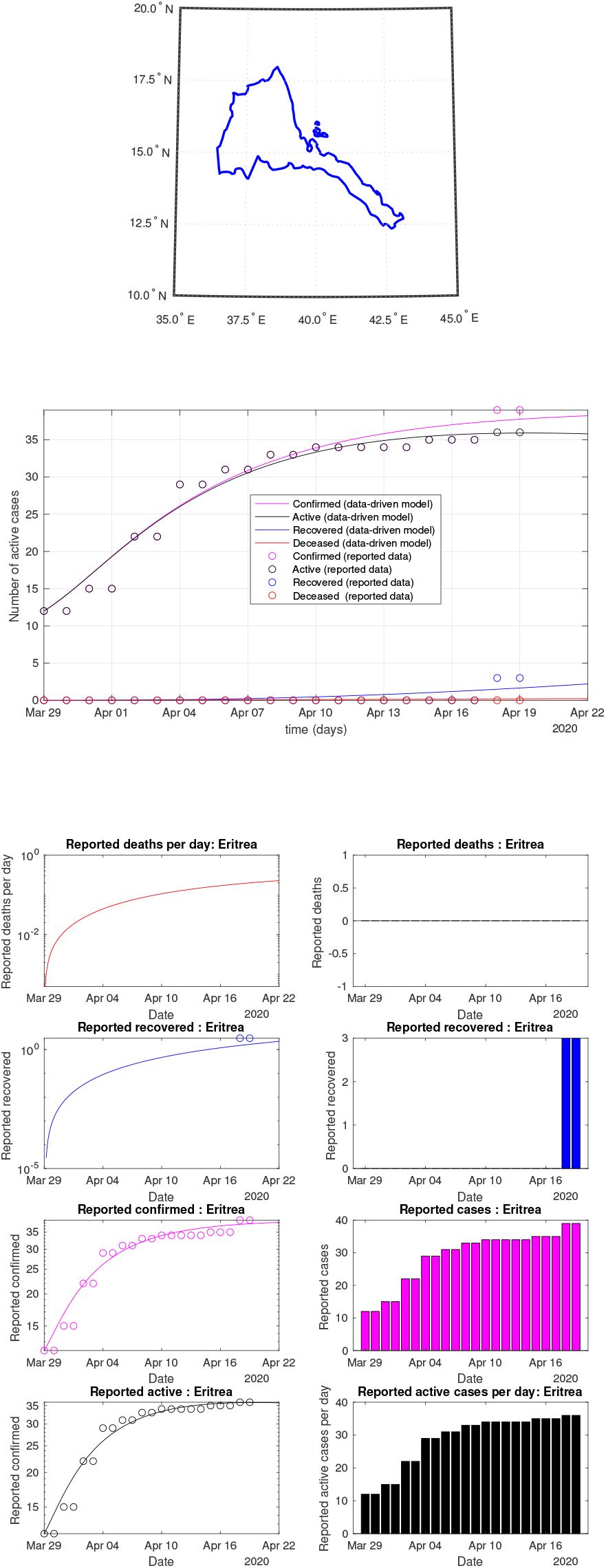
Eritrea: data vs model

**Fig 34:**
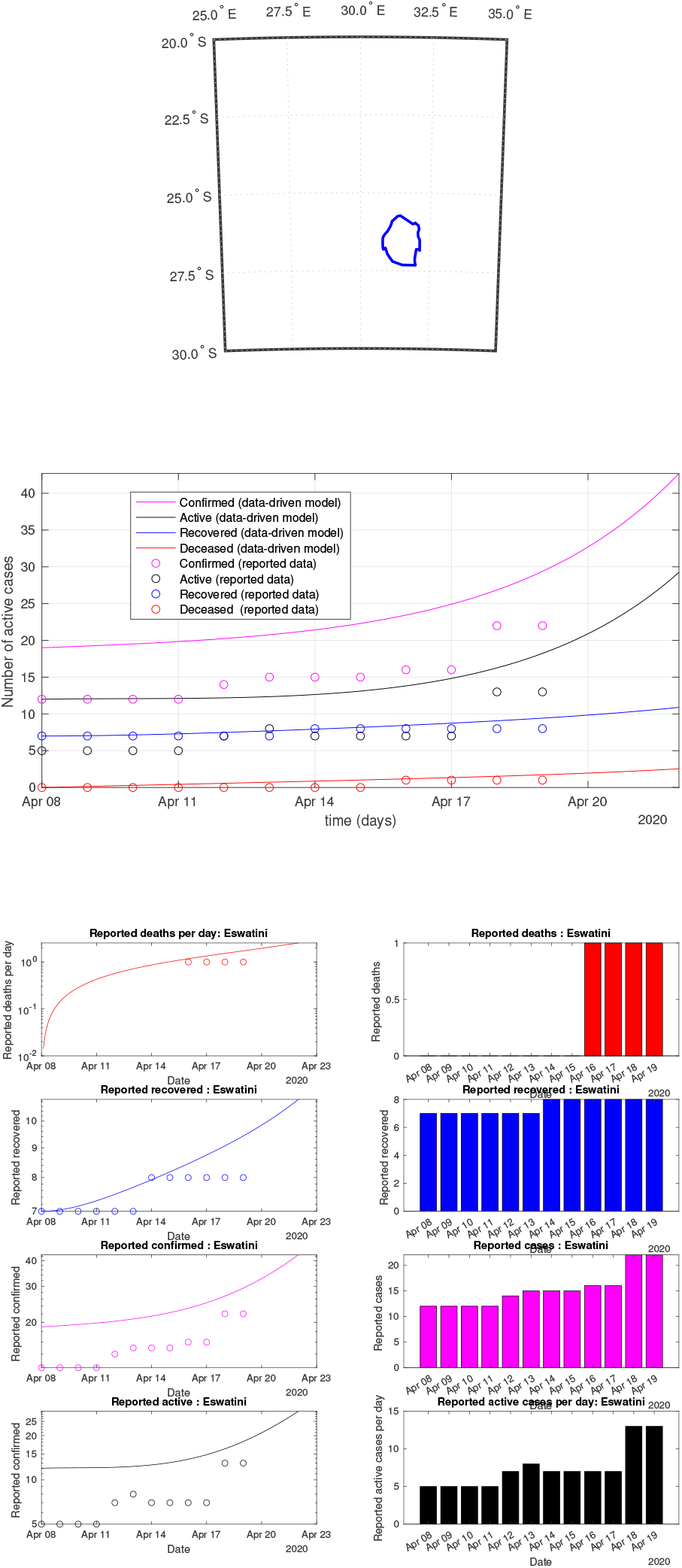
Eswatini: data vs model

**Fig 35:**
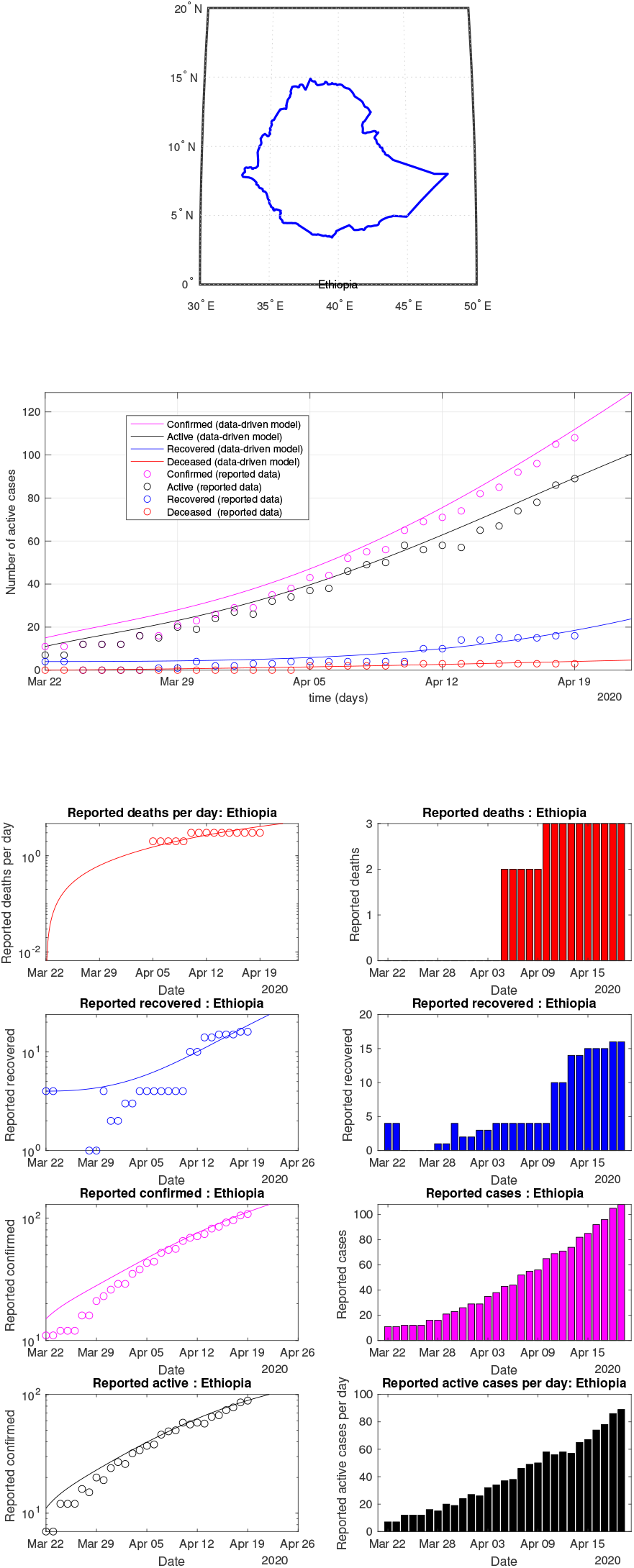
Ethiopia: data vs model

**Fig 36:**
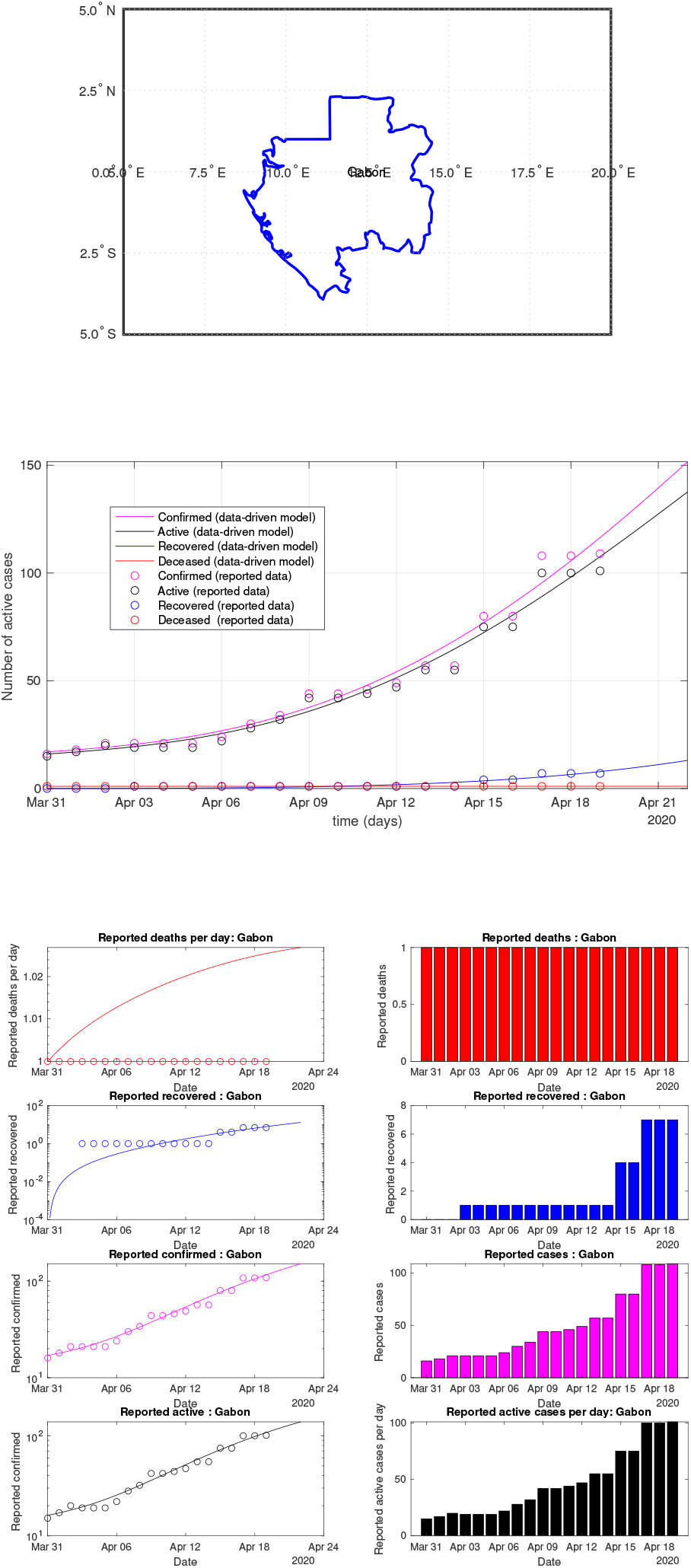
Gabon: data vs model

**Fig 37:**
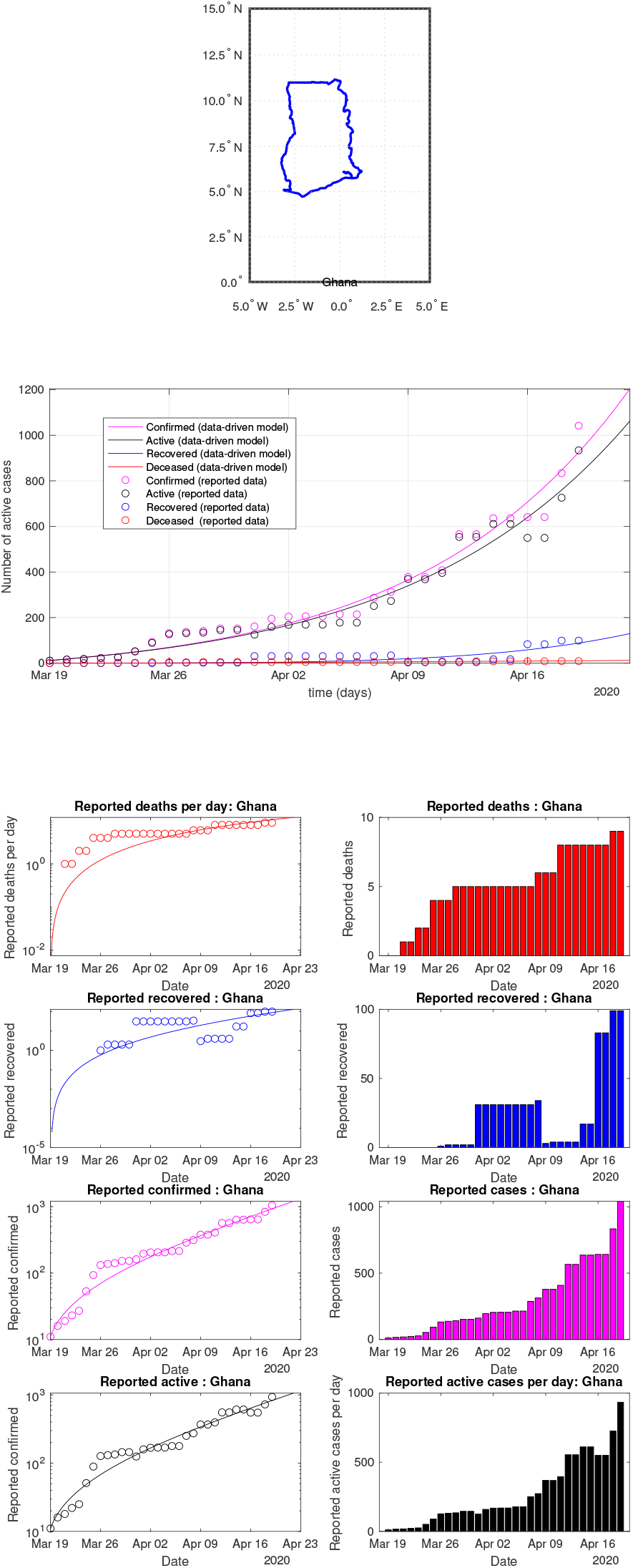
Ghana: data vs model

**Fig 38:**
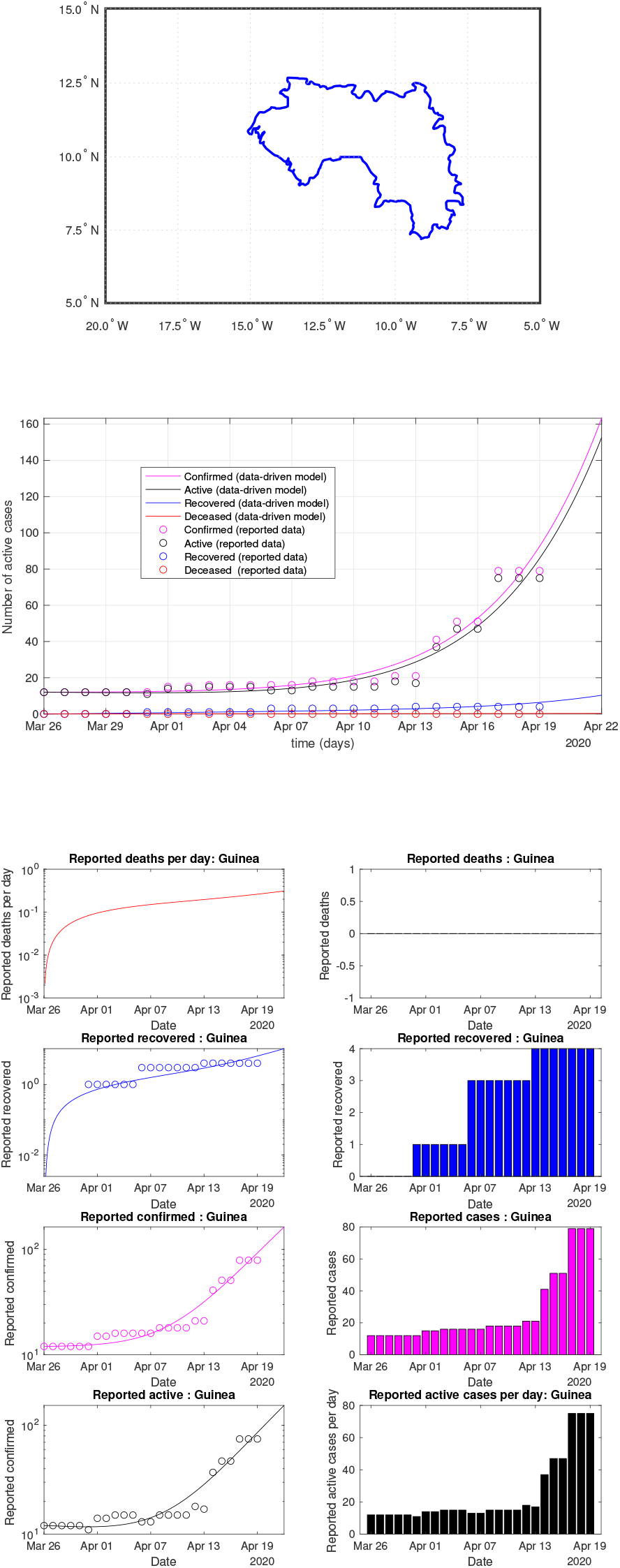
Guinea: data vs model

**Fig 39:**
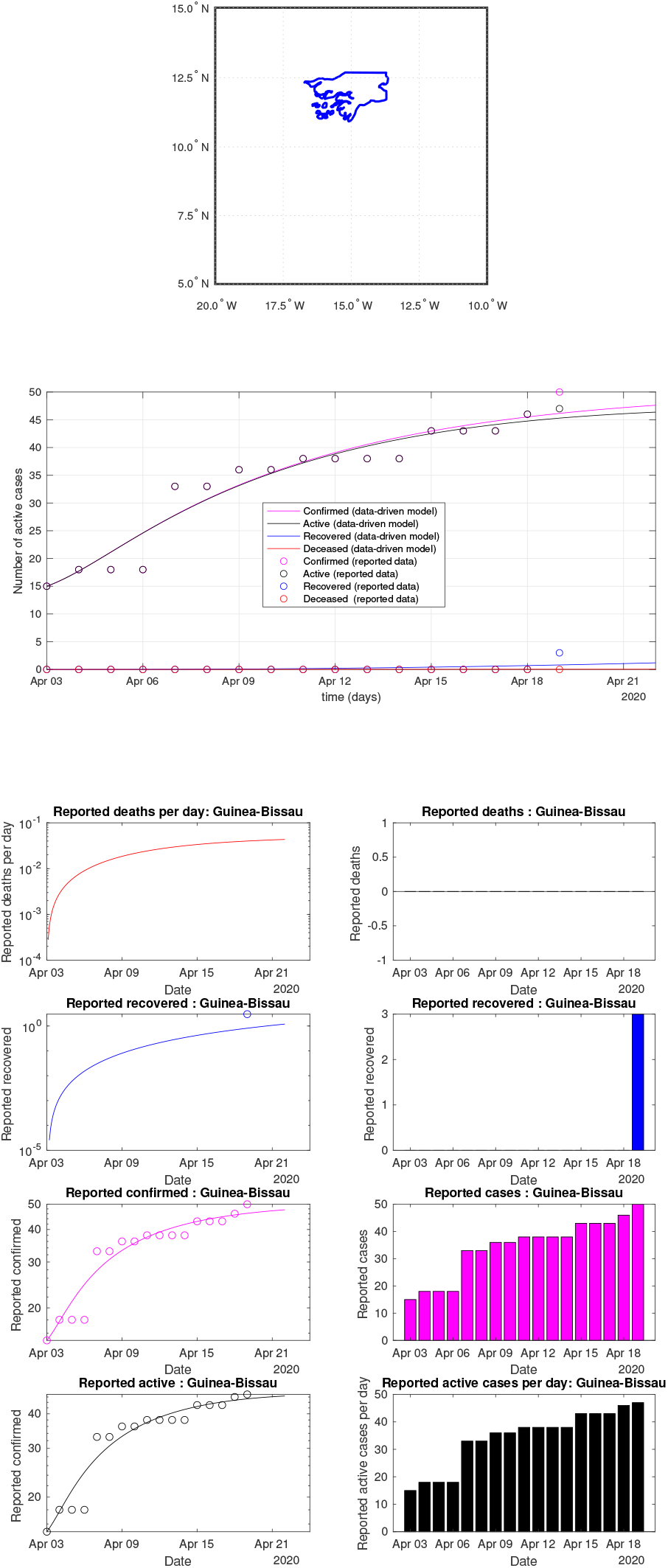
Guinea-Bissau: data vs model

**Fig 40:**
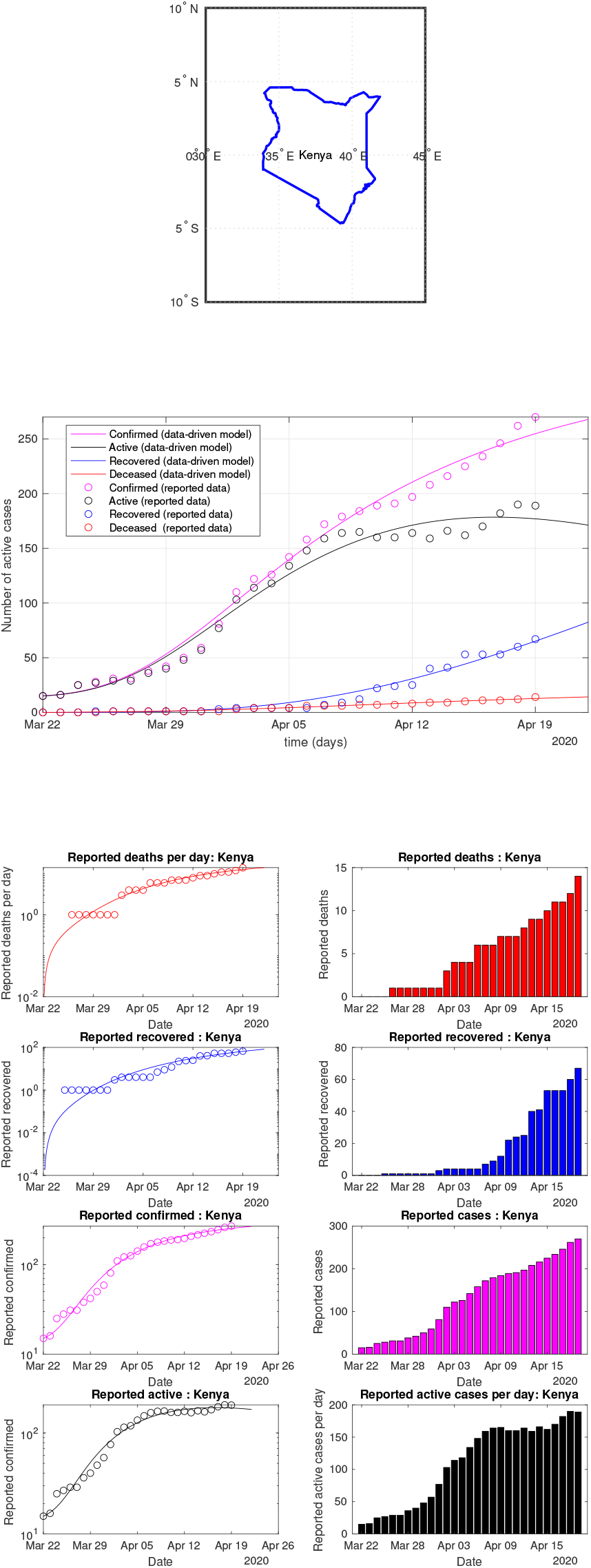
Kenya: data vs model

**Fig 41:**
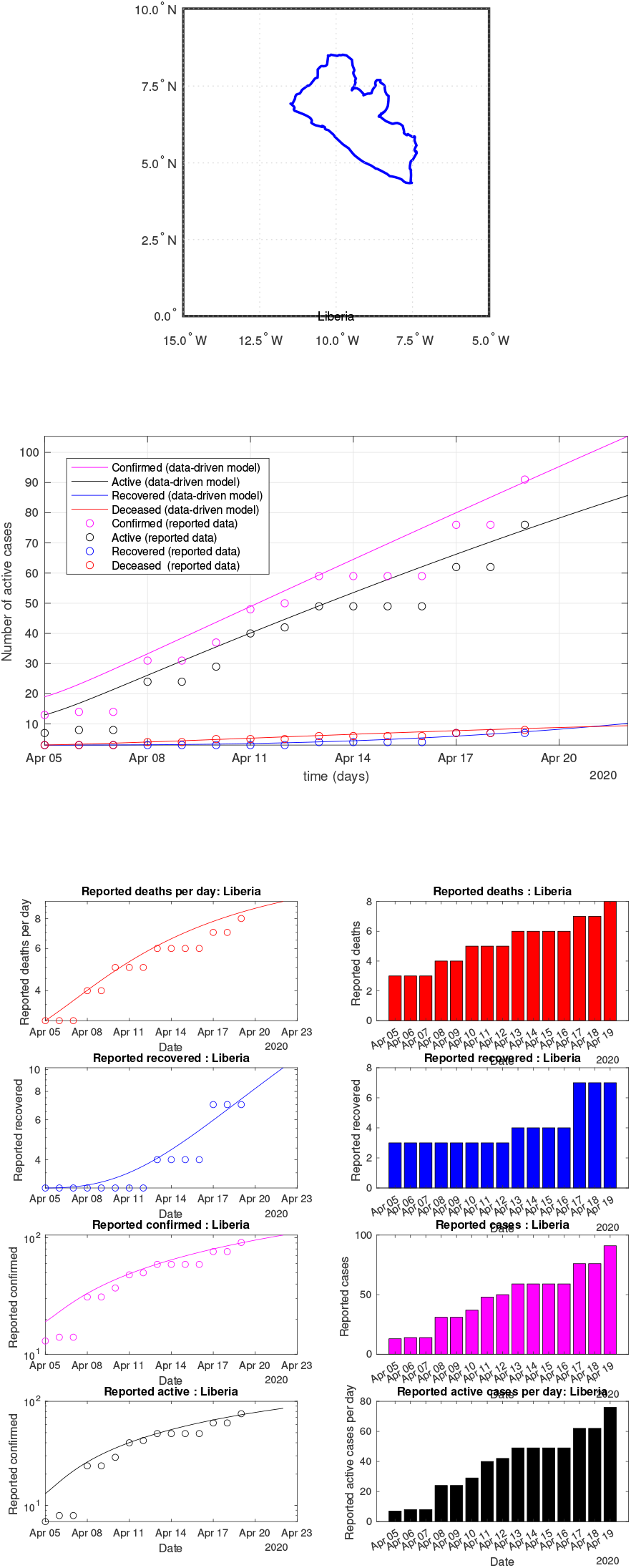
Liberia: data vs model

**Fig 42:**
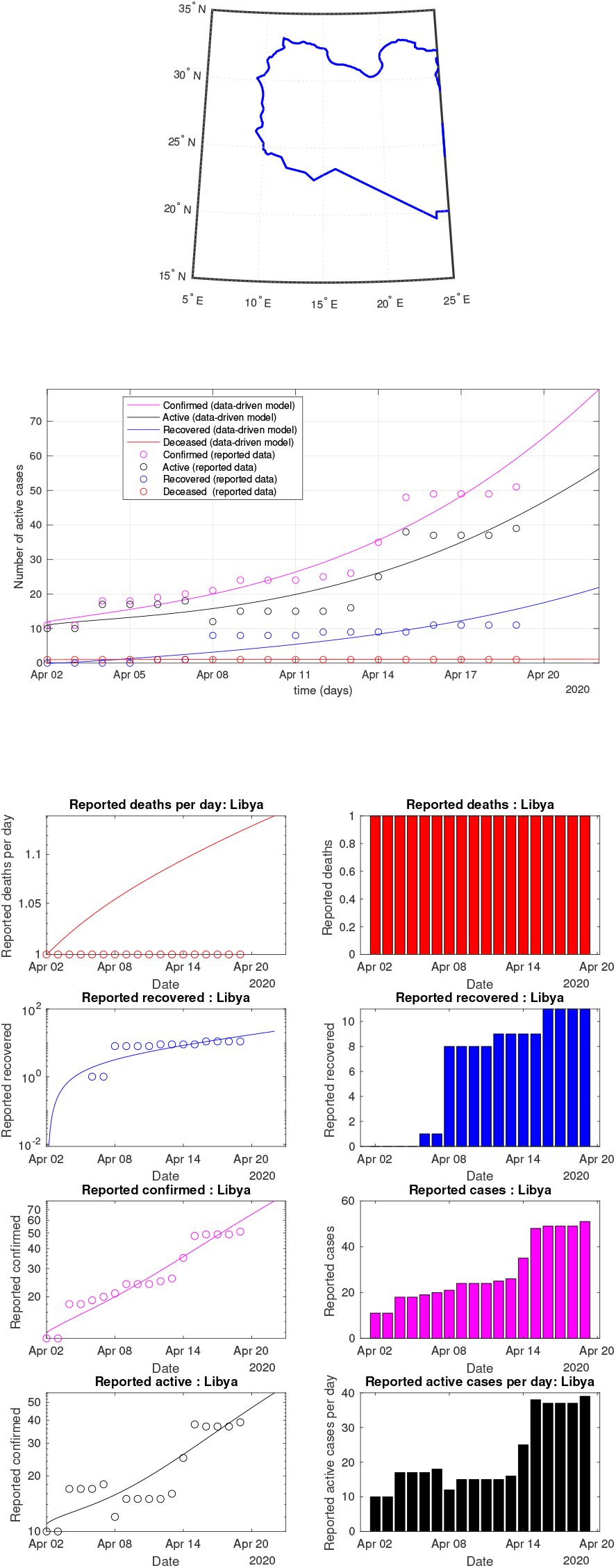
Libya: data vs model

**Fig 43:**
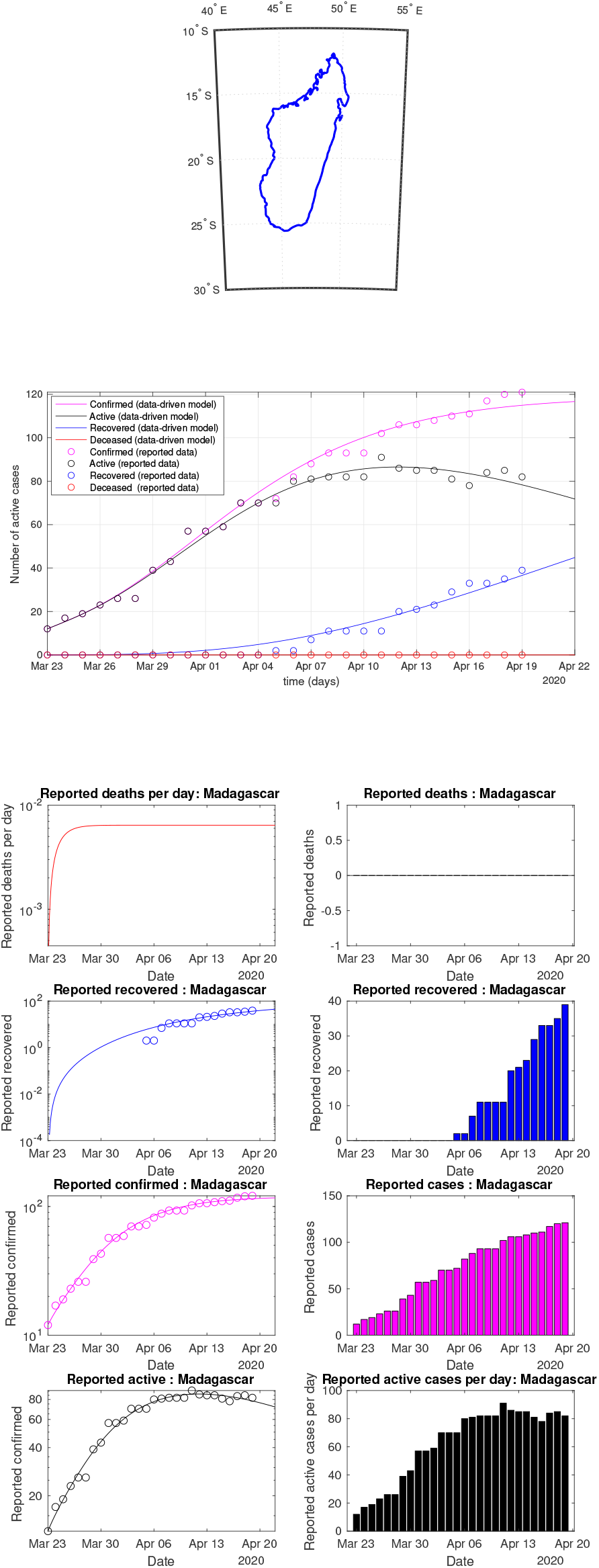
Madagascar: data vs model

**Fig 44:**
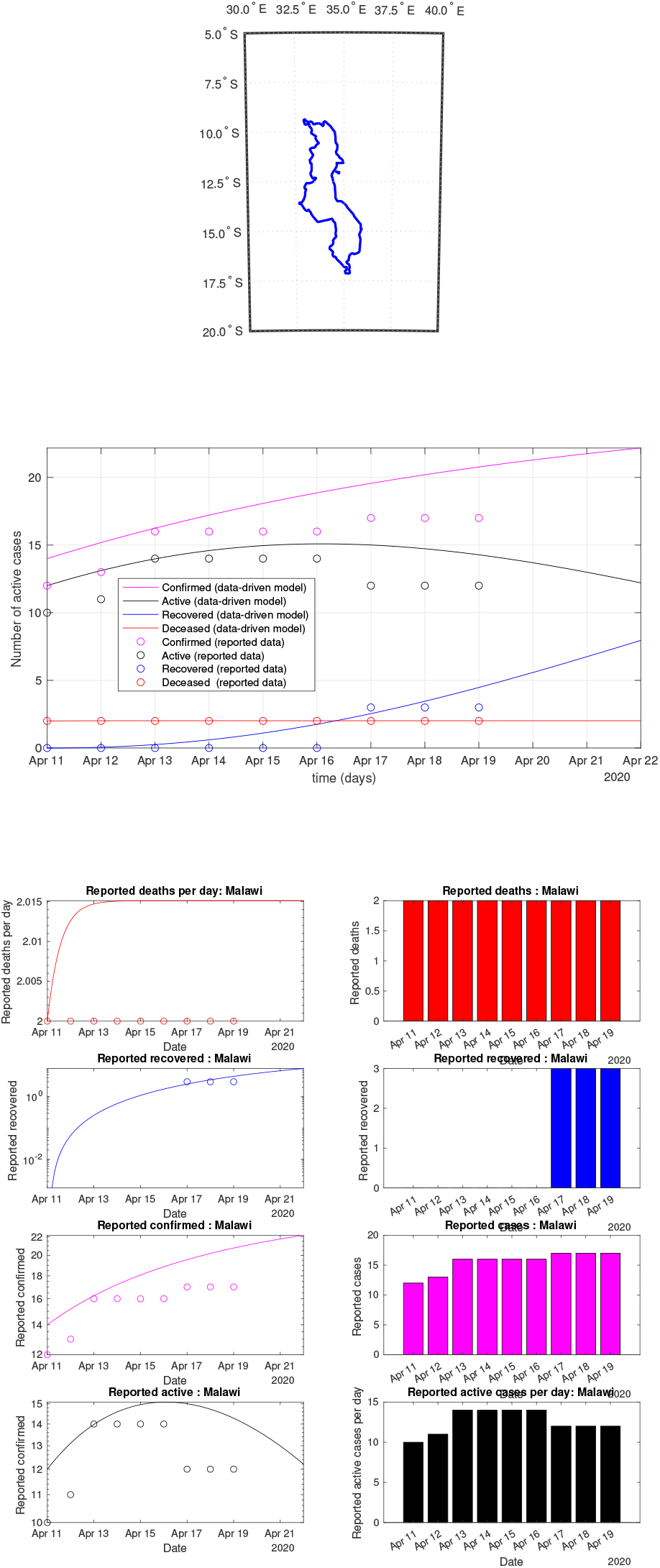
Malawi: data vs model

**Fig 45:**
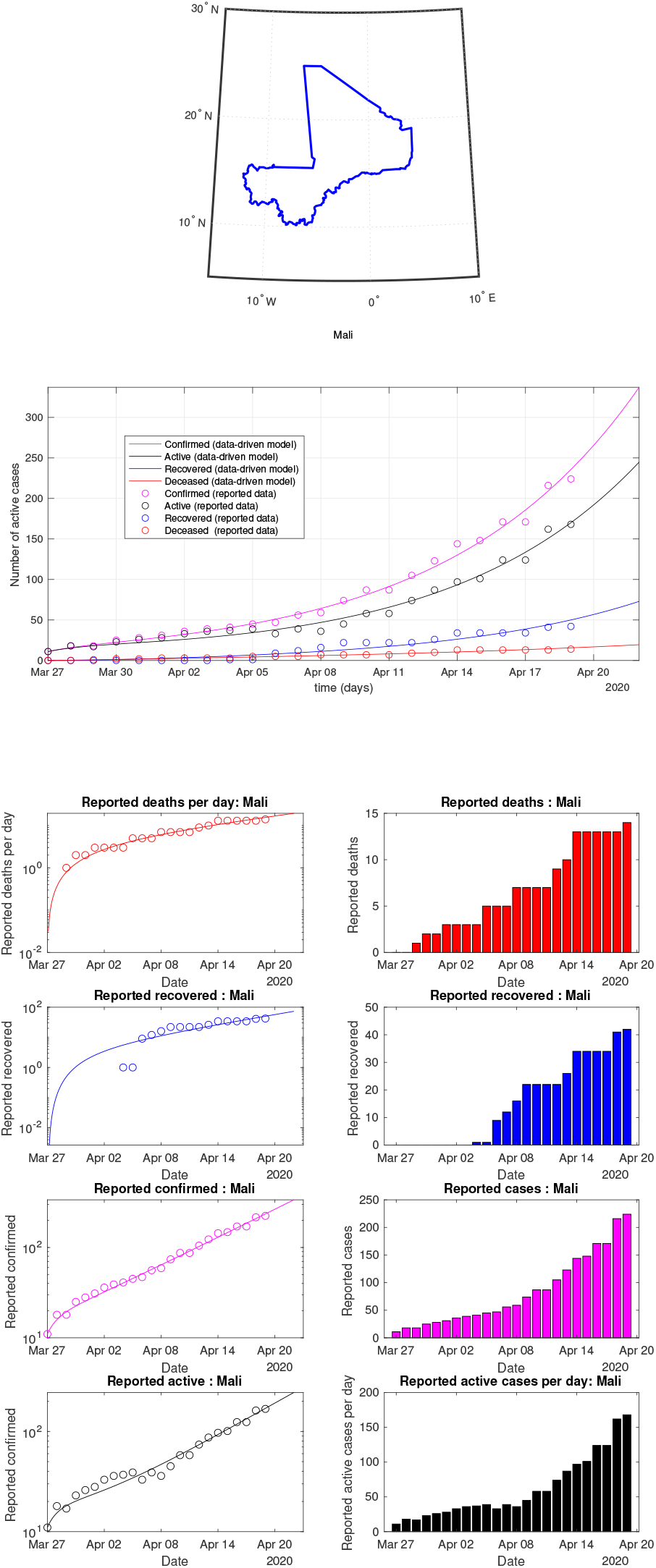
Mali: data vs model

**Fig 46:**
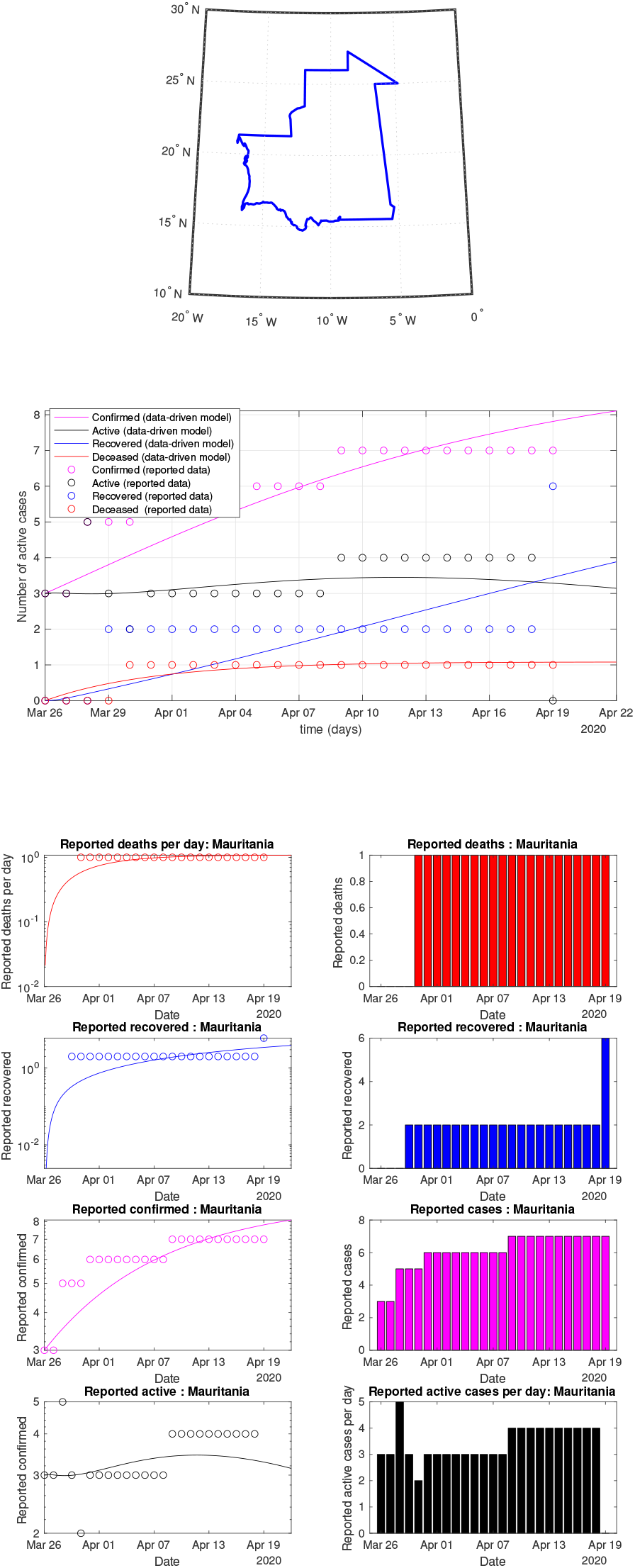
Mauritania: data vs model

**Fig 47:**
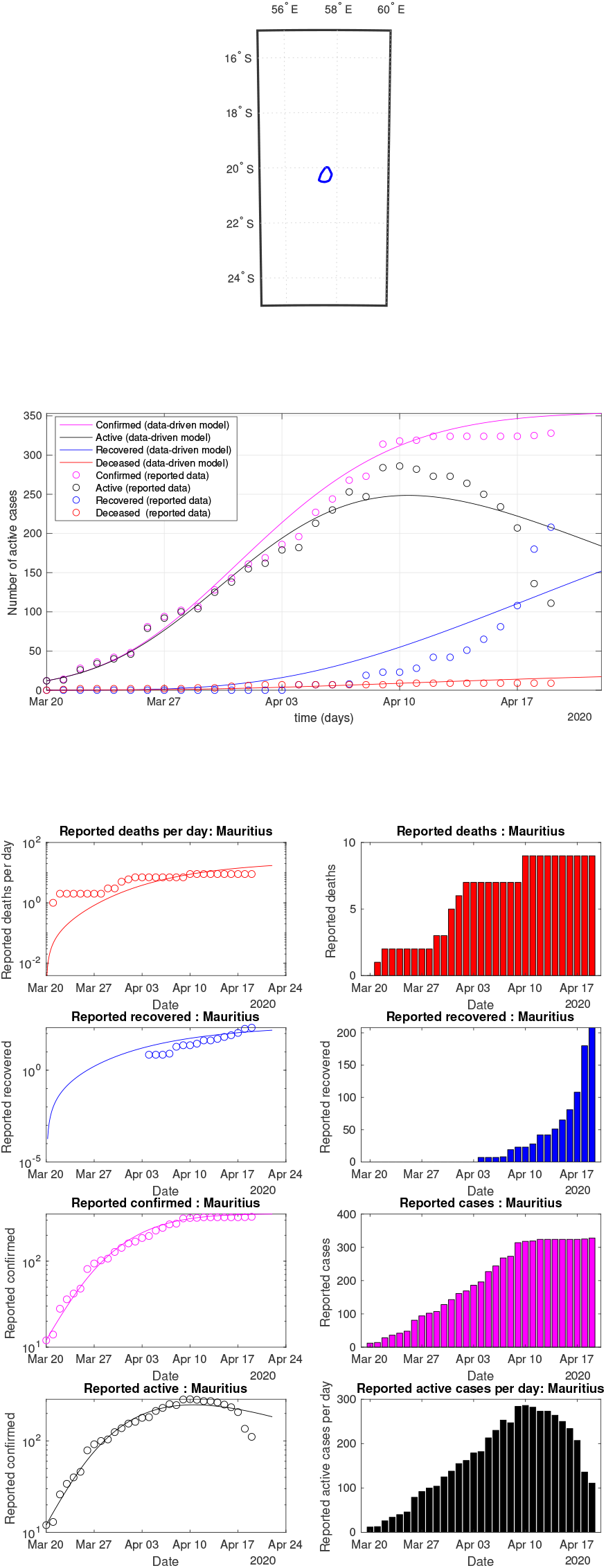
Mauritius: data vs model

**Fig 48:**
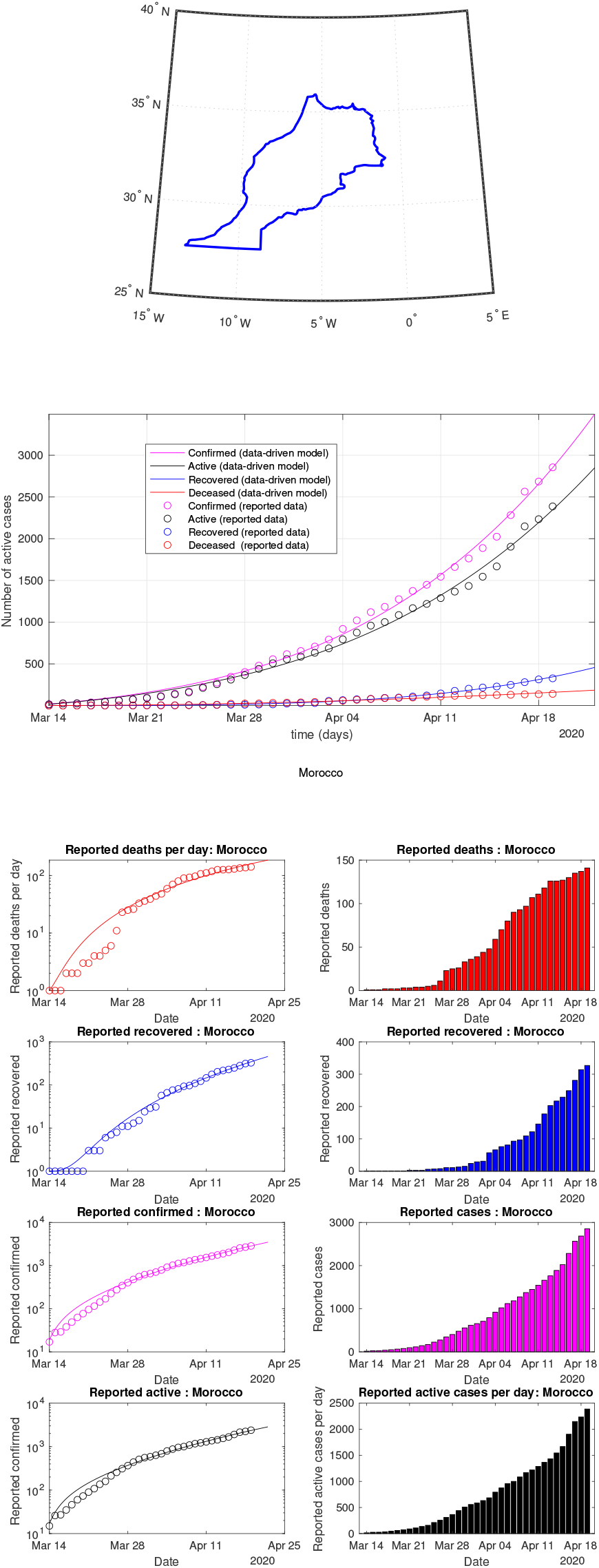
Morocco: data vs model

**Fig 49:**
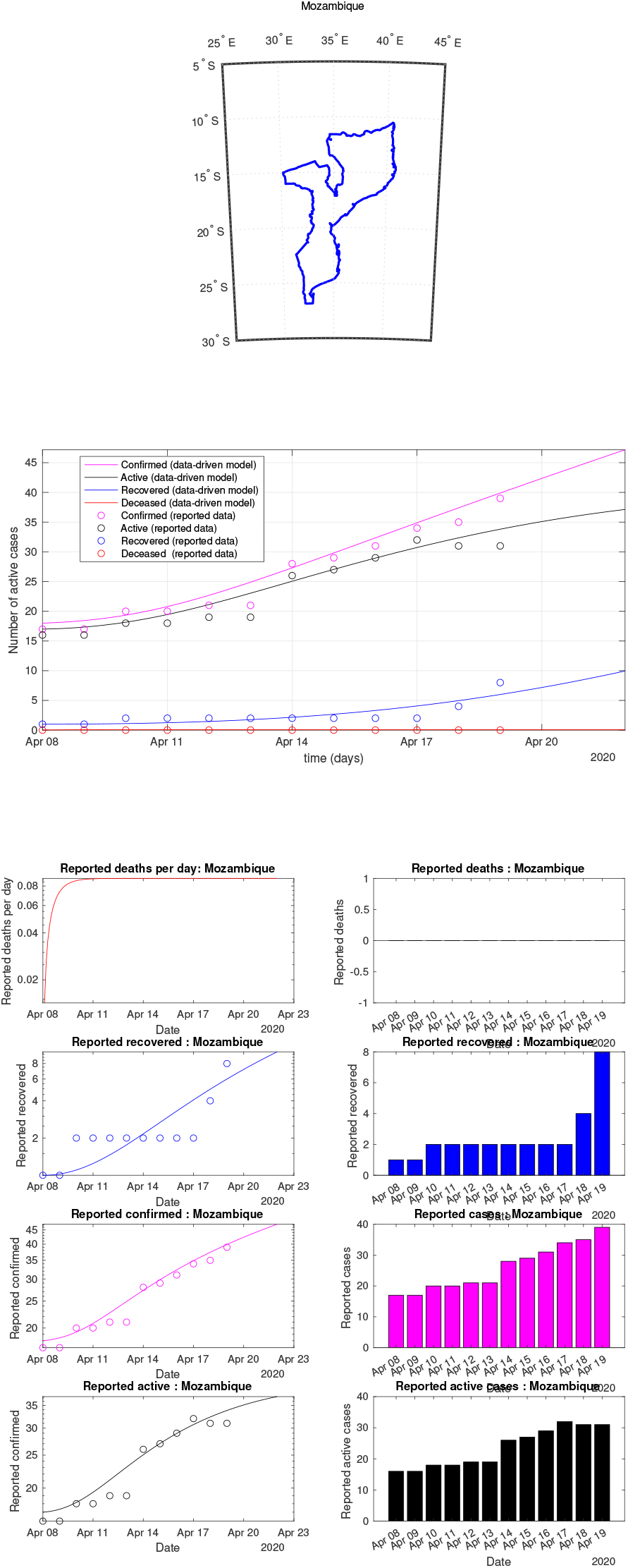
Mozambique: data vs model

**Fig 50:**
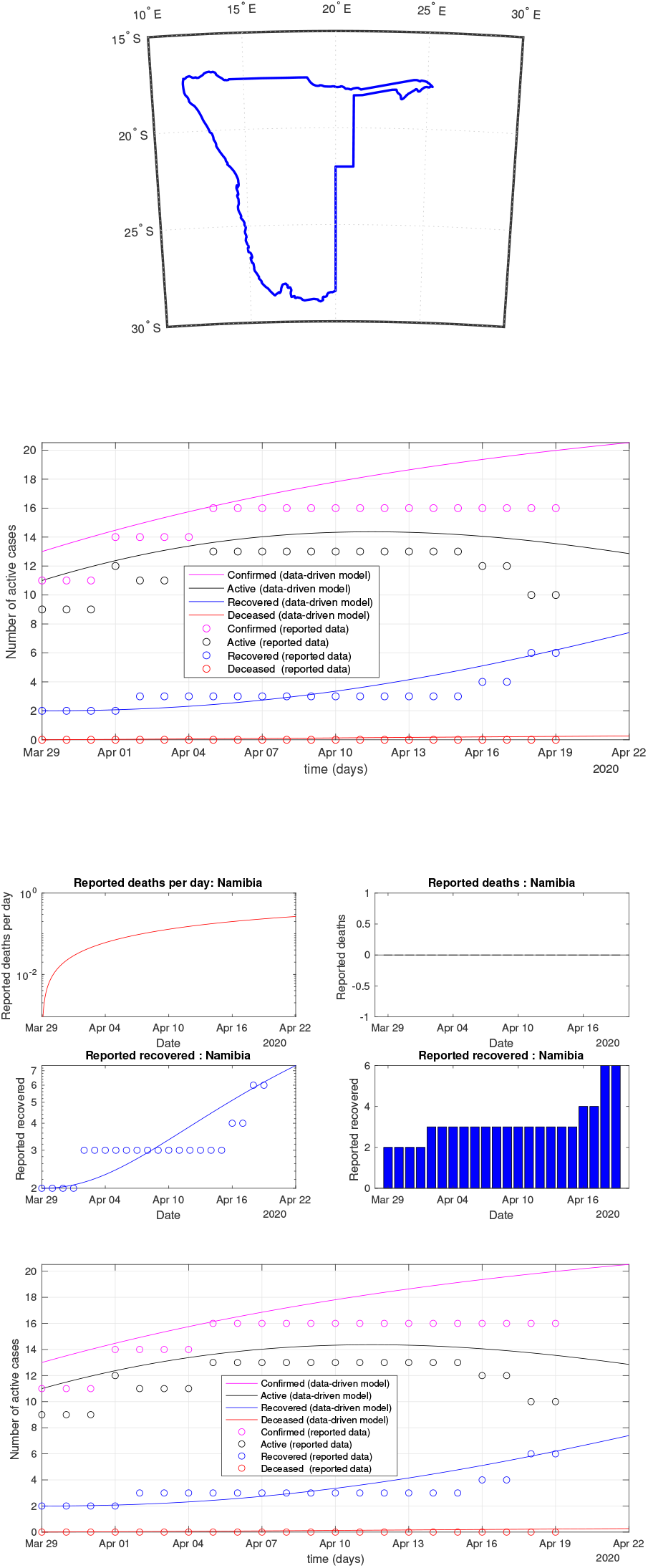
Namibia: data vs model

**Fig 51:**
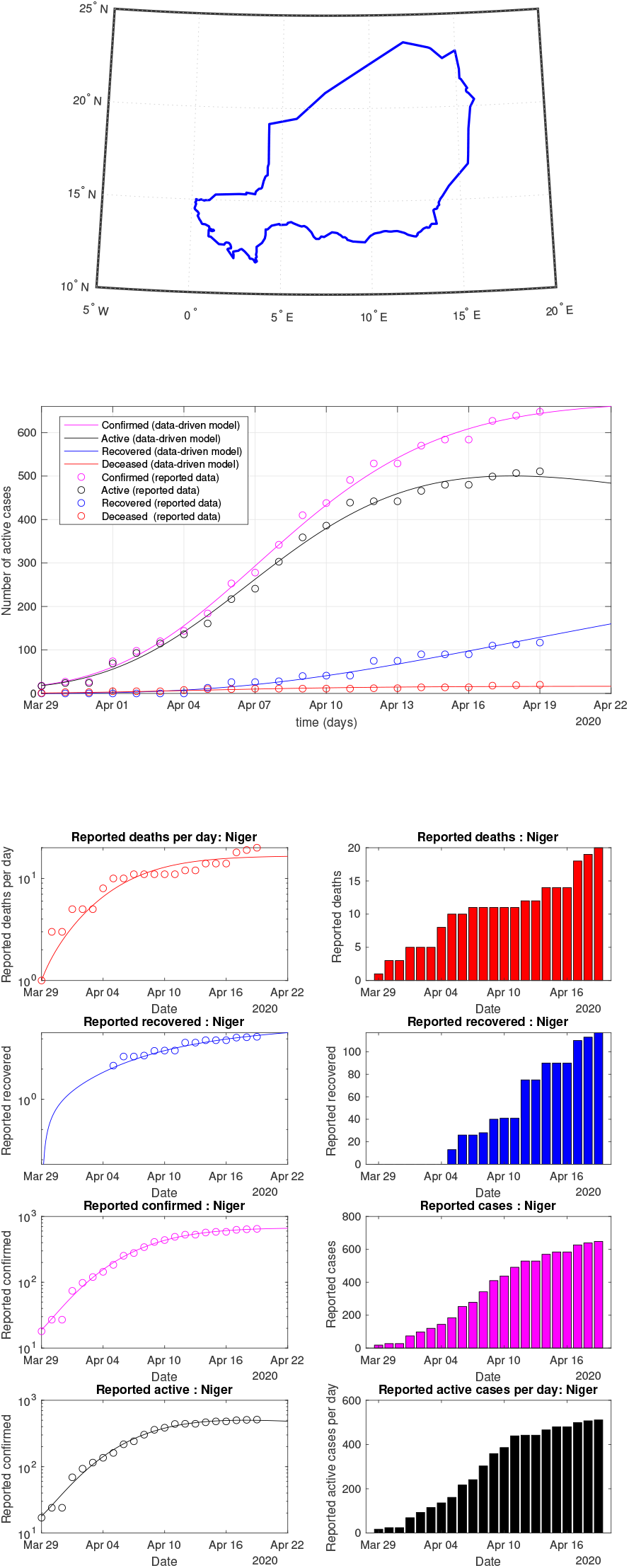
Niger: data vs model

**Fig 52:**
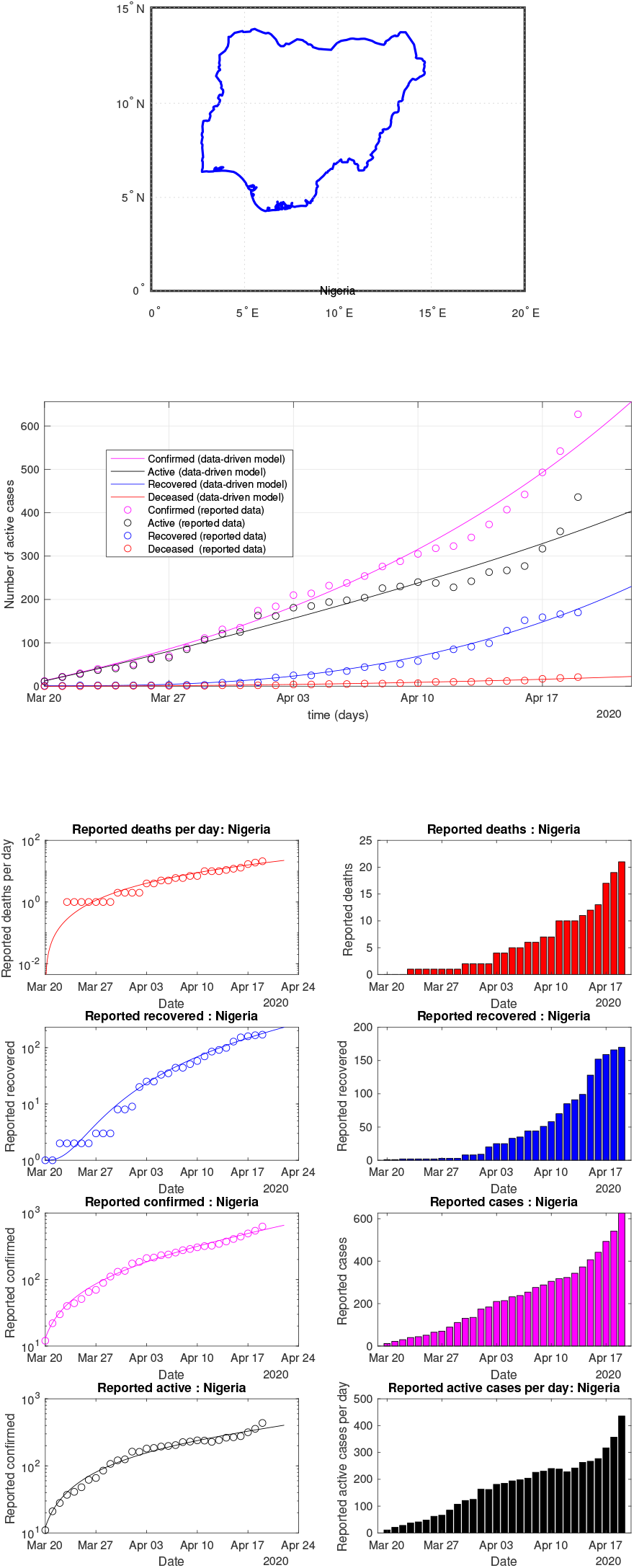
Nigeria: data vs model

**Fig 53:**
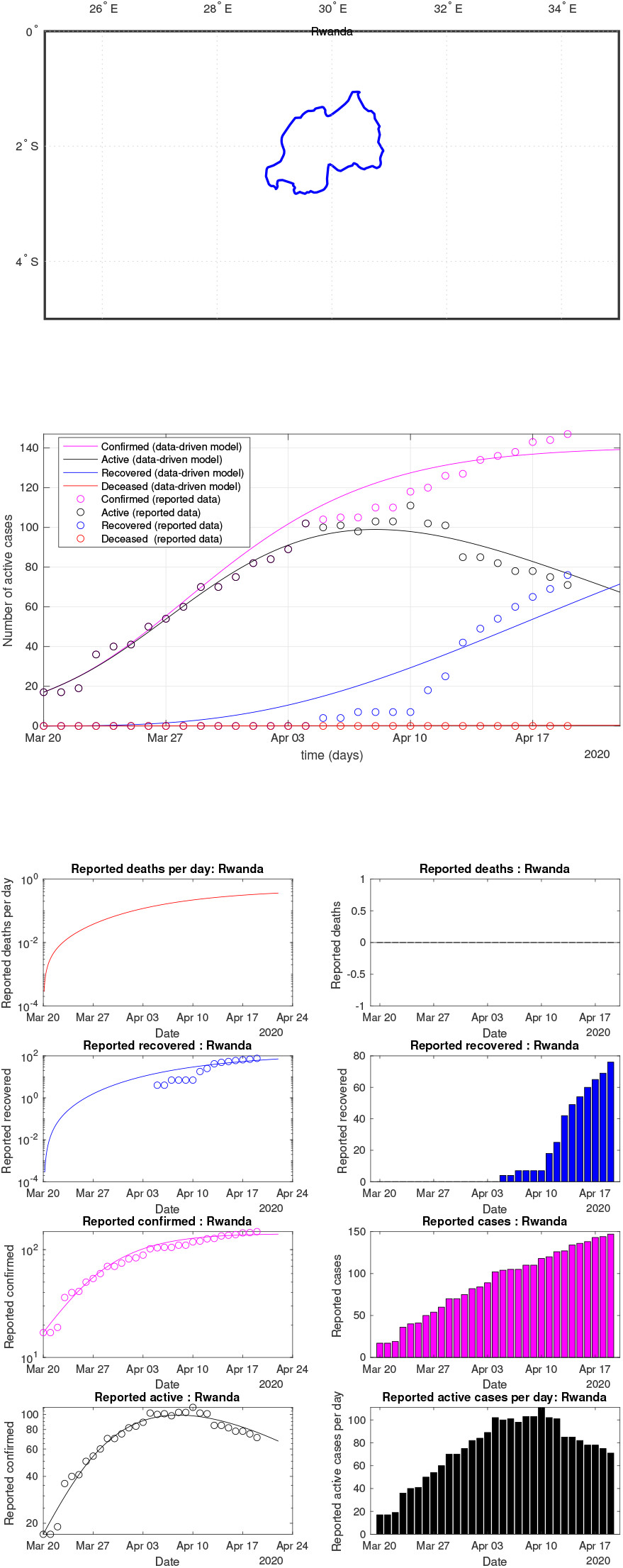
Rwanda: data vs model

**Fig 54:**
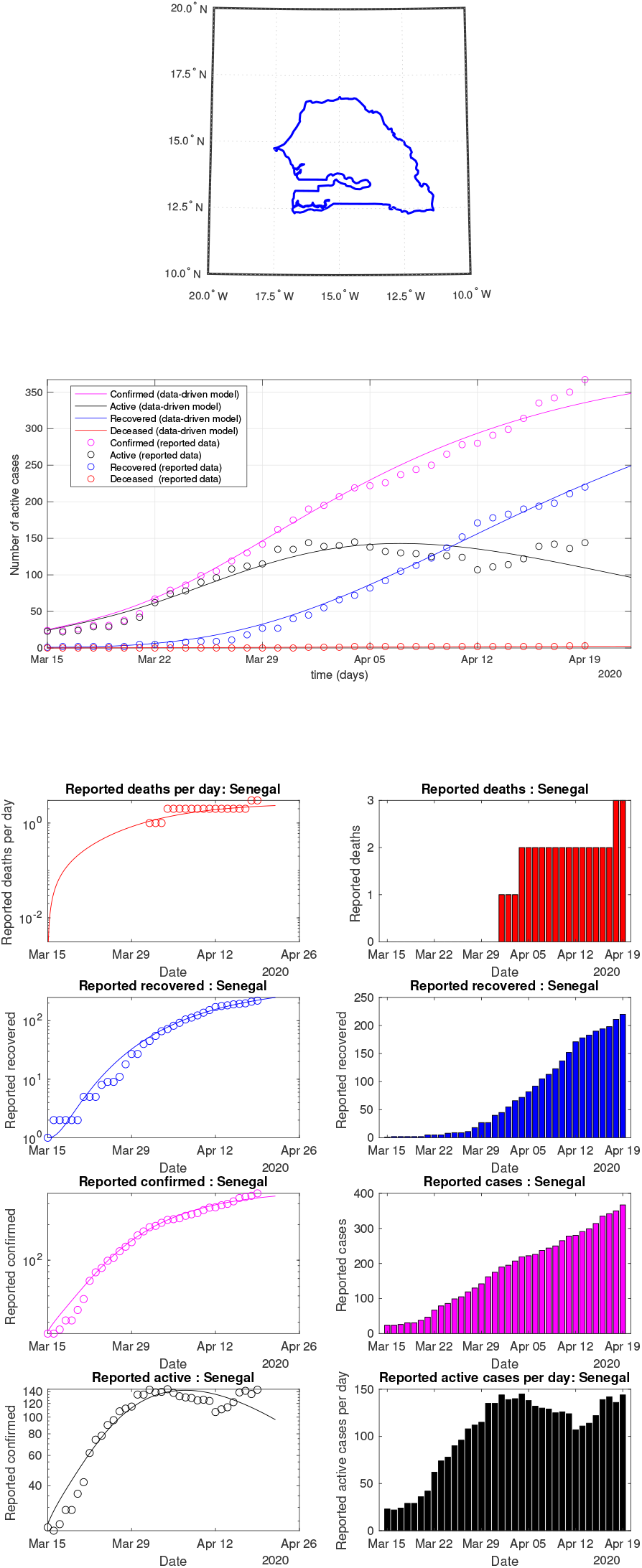
Senegal: data vs model

**Fig 55:**
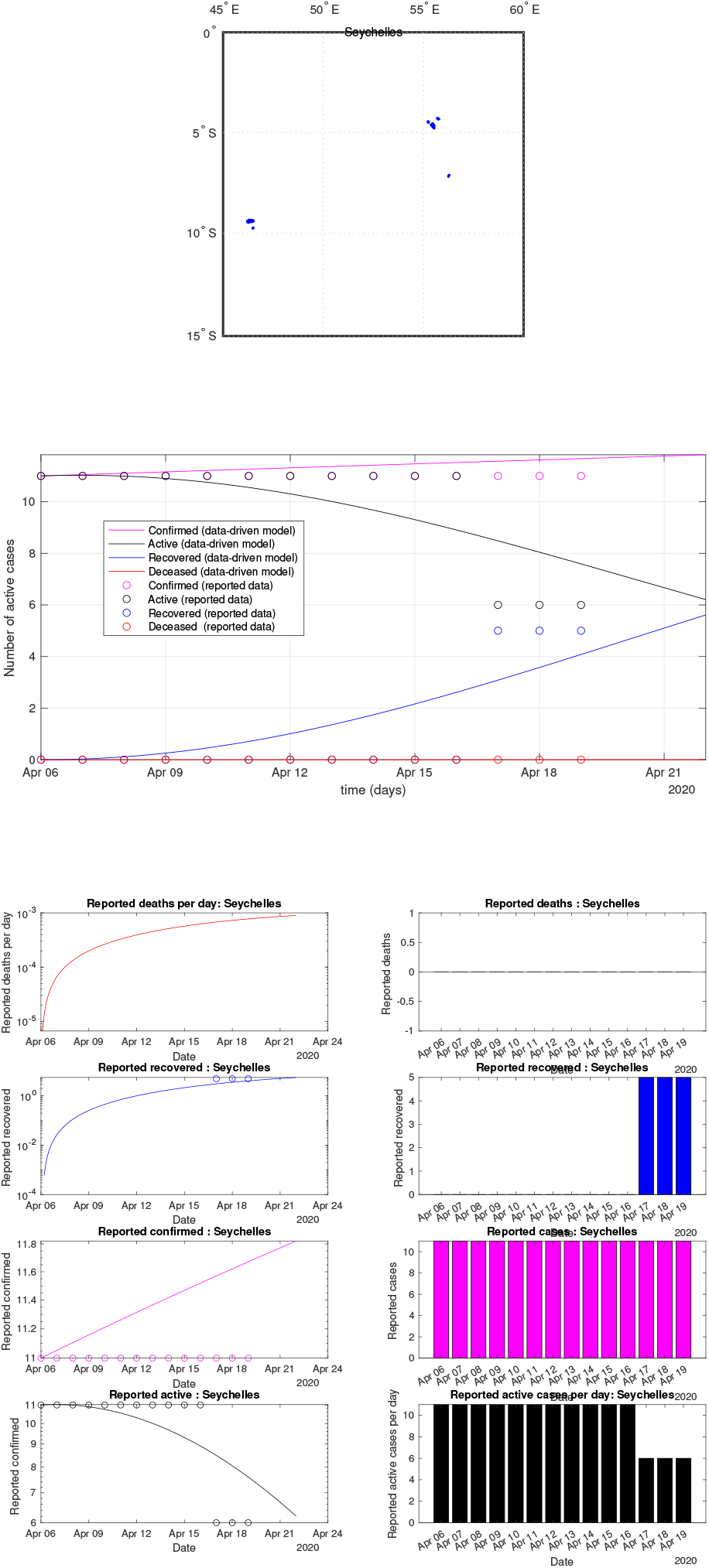
Seychelles: data vs model

**Fig 56:**
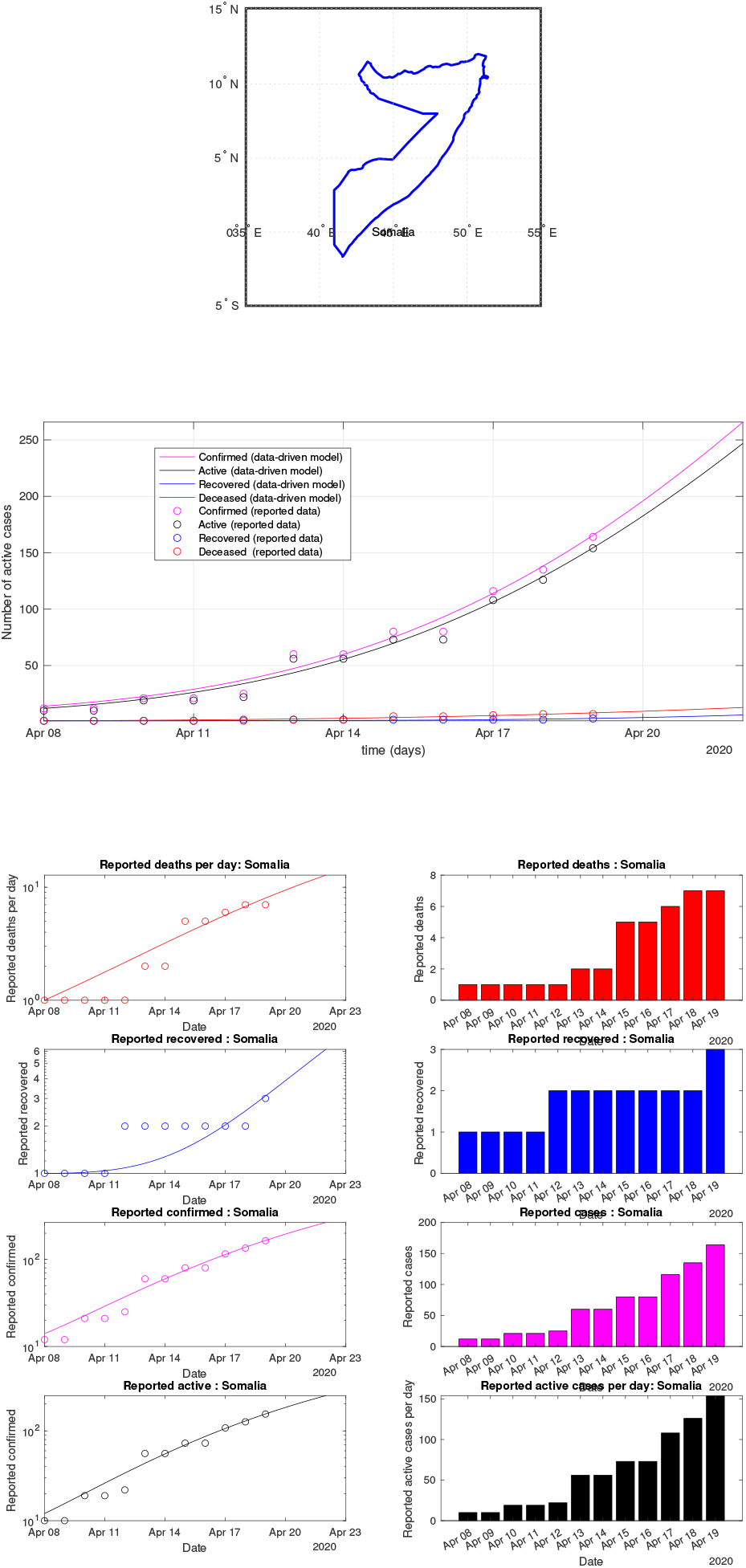
Somalia: data vs model

**Fig 57:**
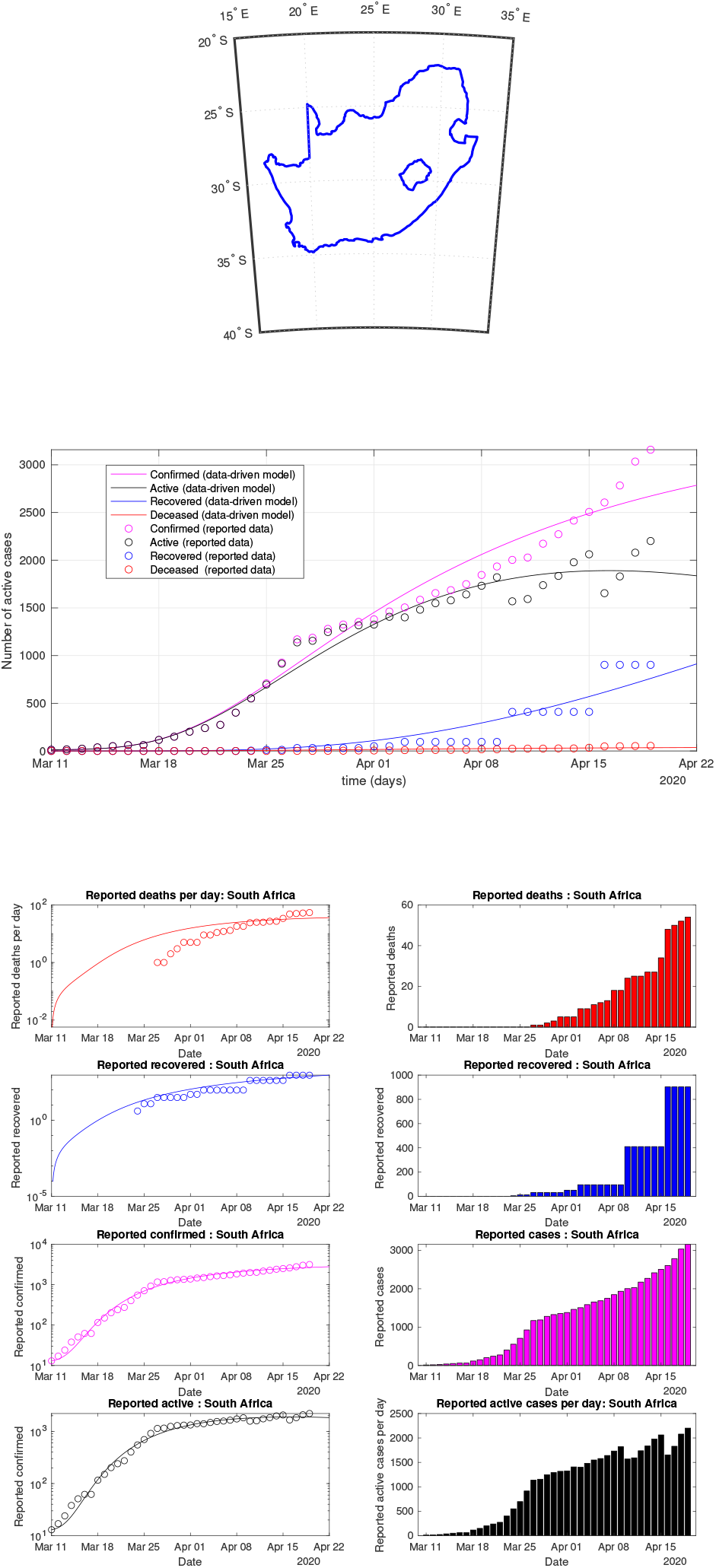
SouthAfrica: data vs model

**Fig 58:**
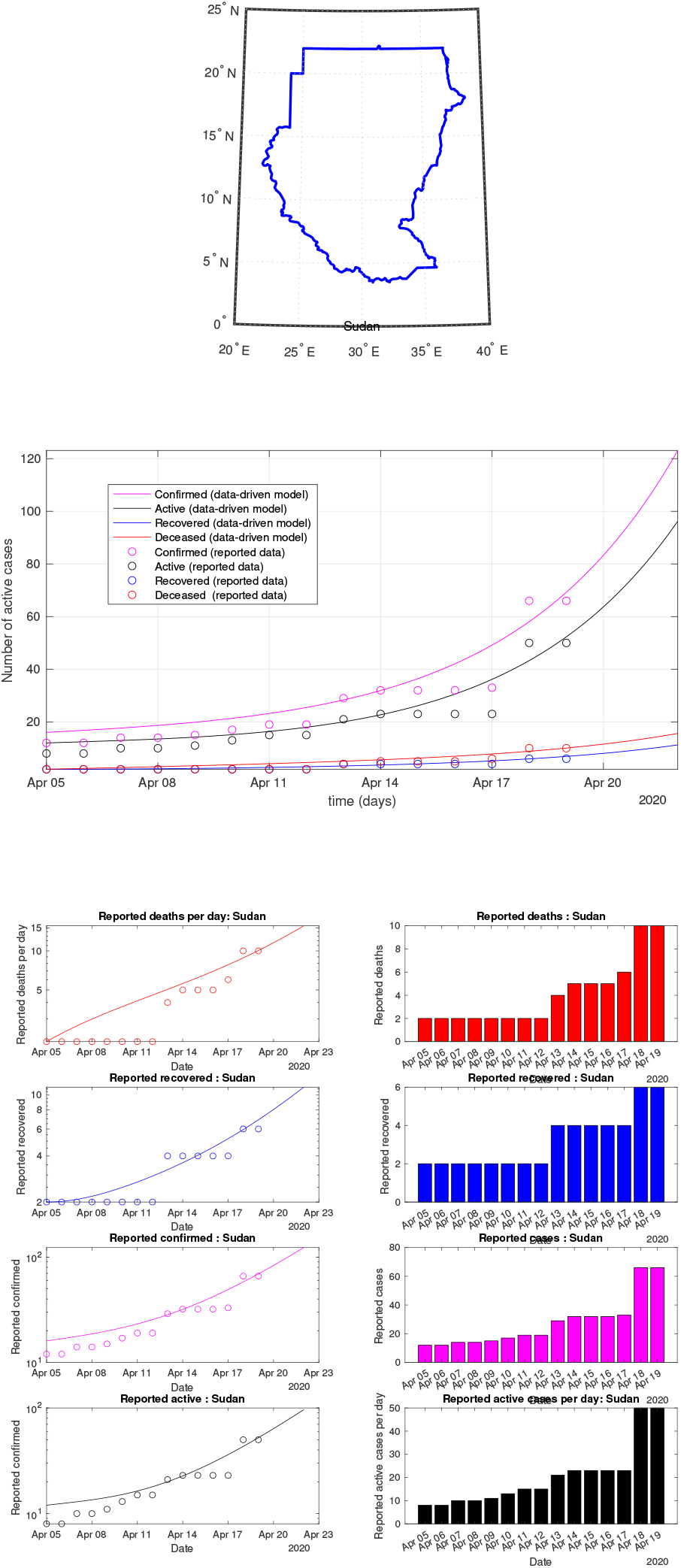
Sudan: data vs model

**Fig 59:**
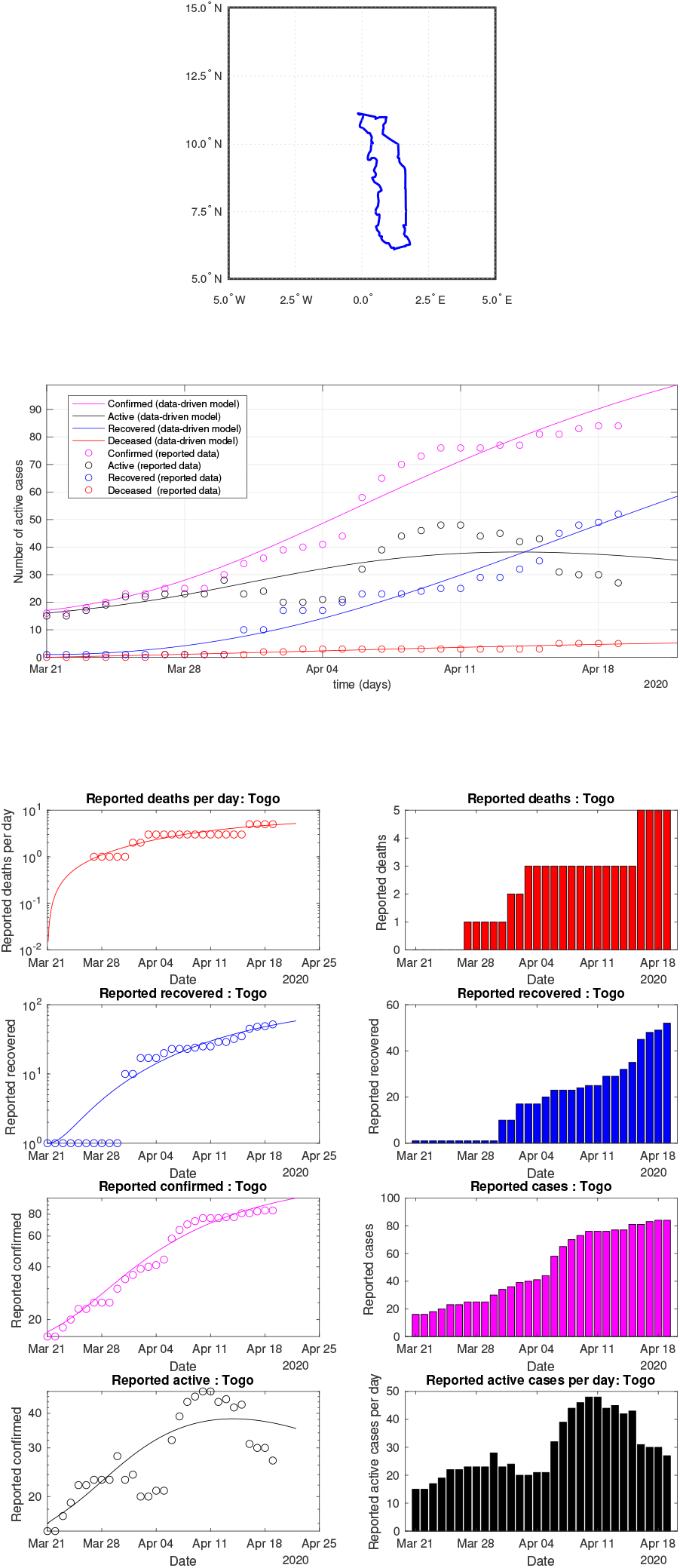
Togo: data vs model

**Fig 60:**
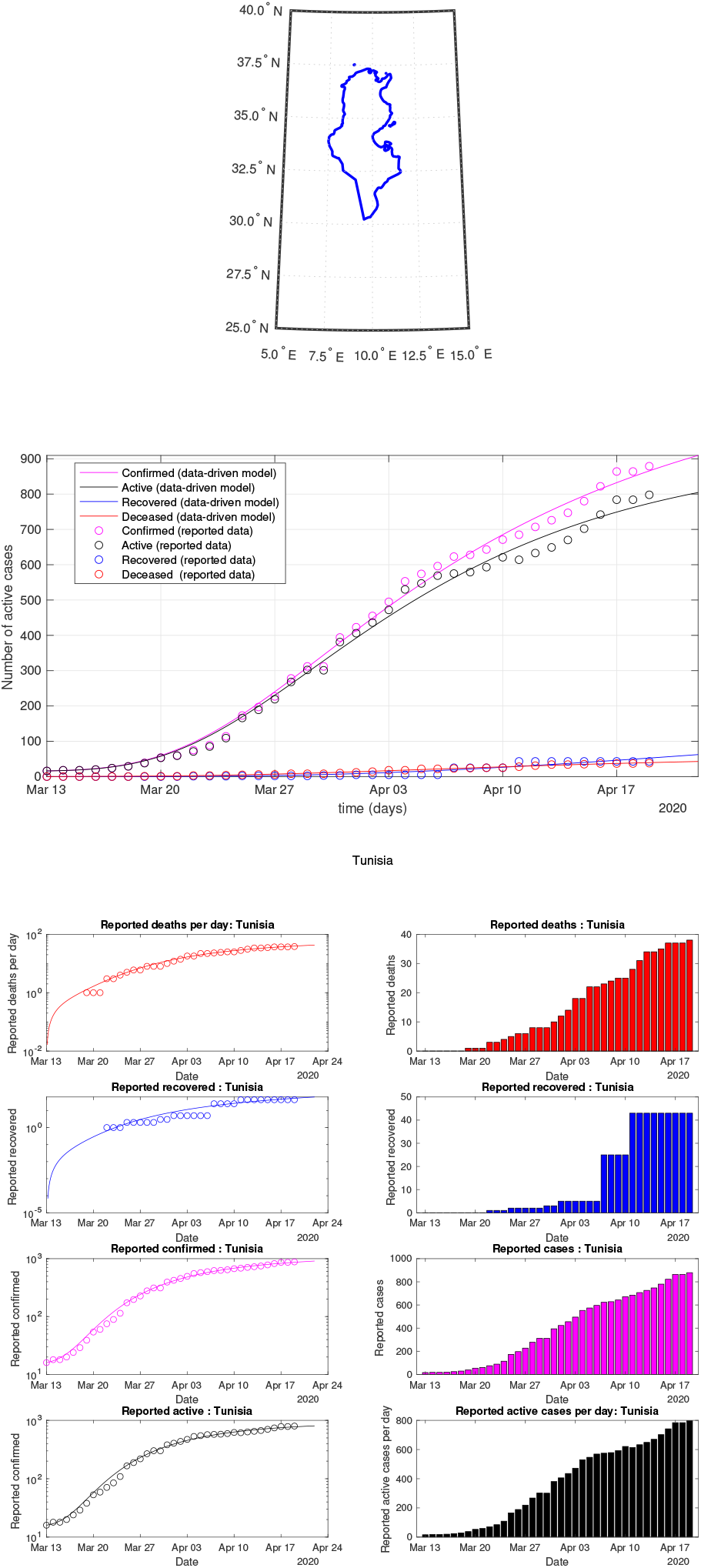
Tunisia: data vs model

**Fig 61:**
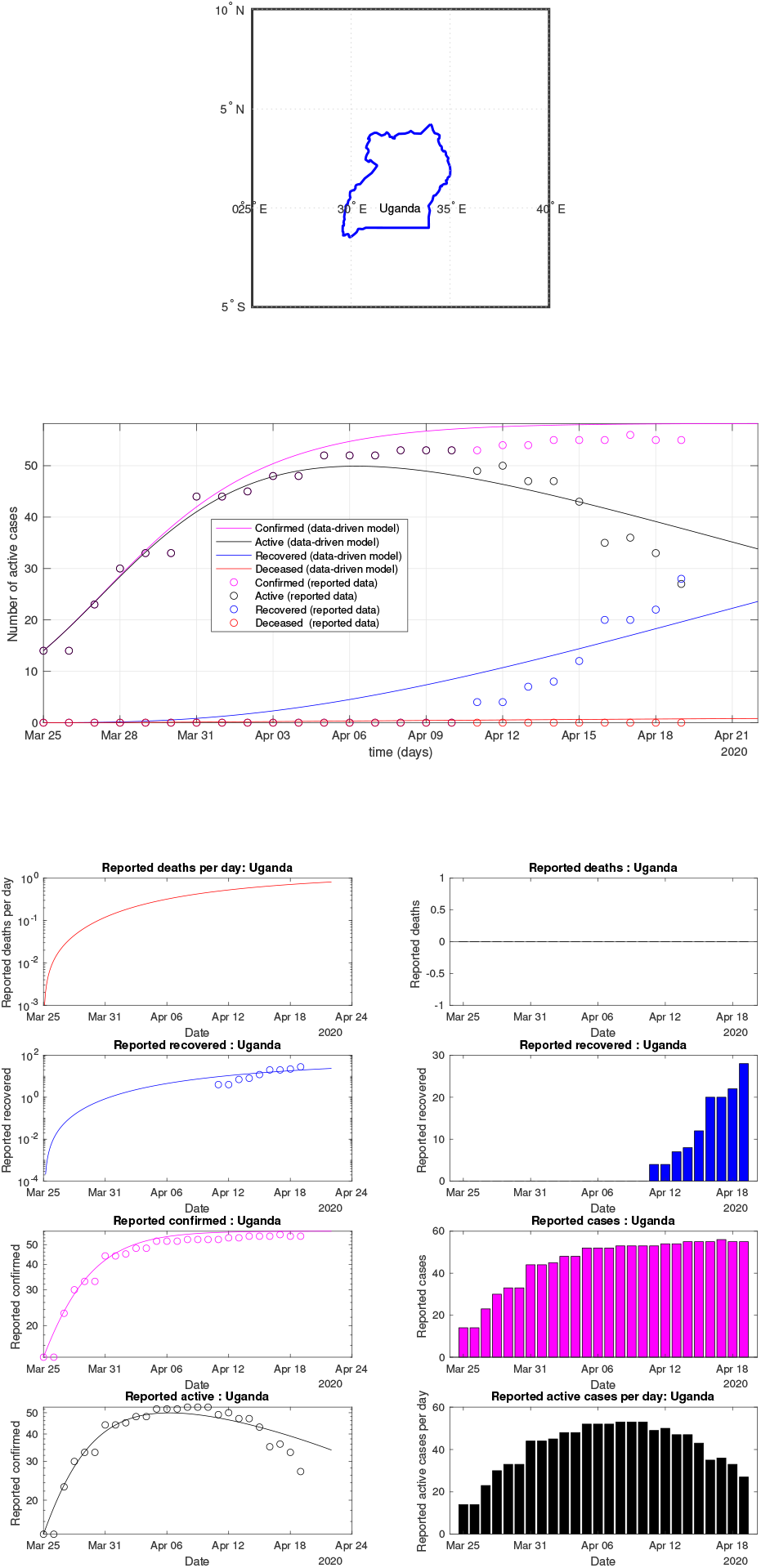
Uganda: data vs model

**Fig 62:**
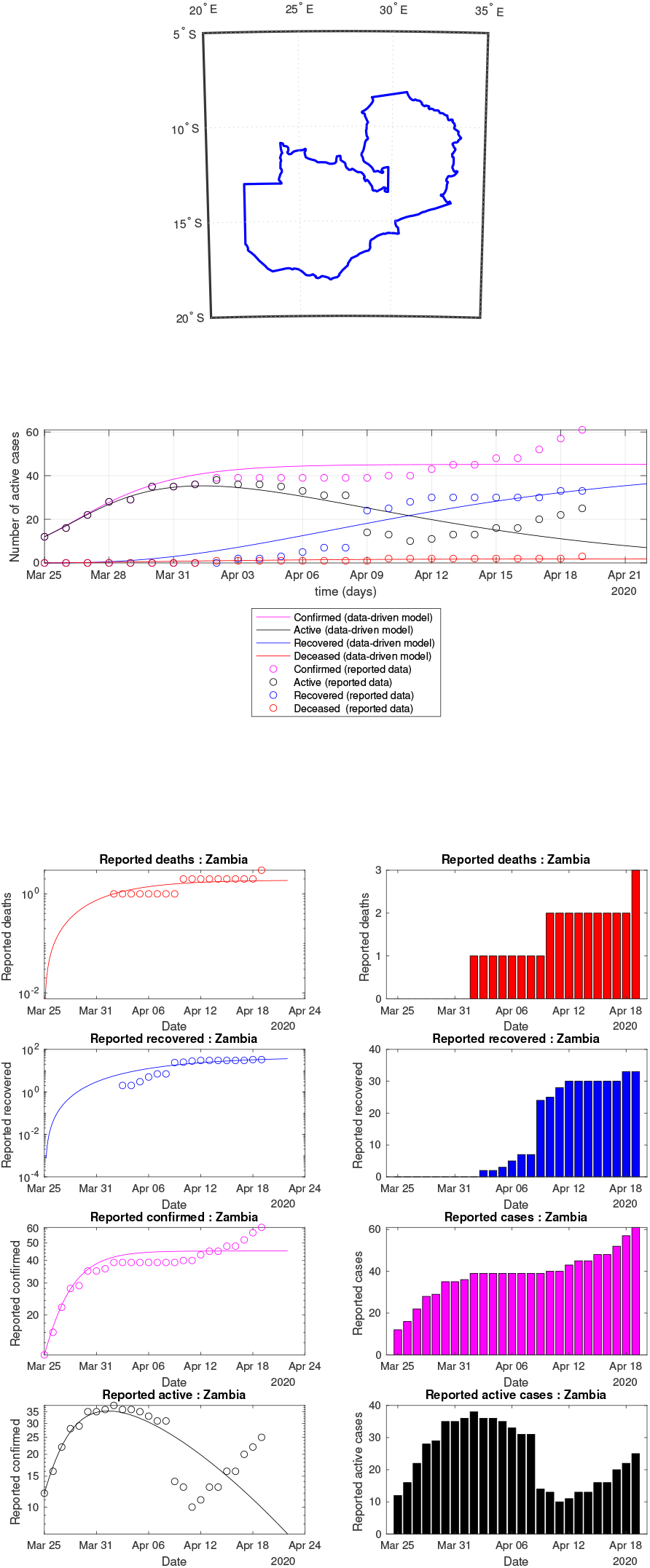
Zambia: data vs model

**Fig 63:**
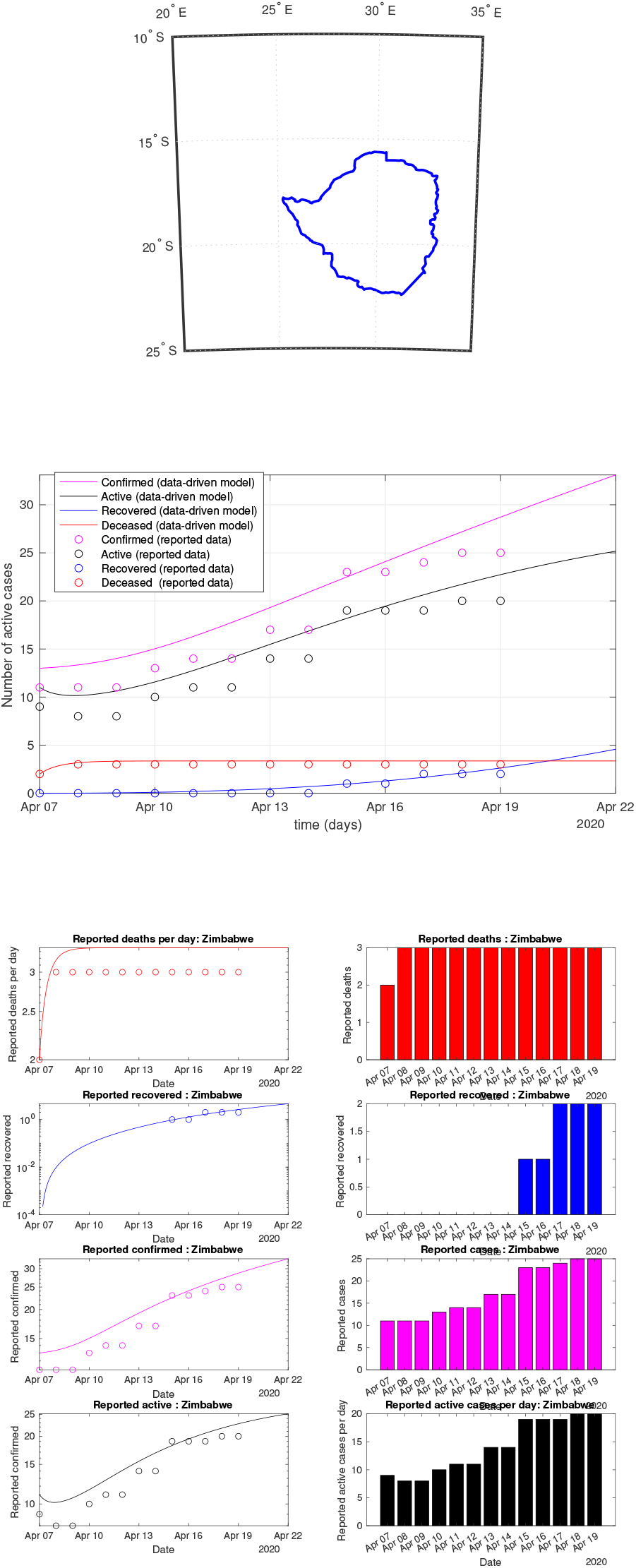
Zimbabwe: data vs model

**Fig 64:**
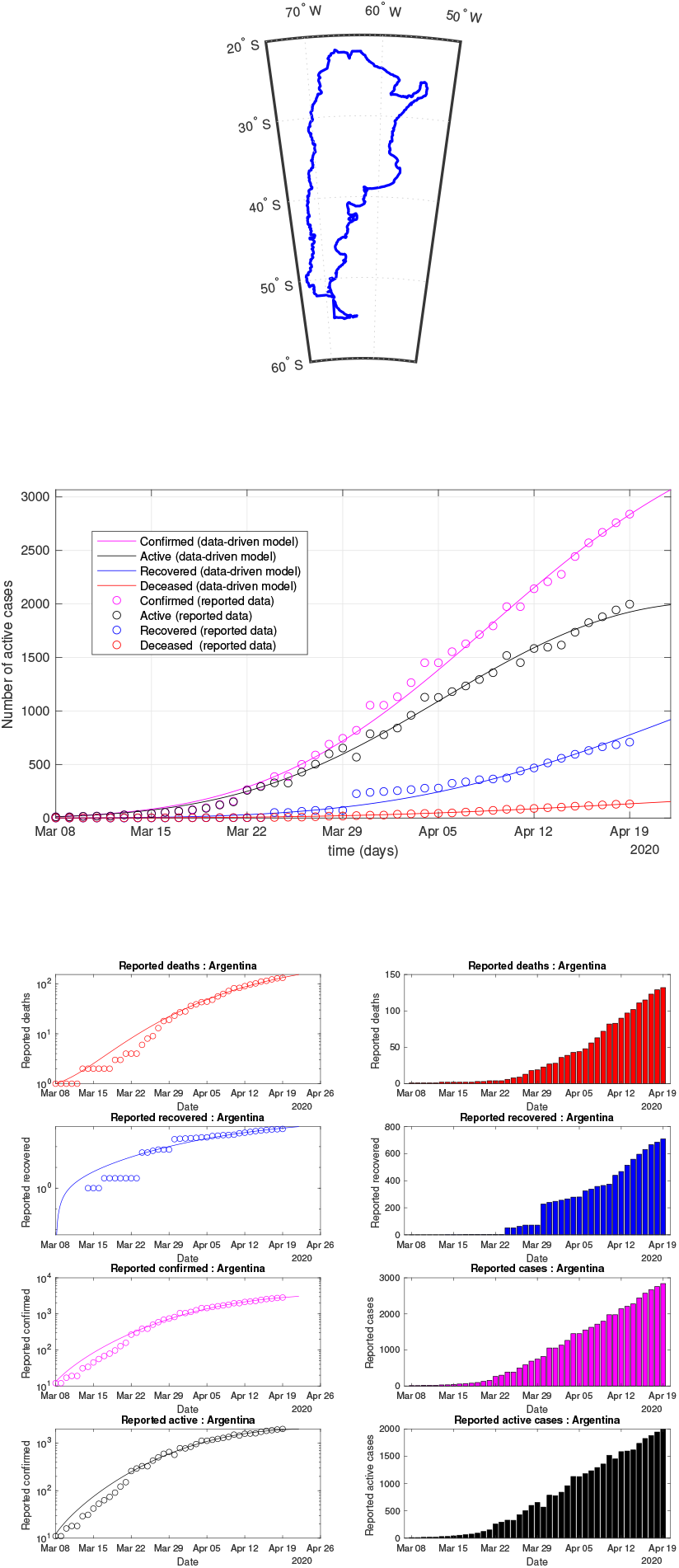
Argentina: data of active, recovered, deceased (in circle or bar) vs model of active, recovered, deceased over time. Tracking the number of active cases. Predictive analytics for the next couple days

**Fig 65:**
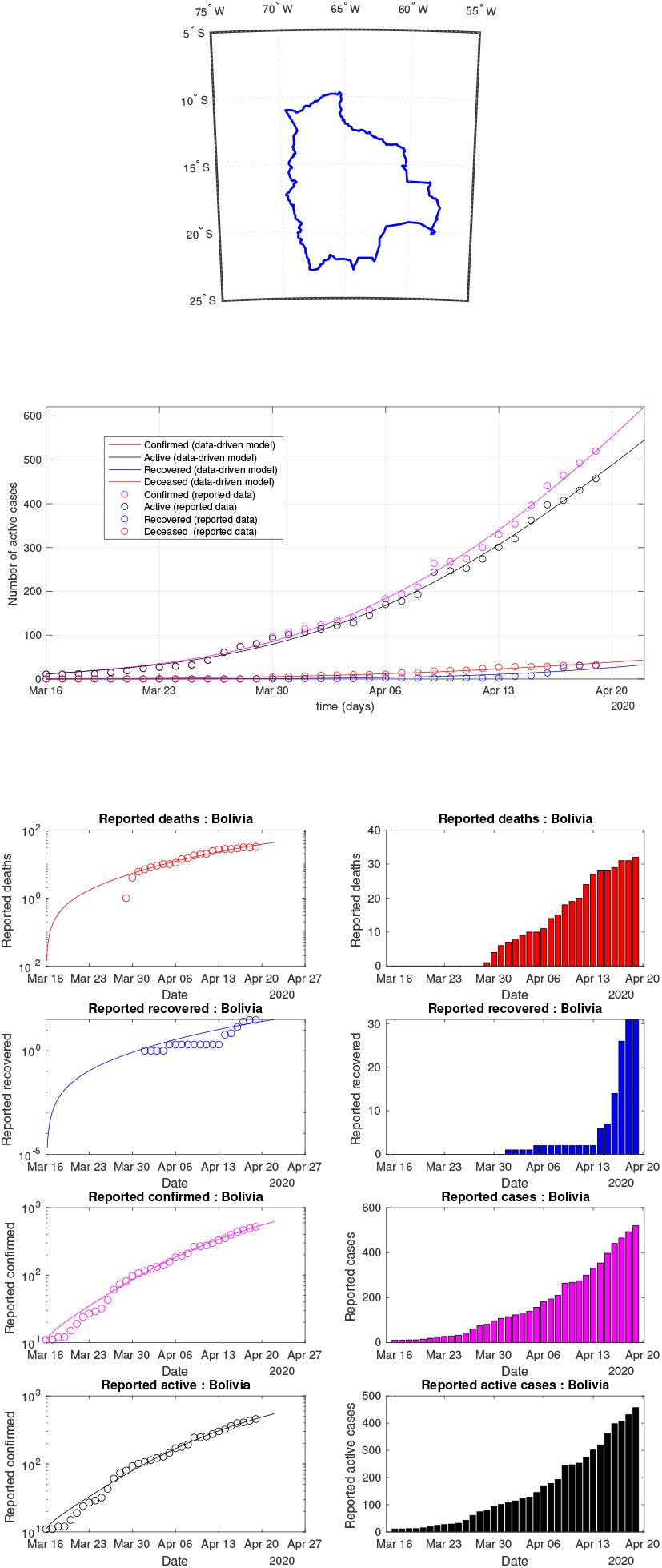
Bolivia: data of active, recovered, deceased (in circle or bar) vs model of active, recovered, deceased over time. Tracking the number of active cases. Predictive analytics for the next couple days

**Fig 66:**
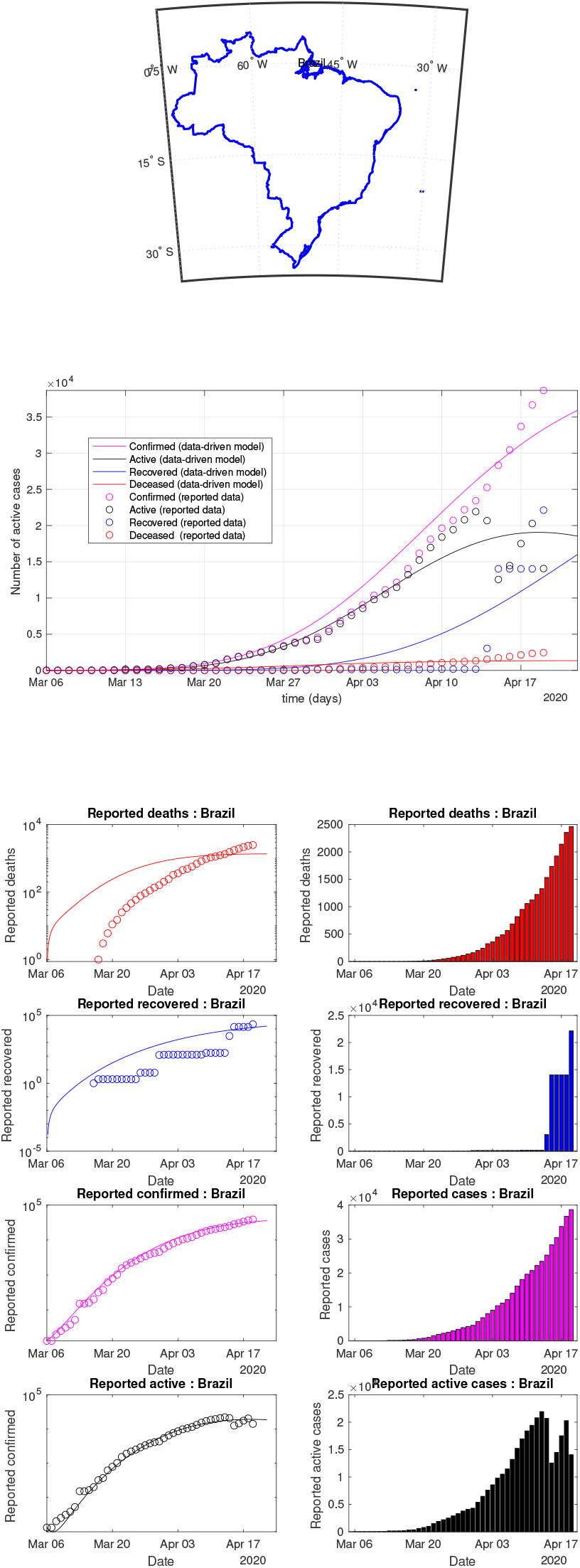
Brazil: data of active, recovered, deceased (in circle or bar) vs model of active, recovered, deceased over time. Tracking the number of active cases. Predictive analytics for the next couple days

**Fig 67:**
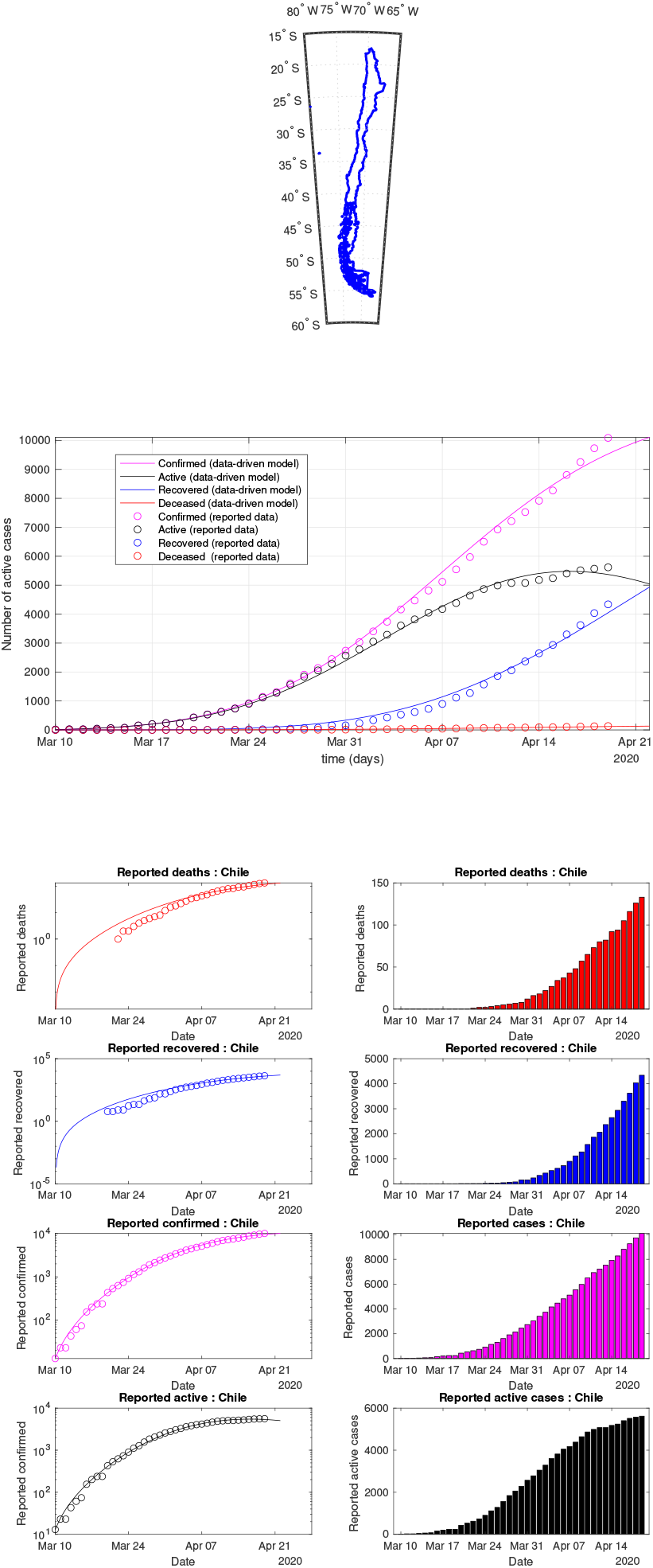
Chile: data of active, recovered, deceased (in circle or bar) vs model of active, recovered, deceased over time. Tracking the number of active cases. Predictive analytics for the next couple days

**Fig 68:**
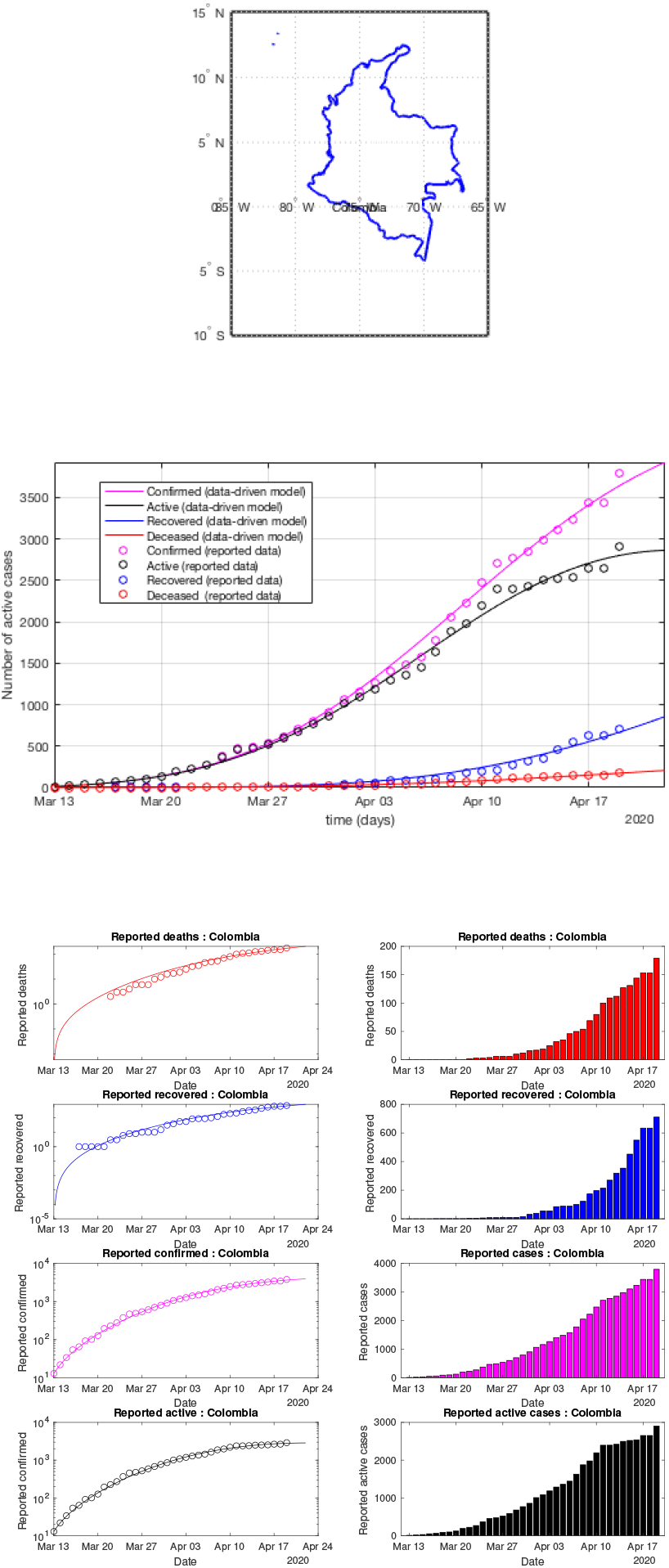
Colombia: data of active, recovered, deceased (in circle or bar) vs model of active, recovered, deceased over time. Tracking the number of active cases. Predictive analytics for the next couple days

**Fig 69:**
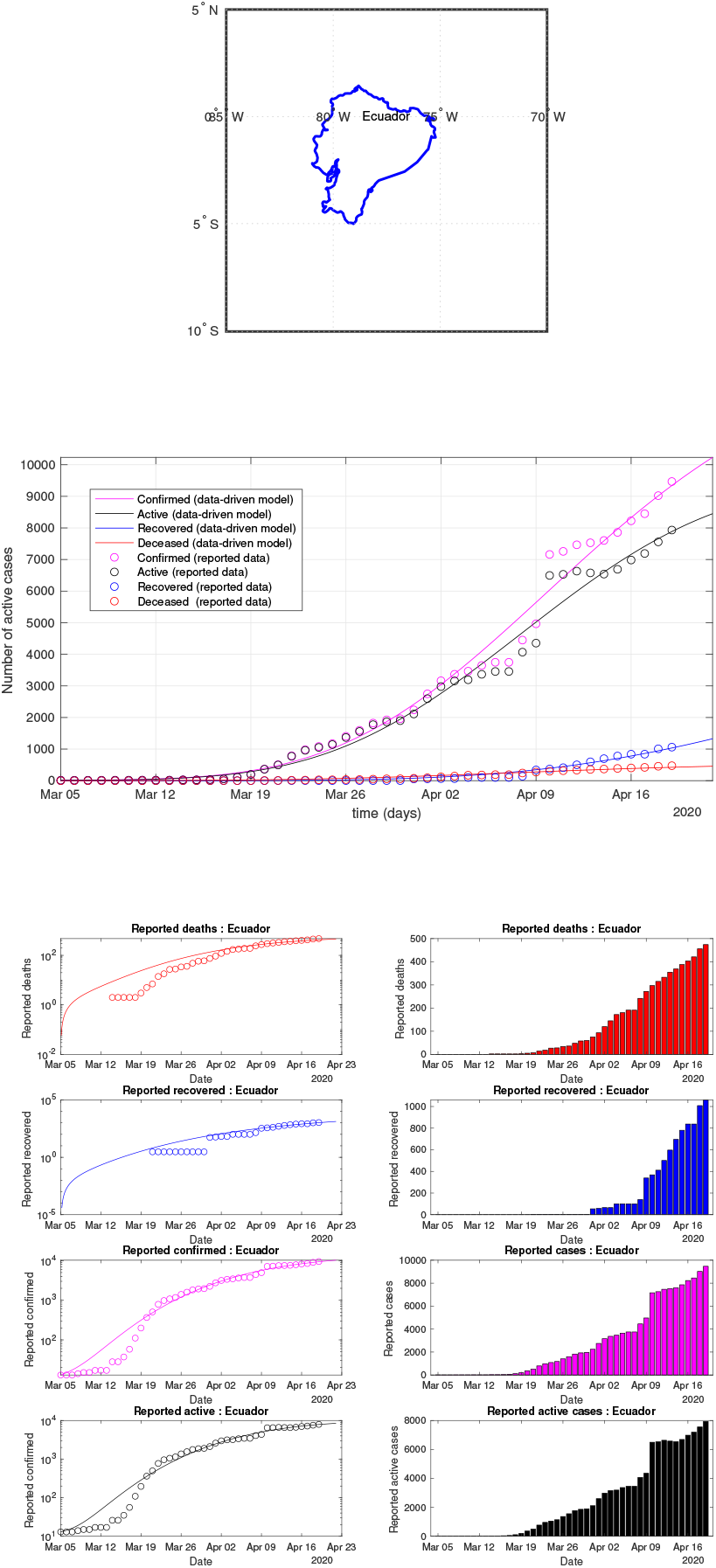
Ecuador: data of active, recovered, deceased (in circle or bar) vs model of active, recovered, deceased over time. Tracking the number of active cases. Predictive analytics for the next couple days

**Fig 70:**
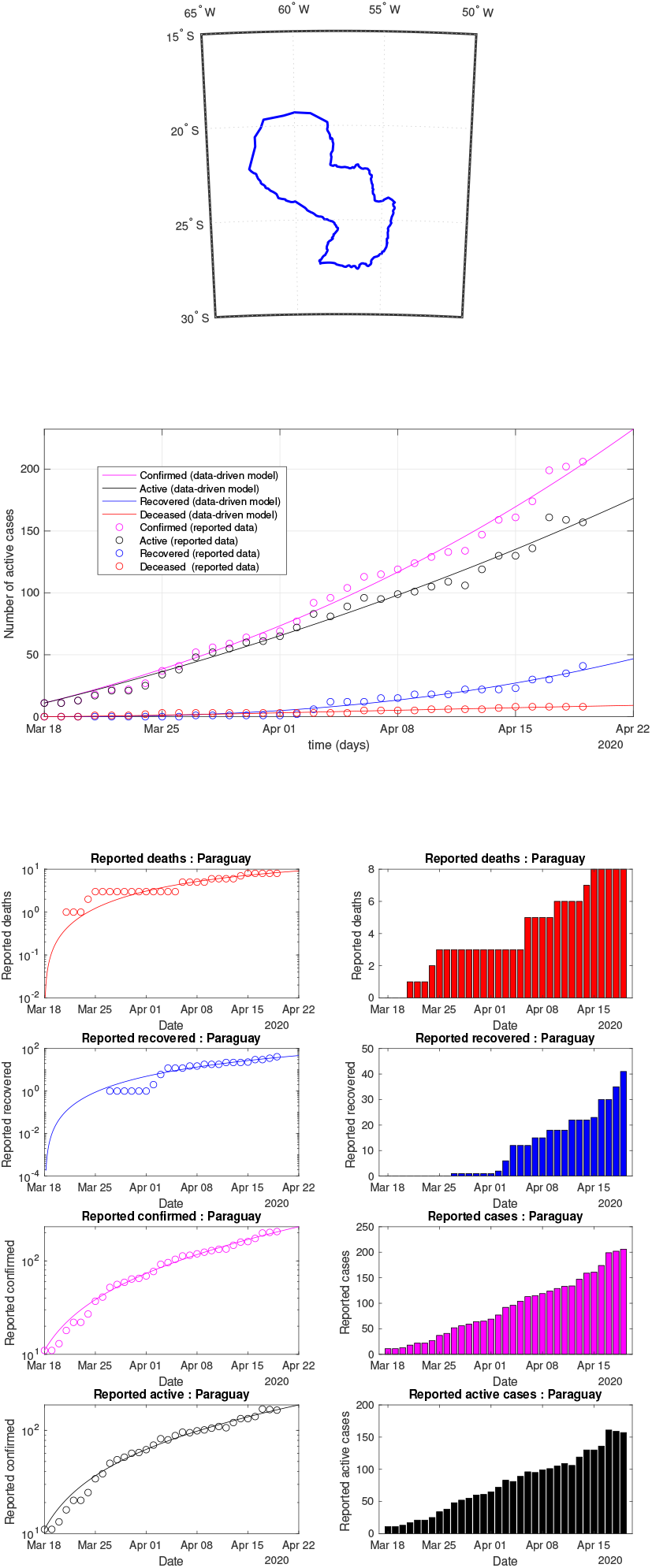
Paraguay: data of active, recovered, deceased (in circle or bar) vs model of active, recovered, deceased over time. Tracking the number of active cases. Predictive analytics for the next couple days

**Fig 71:**
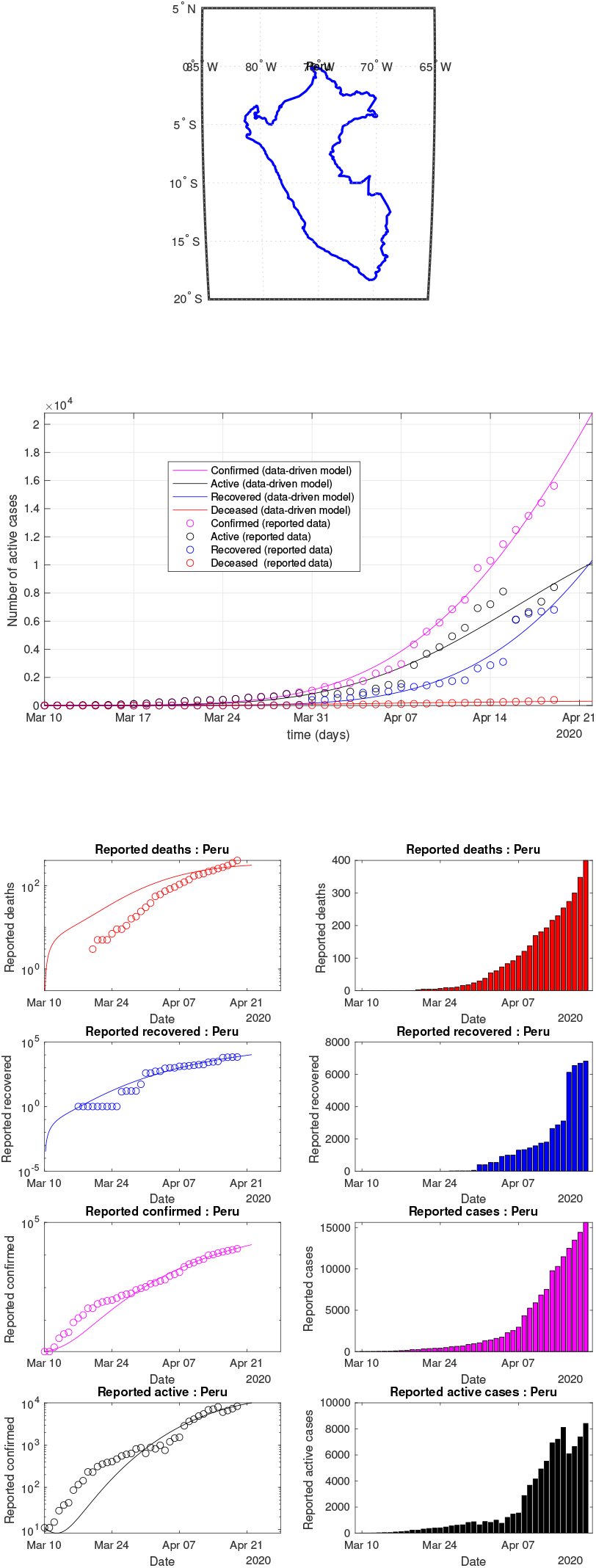
Peru: data of active, recovered, deceased (in circle or bar) vs model of active, recovered, deceased over time. Tracking the number of active cases. Predictive analytics for the next couple days

**Fig 72:**
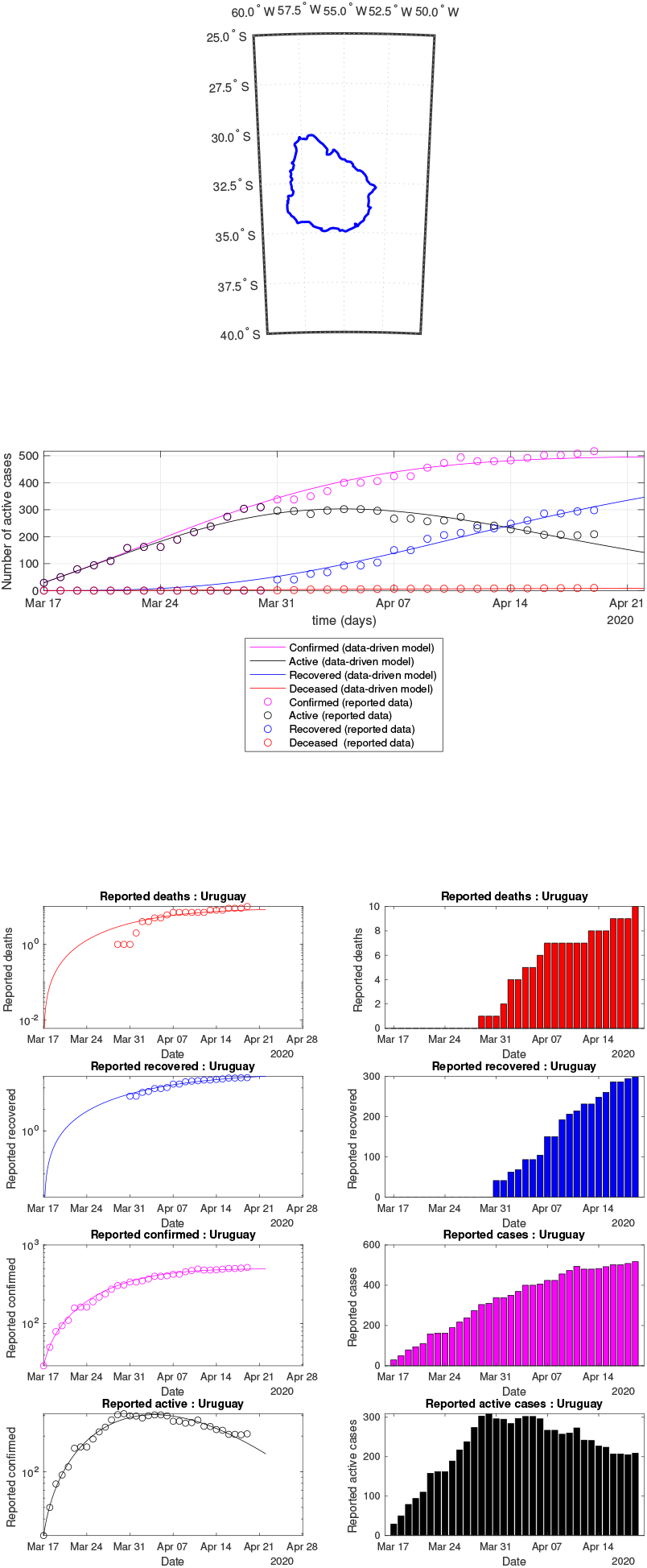
Uruguay: data of active, recovered, deceased (in circle or bar) vs model of active, recovered, deceased over time. Tracking the number of active cases. Predictive analytics for the next couple days

**Fig 73:**
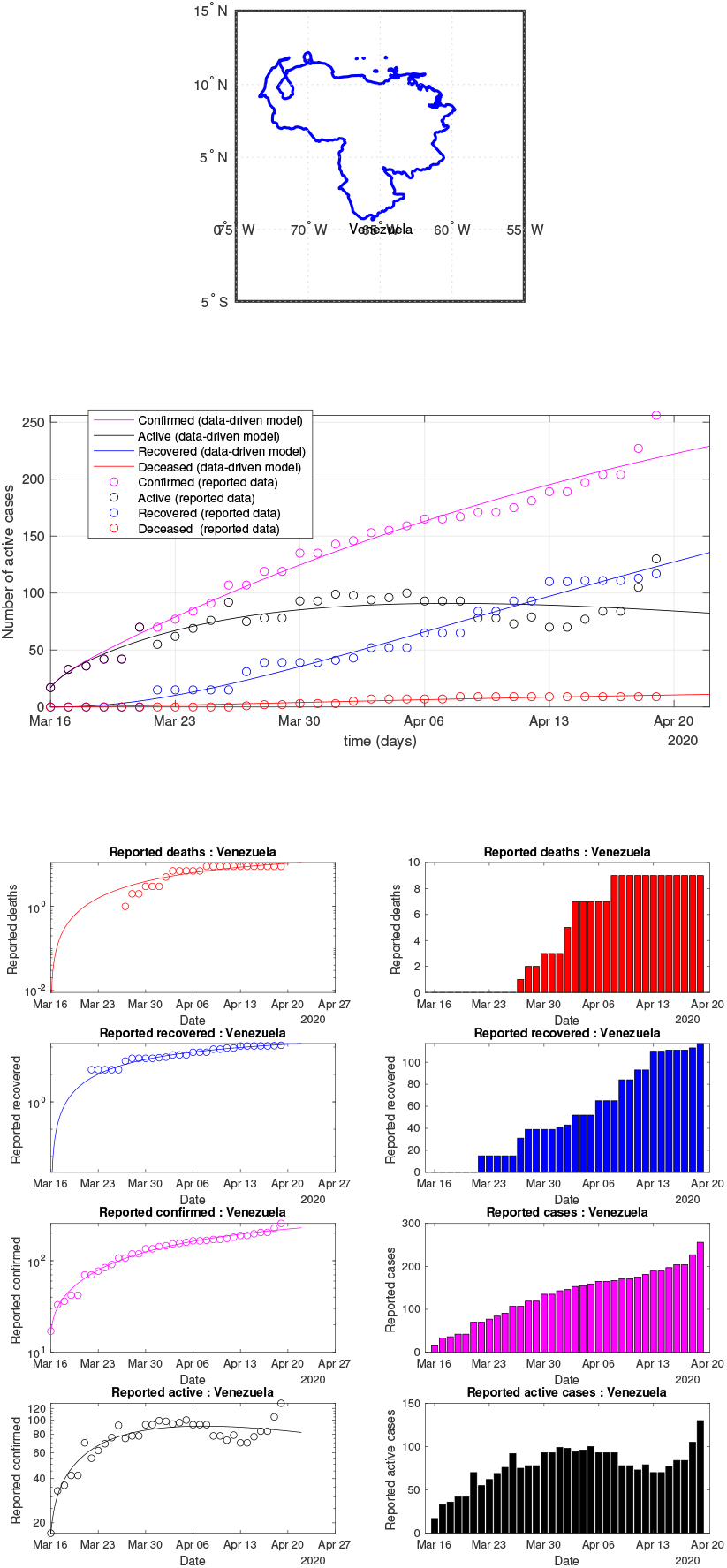
Venezuela: data of active, recovered, deceased (in circle or bar) vs model of active, recovered, deceased over time. Tracking the number of active cases. Predictive analytics for the next couple days

**Fig 74:**
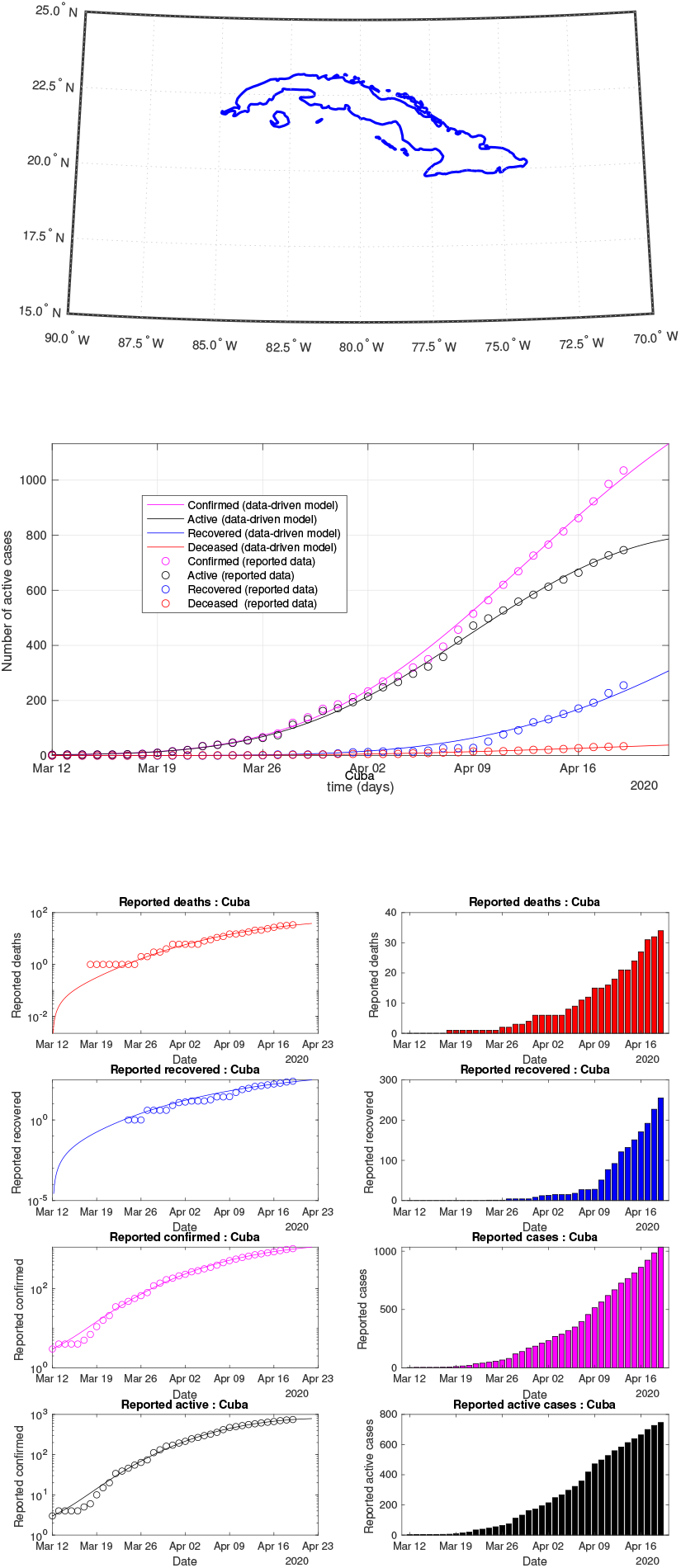
Cuba: data of active, recovered, deceased (in circle or bar) vs model of active, recovered, deceased over time. Tracking the number of active cases. Predictive analytics for the next couple days

**Fig 75:**
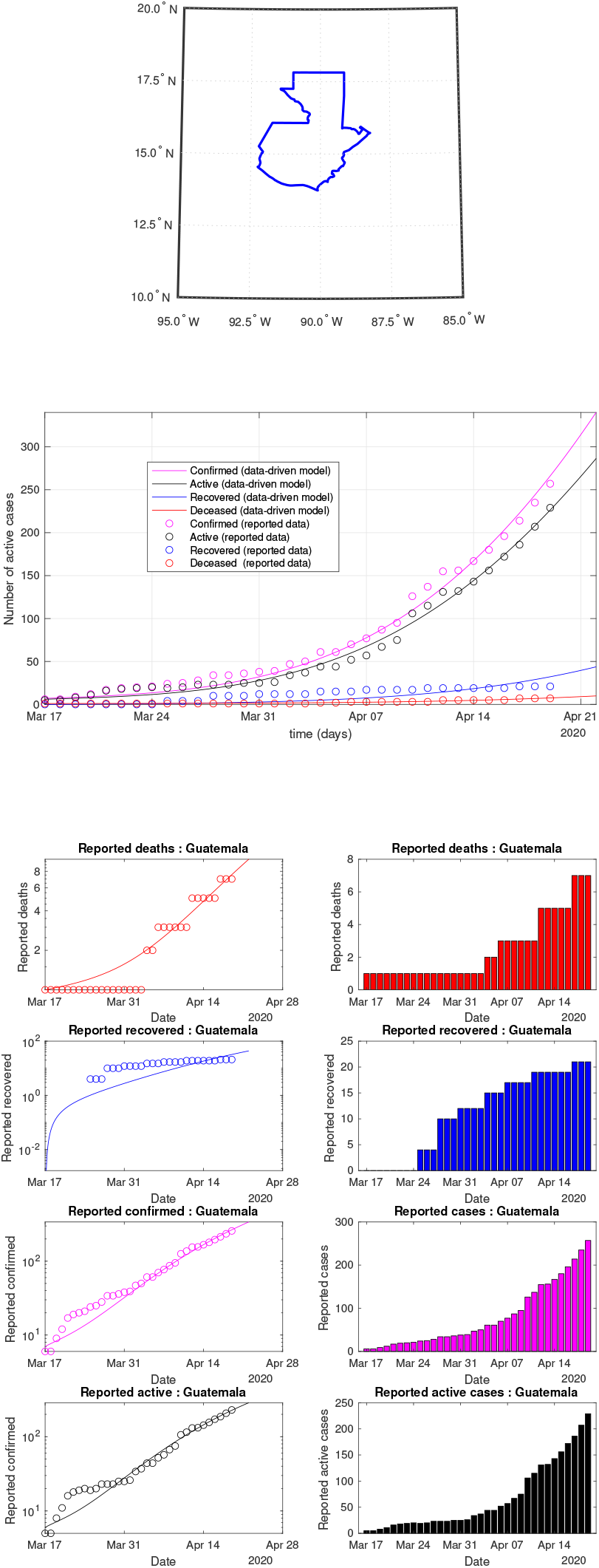
Guatemala: data of active, recovered, deceased (in circle or bar) vs model of active, recovered, deceased over time. Tracking the number of active cases. Predictive analytics for the next couple days

**Fig 76:**
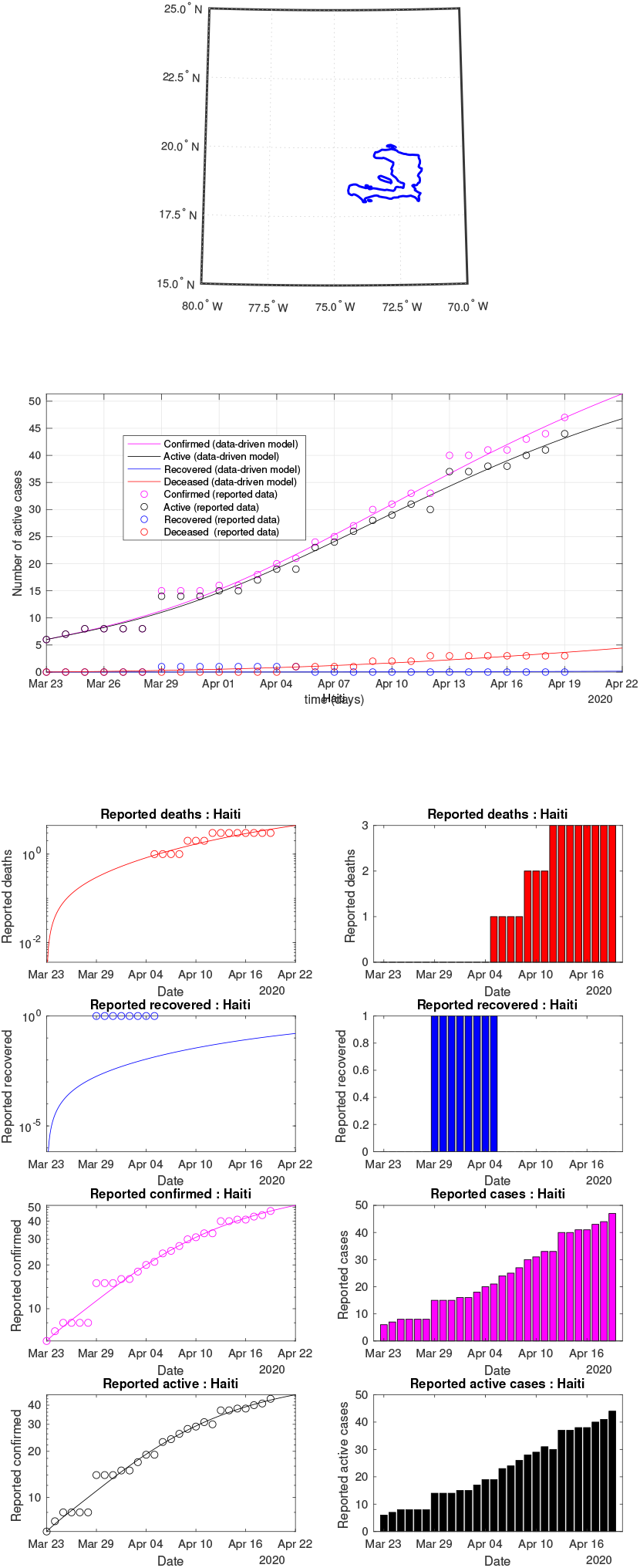
Haiti: data of active, recovered, deceased (in circle or bar) vs model of active, recovered, deceased over time. Tracking the number of active cases. Predictive analytics for the next couple days

**Fig 77:**
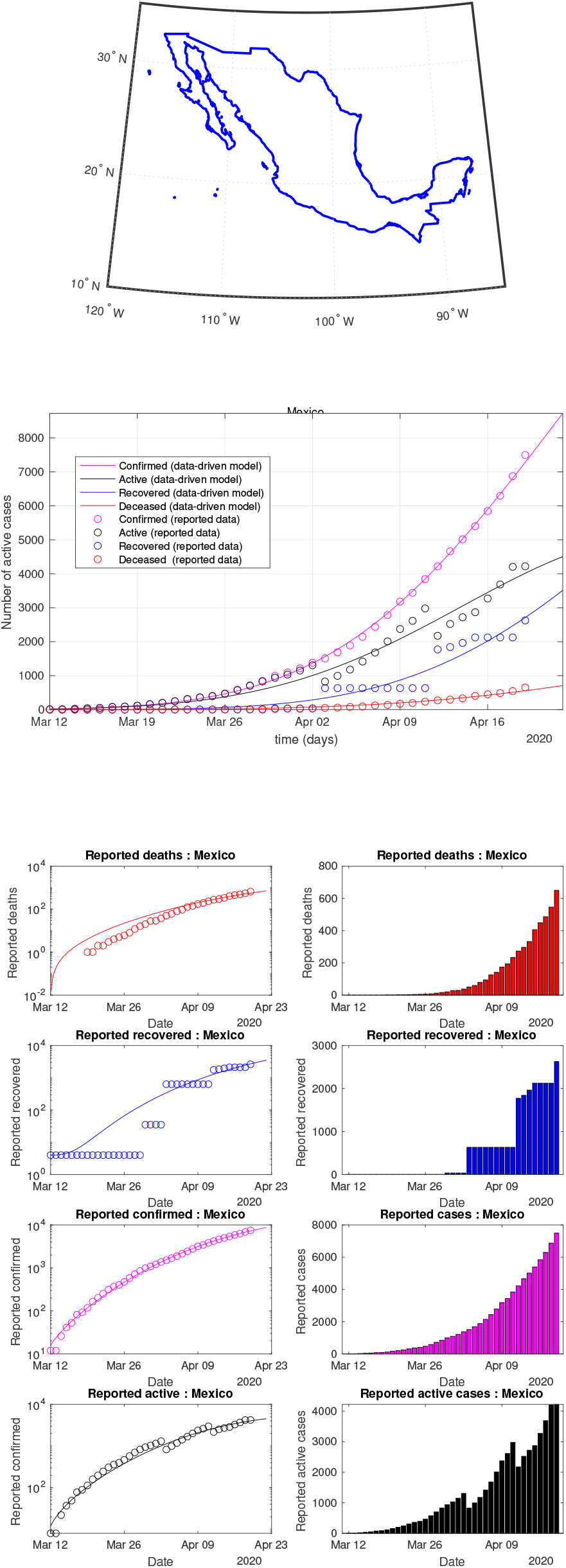
Mexico: data of active, recovered, deceased (in circle or bar) vs model of active, recovered, deceased over time. Tracking the number of active cases over time. Predictive analytics for the next couple days

**Fig 78:**
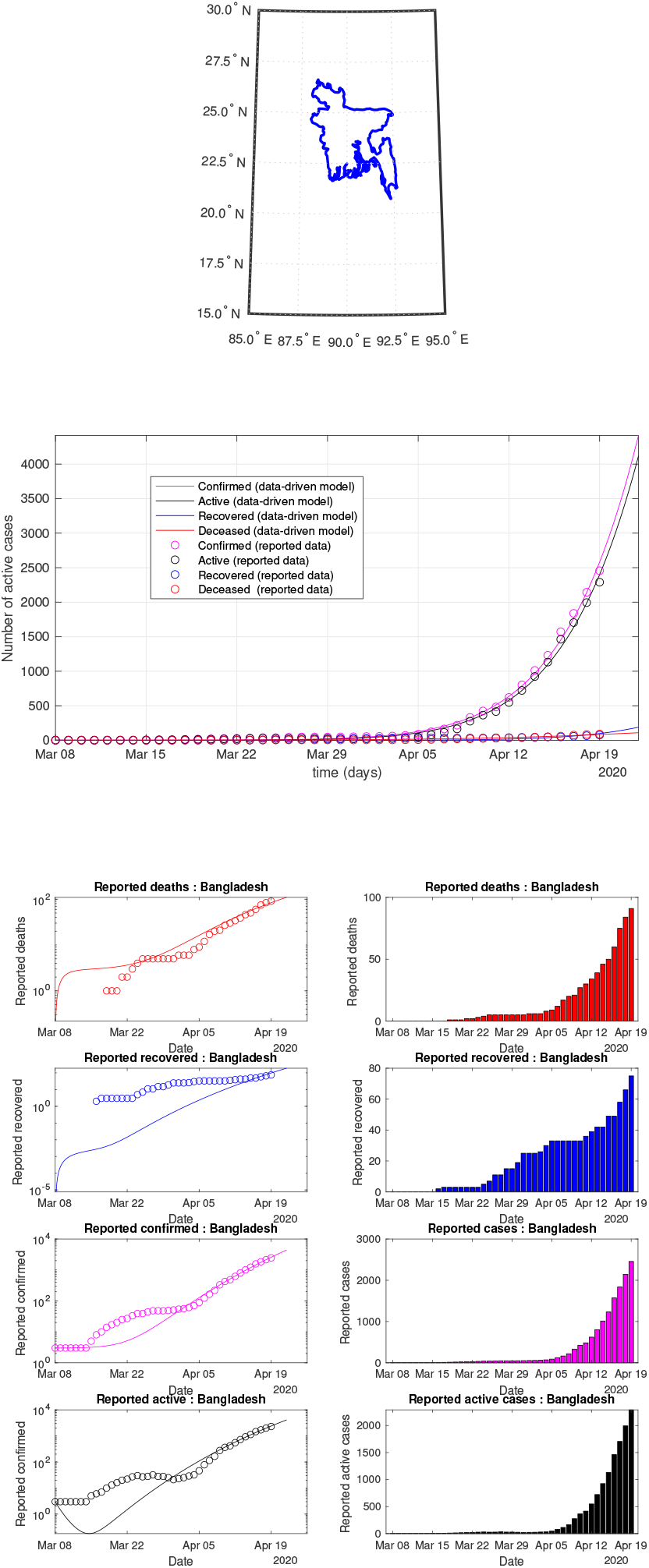
Bangladesh: data of active, recovered, deceased (in circle or bar) vs model of active, recovered, deceased over time. Tracking the number of active cases. Predictive analytics for the next couple days

**Fig 79:**
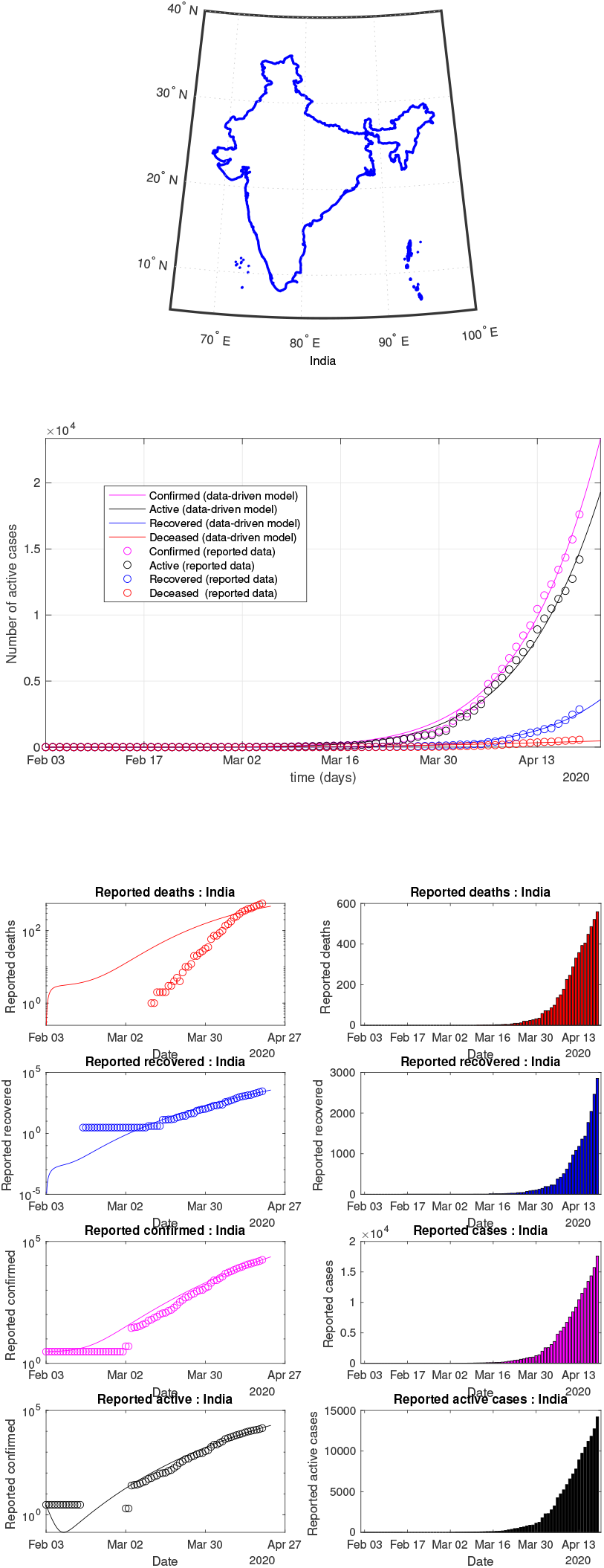
India: data of active, recovered, deceased (in circle or bar) vs model of active, recovered, deceased over time. Tracking the number of active cases. Predictive analytics for the next couple days

**Fig 80:**
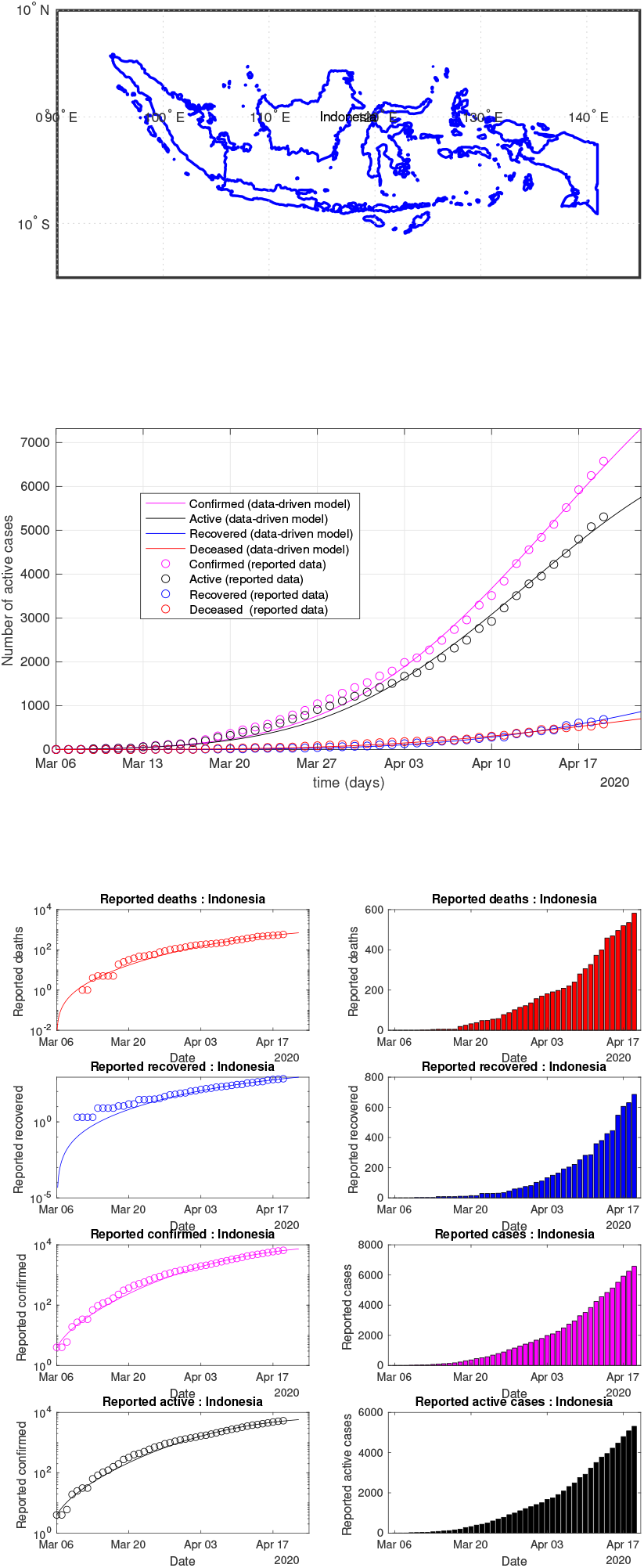
Indonesia: data of active, recovered, deceased (in circle or bar) vs model of active, recovered, deceased over time. Tracking the number of active cases. Predictive analytics for the next couple days

**Fig 81:**
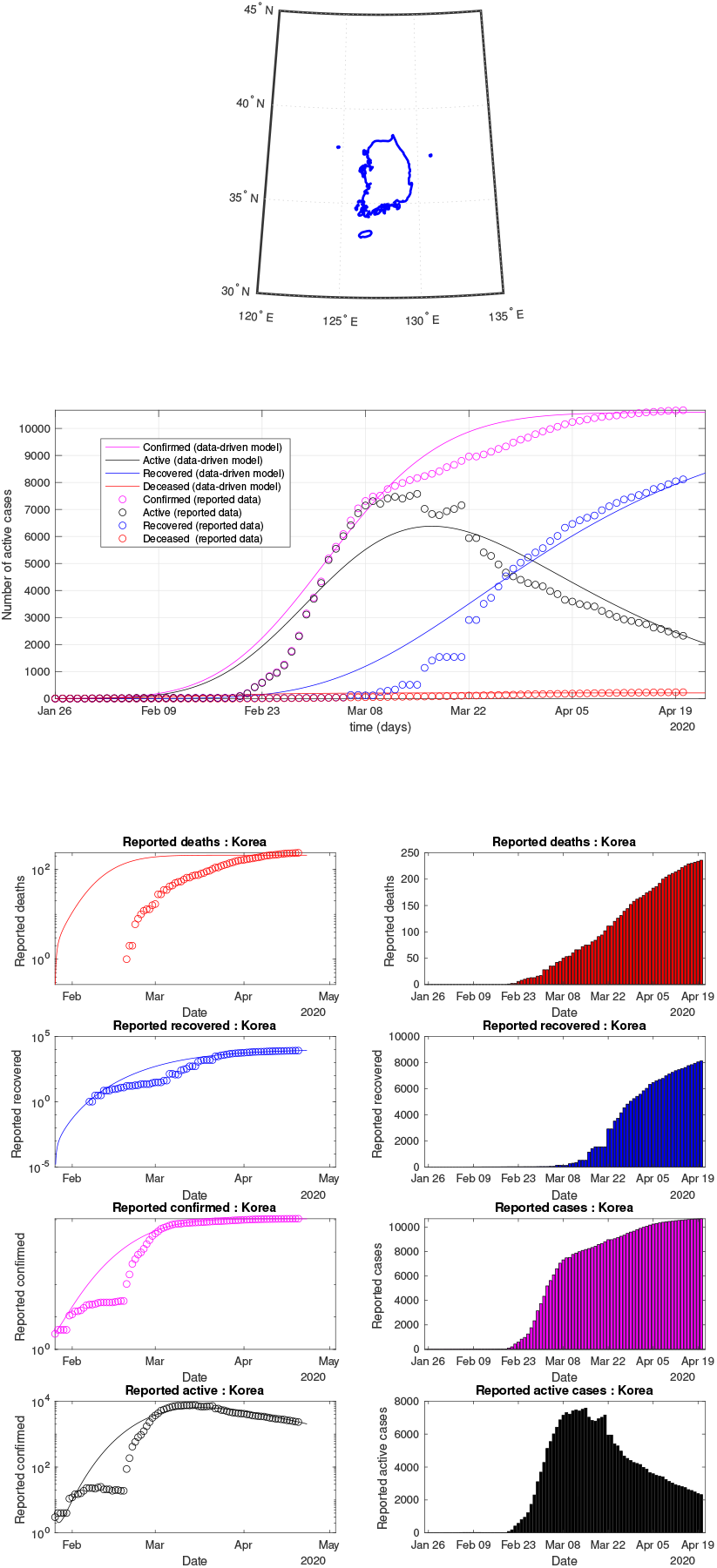
South Korea: data of active, recovered, deceased (in circle or bar) vs model of active, recovered, deceased over time. Tracking the number of active cases. Predictive analytics for the next couple days

**Fig 82:**
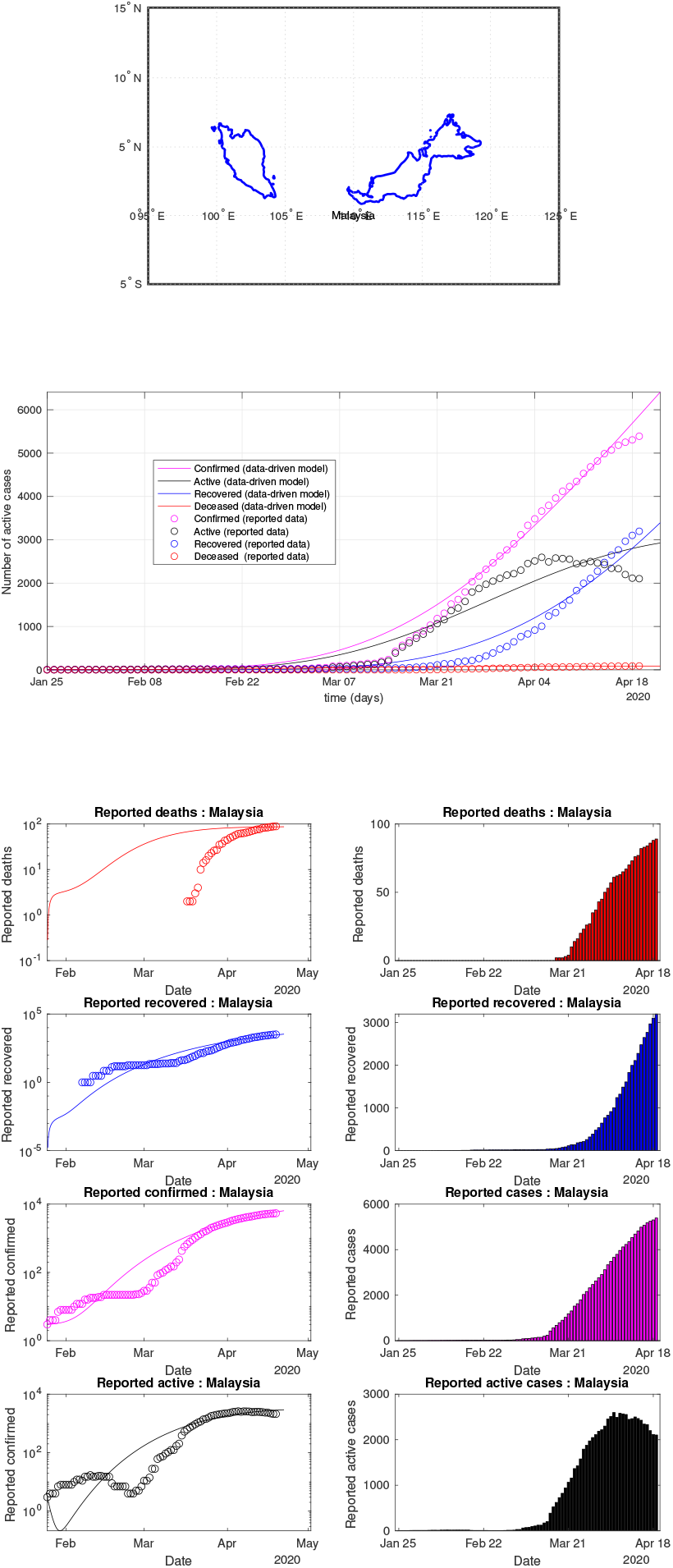
Malaysia: data of active, recovered, deceased (in circle or bar) vs model of active, recovered, deceased over time. Tracking the number of active cases. Predictive analytics for the next couple days

**Fig 83:**
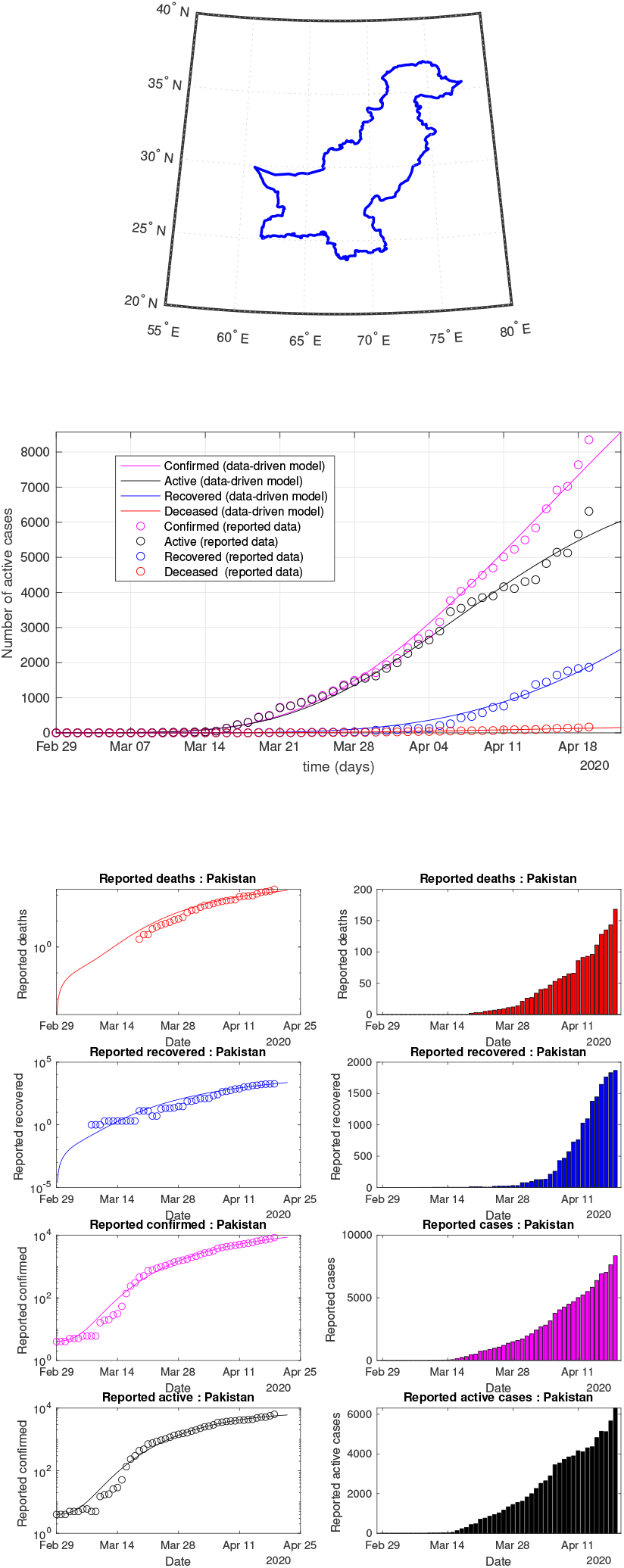
Pakistan: data of active, recovered, deceased (in circle or bar) vs model of active, recovered, deceased over time. Tracking the number of active cases. Predictive analytics for the next couple days

**Fig 84:**
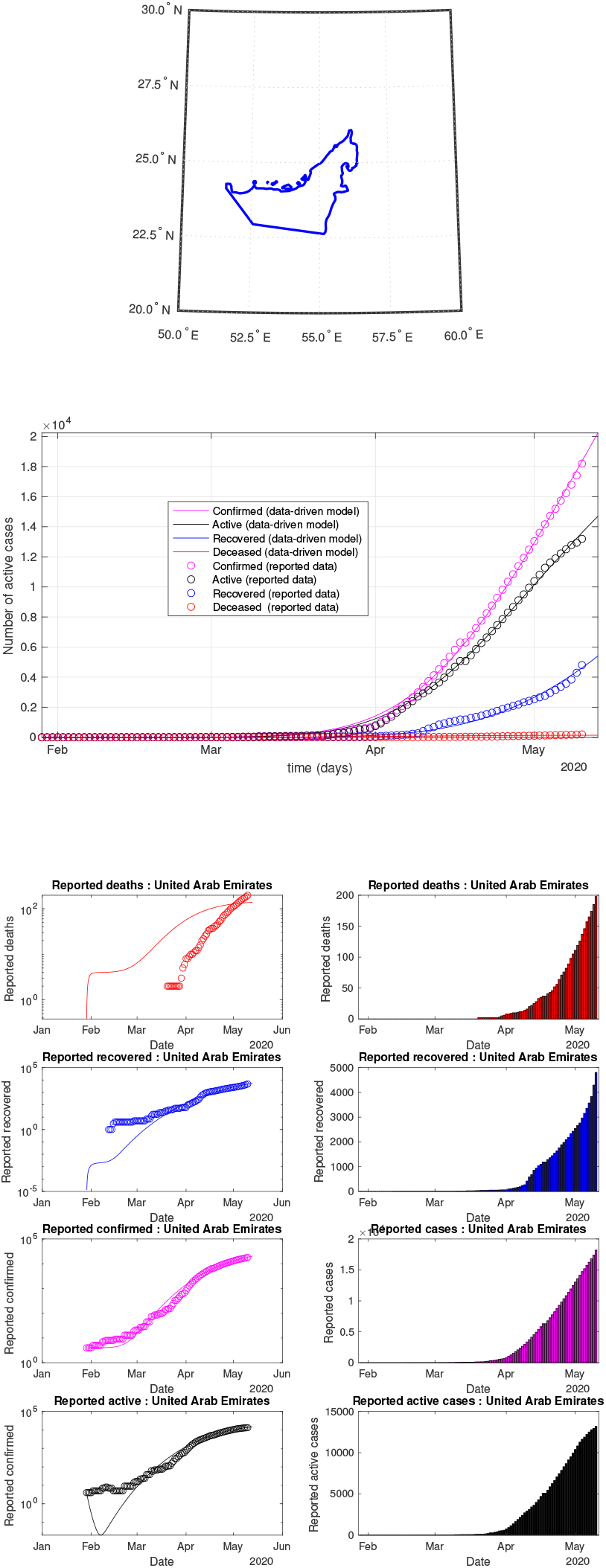
UAE: data of active, recovered, deceased (in circle or bar) vs model of active, recovered, deceased over time. Tracking the number of active cases. Predictive analytics for the next couple days

**Fig 85:**
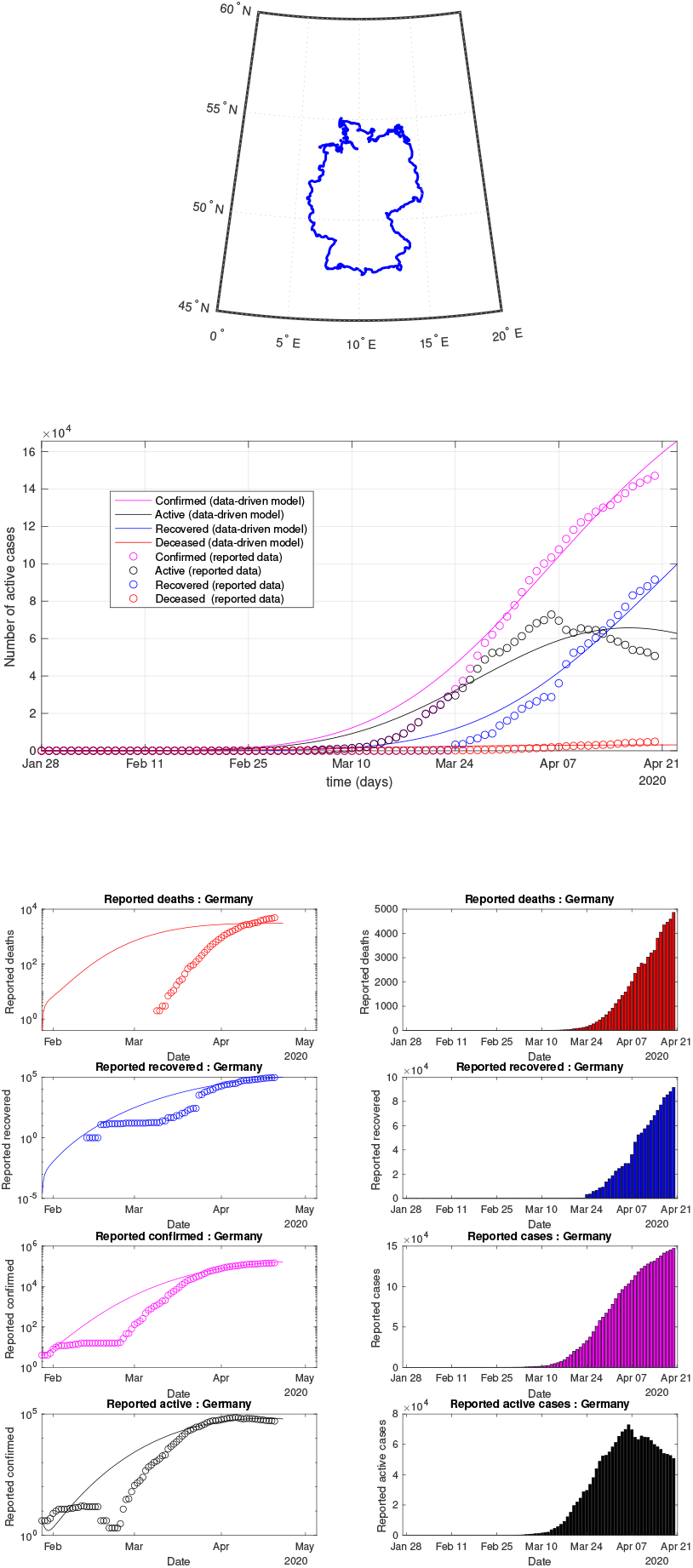
Germany: data of active, recovered, deceased (in circle or bar) vs model of active, recovered, deceased over time. Tracking the number of active cases. Predictive analytics for the next couple days

**Fig 86:**
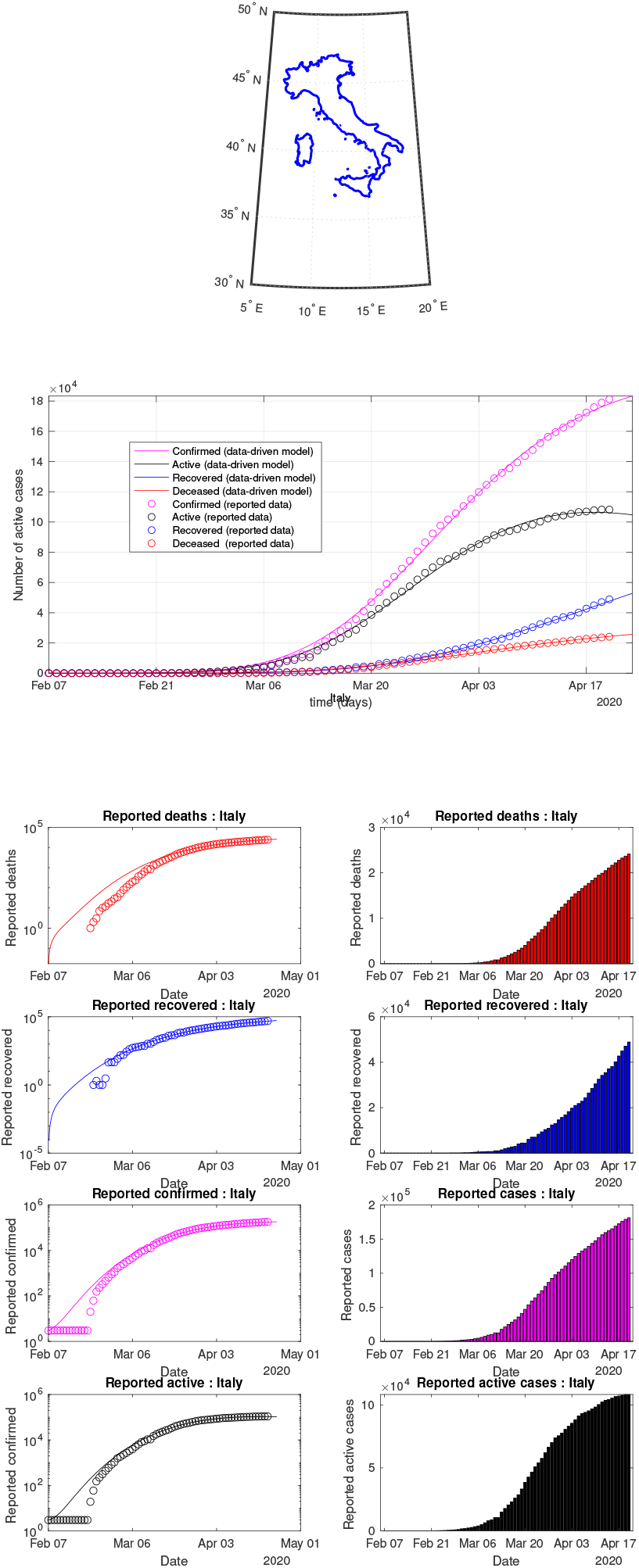
Italy: data of active, recovered, deceased (in circle or bar) vs model of active, recovered, deceased over time. Tracking the number of active cases. Predictive analytics for the next couple days

**Fig 87:**
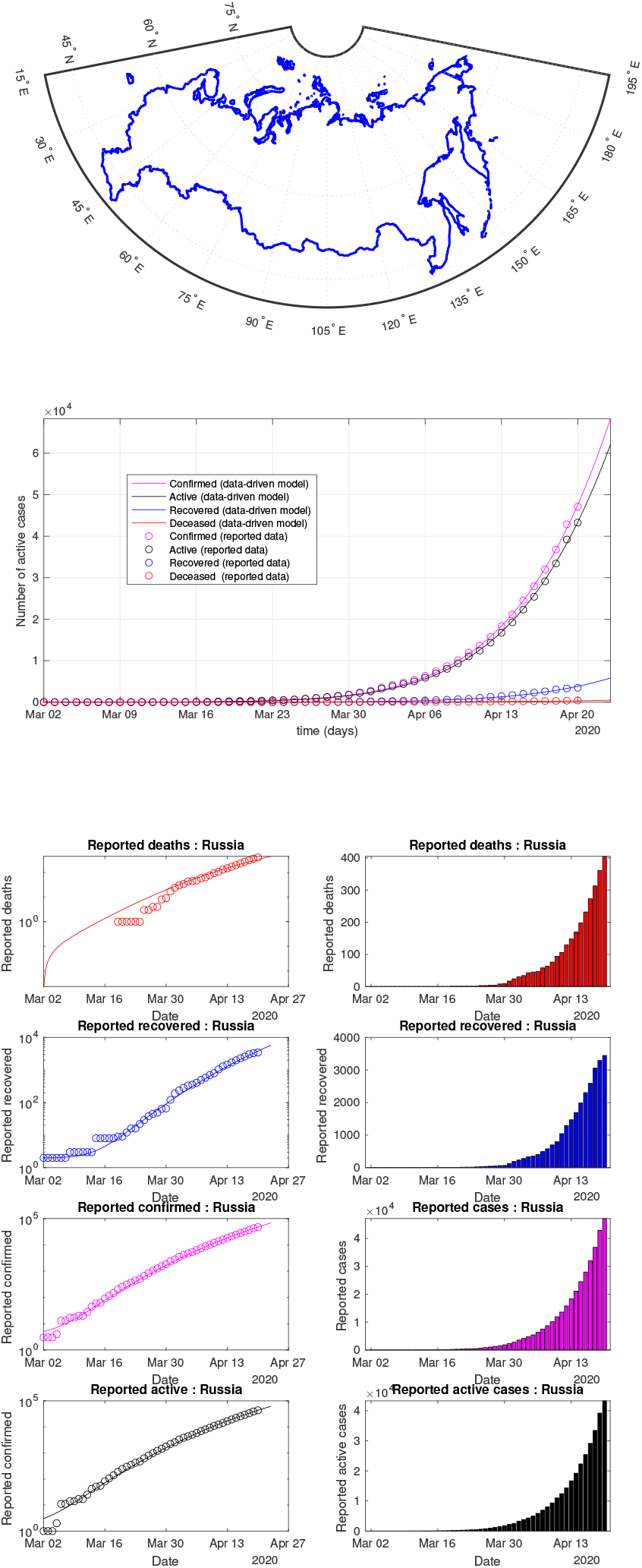
Russia: data of active, recovered, deceased (in circle or bar) vs model of active, recovered, deceased over time. Tracking the number of active cases. Predictive analytics for the next couple days

**Fig 88:**
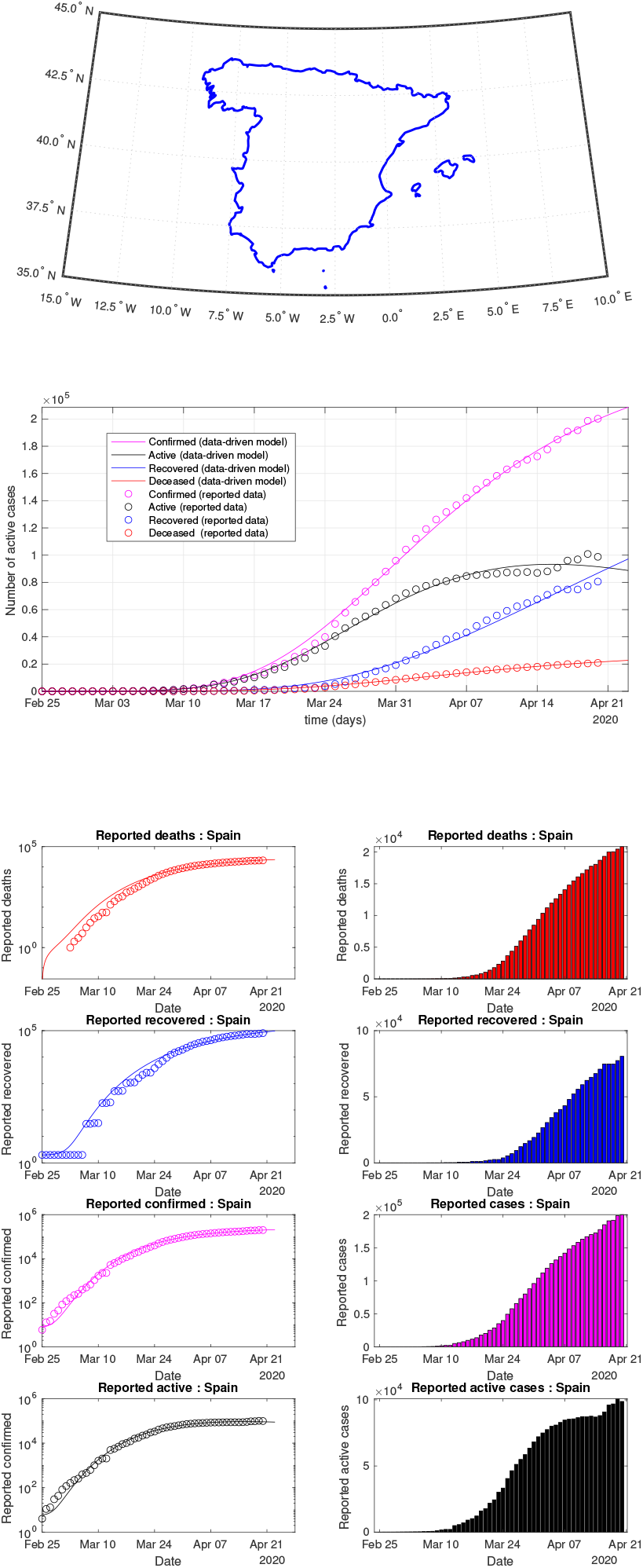
Spain: data of active, recovered, deceased (in circle or bar) vs model of active, recovered, deceased over time. Tracking the number of active cases. Predictive analytics for the next couple days

**Fig 89:**
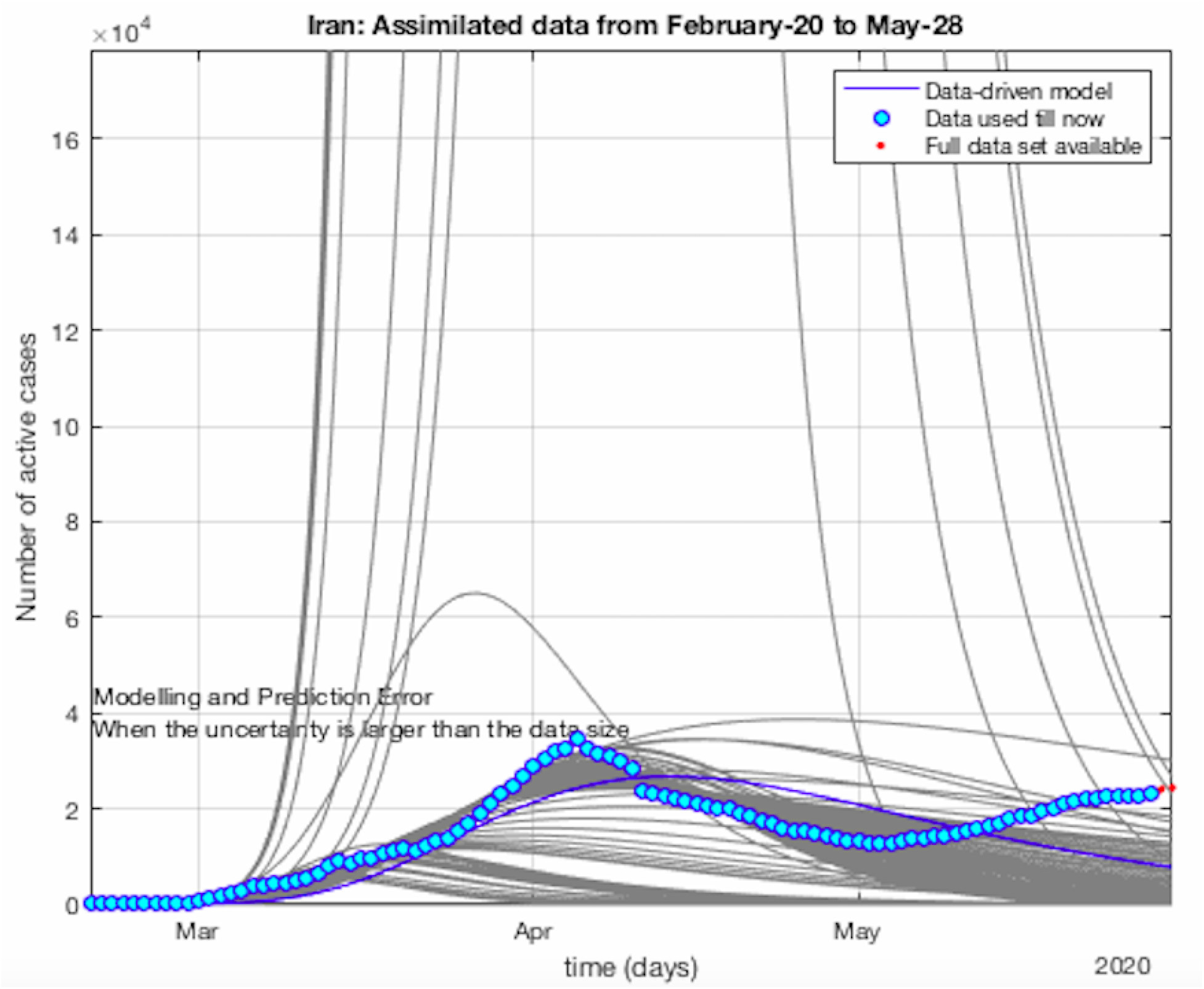
COVID-19 sample of Non-Gaussianity and non-exponential. The plots are obtained by using the data set sequentially till the latest data point. A **Non-Gausianity** of the reported active cases and hospitalized cases is observed. The shape of the sequential data-driven curve is clearly not a single exponential.

**Fig 90:**
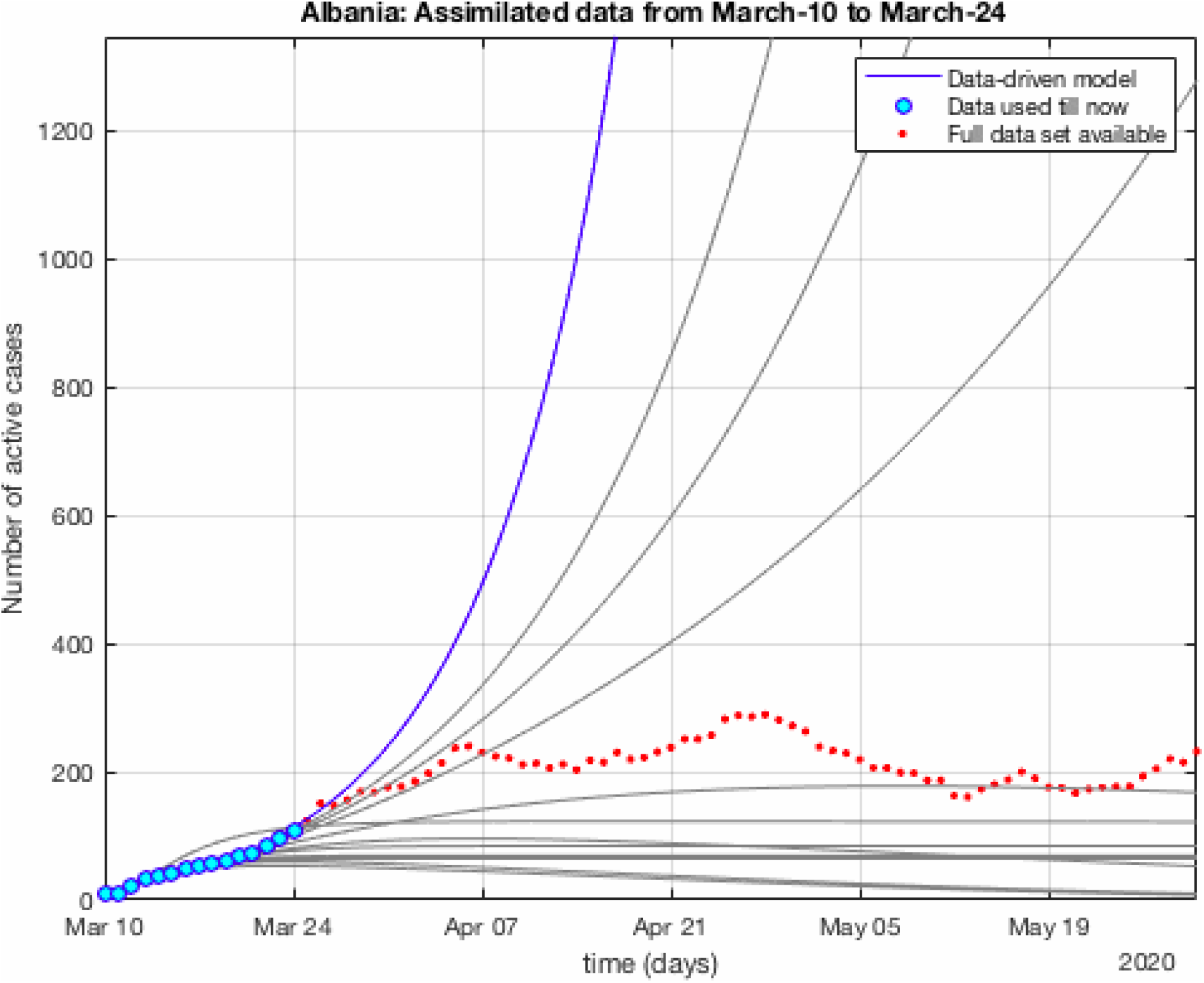
Another COVID-19 sample of Non-Gaussianity and non-exponential. The plots are obtained by using the data set sequentially till the latest data point. A **Non-Gausianity** of the reported active cases and hospitalized cases is observed. The shape of the sequential data-driven curve is clearly not a single exponential.

**TABLE IV:** List 1 of figures and illustrations

| Illustration | Meaning |
| --- | --- |
| Figure 1 | Flow diagram |
| Figure 2 | first wave of spread |
| Figure 3 | death rate distribution by age |
| Figure 4 | sample age distribution |
| Figure 10 | evolution of the epidemiological ODE with 17 states |
| Figure 6 | connection with SIR |
| Figure 7 | connection with SEIR |
| Figure 8 | connection with SIRD |
| Figure 9 | connection with SEIRD |
| Figure 11 | pandemic vs economic<br>consequences of COVID-19 |
| Figure 12 | spread worldwide |
| Figure 13 | propagation of the virus in<br>15 most reported active cases<br>as of April 11, 2020 |
| Figure 14 | G7 countries |
| Figure 15 | New York and Los Angeles<br>testing results: data vs model |
| Figure 16 | Brazil: Sequential data-driven model |
| Figure 17 | USA: Sequential data-driven model |

**TABLE V:** List 2 of figures and illustrations. data of active, recovered, deceased (in circle or bar) vs model of active, recovered, deceased over time. Tracking the number of active cases. Predictive analytics for the next couple days

| Illustration | Meaning |
| --- | --- |
| Figure 64 | Argentina : data vs model |
| Figure 65 | Bolivia: data vs model |
| Figure 66 | Brazil : data vs model |
| Figure 67 | Chile : data vs model |
| Figure 68 | Colombia : data vs model |
| Figure 69 | Ecuador : data vs model |
| Figure 70 | : data vs model |
| Figure 71 | Paraguay : data vs model |
| Figure 72 | Uruguay : data vs model |
| Figure 73 | Venezuela : data vs model |
| Figure 74 | Cuba : data vs model |
| Figure 75 | Guatemala : data vs model |
| Figure 76 | Haiti : data vs model |
| Figure 77 | Mexico : data vs model |
| Figure 78 | Bangladesh : data vs model |
| Figure 79 | India : data vs model |
| Figure 80 | Indonesia : data vs model |
| Figure 81 | Korea : data vs model |
| Figure 82 | Malaysia : data vs model |
| Figure 83 | Pakistan : data vs model |
| Figure 84 | UAE : data vs model |
| Figure 85 | Germany : data vs model |
| Figure 86 | Italy : data vs model |
| Figure 87 | Russia : data vs model |
| Figure 88 | Spain : data vs model |
| Figure 89 | Non-Gaussianity and non-exponential |
| Figure 90 | Not a single exponential |

**TABLE VI:**
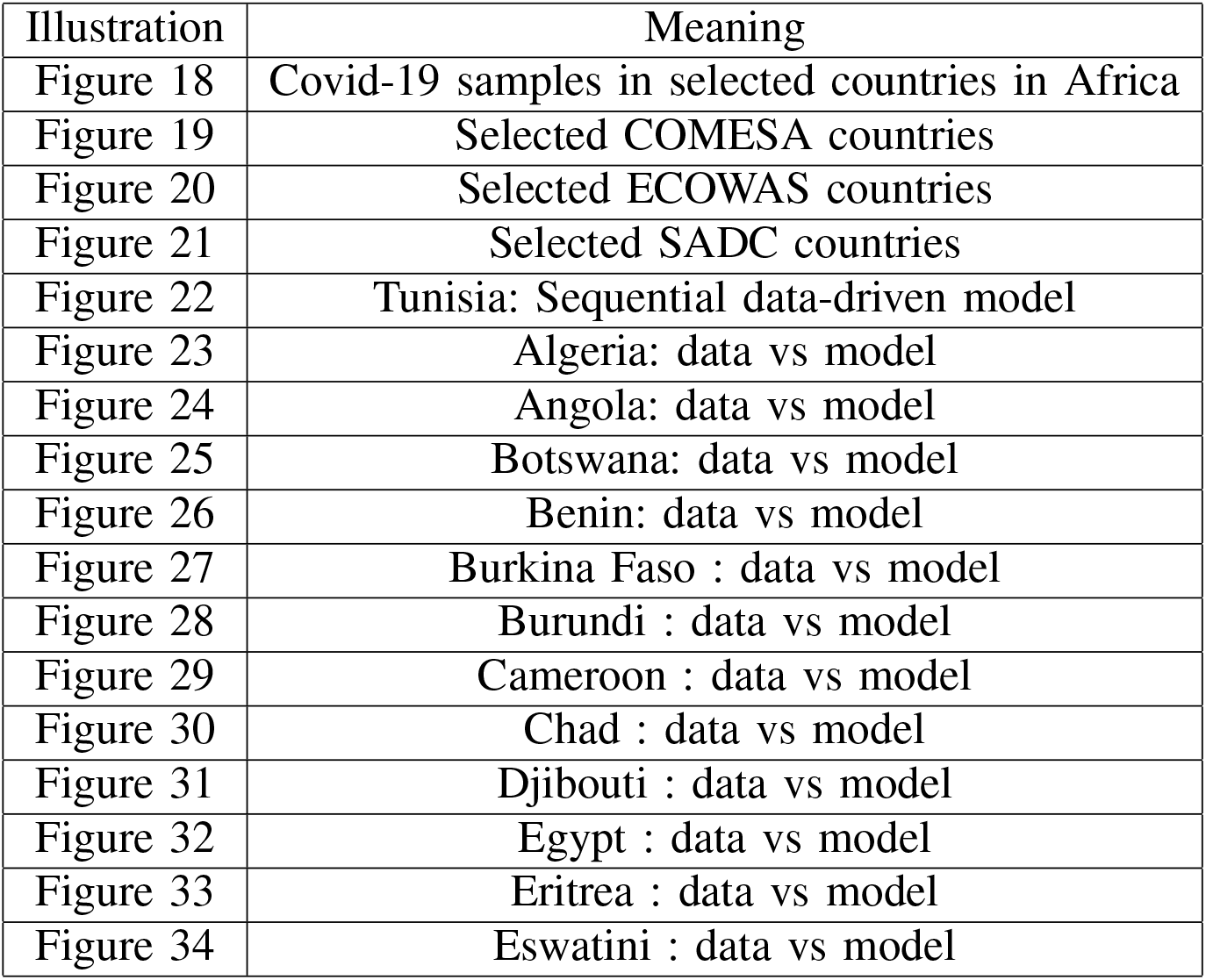
List 3 of figures and illustrations. data of active, recovered, deceased (in circle or bar) vs model of active, recovered, deceased over time. Tracking the number of active cases. Predictive analytics for the next couple days

| Illustration | Meaning |
| --- | --- |
| Figure 18 | Covid-19 samples in selected countries in Africa |
| Figure 19 | Selected COMESA countries |
| Figure 20 | Selected ECOWAS countries |
| Figure 21 | Selected SADC countries |
| Figure 22 | Tunisia: Sequential data-driven model |
| Figure 23 | Algeria: data vs model |
| Figure 24 | Angola: data vs model |
| Figure 25 | Botswana: data vs model |
| Figure 26 | Benin: data vs model |
| Figure 27 | Burkina Faso : data vs model |
| Figure 28 | Burundi : data vs model |
| Figure 29 | Cameroon : data vs model |
| Figure 30 | Chad : data vs model |
| Figure 31 | Djibouti : data vs model |
| Figure 32 | Egypt : data vs model |
| Figure 33 | Eritrea : data vs model |
| Figure 34 | Eswatini : data vs model |

**TABLE VII:** List 4 of figures and illustrations. data of active, recovered, deceased (in circle or bar) vs model of active, recovered, deceased over time. Tracking the number of active cases. Predictive analytics for the next couple days

| Illustration | Meaning |
| --- | --- |
| Figure 35 | Ethiopi : data vs model |
| Figure 36 | Gabon : data vs model |
| Figure 37 | Ghana : data vs model |
| Figure 38 | Guinea : data vs model |
| Figure 39 | Guinea Bissau : data vs model |
| Figure 40 | Kenya : data vs model |
| Figure 41 | Liberia : data vs model |
| Figure 42 | Libya : data vs model |
| Figure 43 | Madagascar : data vs model |
| Figure 44 | Malawi : data vs model |
| Figure 45 | Mali : data vs model |
| Figure 46 | Mauritania : data vs model |
| Figure 47 | Mauritius : data vs model |
| Figure 48 | Morocco : data vs model |
| Figure 49 | Mozambique : data vs model |
| Figure 50 | Namibia : data vs model |
| Figure 51 | Niger : data vs model |
| Figure 52 | Nigeria : data vs model |
| Figure 53 | Rwanda : data vs model |
| Figure 54 | Senegal : data vs model |
| Figure 55 | Seychelles : data vs model |
| Figure 56 | Somalia : data vs model |
| Figure 57 | South Africa : data vs model |
| Figure 58 | Sudan : data vs model |
| Figure 59 | Togo : data vs model |
| Figure 60 | Tunisia : data vs model |
| Figure 61 | Uganda : data vs model |
| Figure 62 | Zambia : data vs model |
| Figure 63 | Zimbabwe : data vs model |

## Limitations of the proposed model

The proposed data-driven modelling has several interesting features including epidemiological aspects (hospitalization, tested negative, untested active, untested deceased, untested active) as well as economics aspects (average revenue per family size, income status, on-site worked hours during COVID-19) and mobility aspects (pedestrian within a sector, within a city, between cities, provinces, countries by ground/air/sea transportation, and movement to hotspots, local markets, grocery stores, pharmacies, etc). However, the model still need extension and integration of food security and supply chain which are relevant aspects in many countries.

## Data Availability

https://github.com/CSSEGISandData/COVID-19

## Acknowledgment

The author would like to thank the six webinars’ audience at UCLA IPAM, NYU, Next Einstein Forum, CNRIA 2020, CDC meeting on COVID-19, and IEEE St-Maurice section for their valuable inputs. The author would like to thank Mamadou Lamine Doumbia, Boualem Djehiche, Nader Masmoudi, Julian Barreiro-Gomez, and Salah Eddine Choutri for interesting discussions on the topic of data-driven modelling. We gratefully acknowledge support from U.S. Air Force Office of Scientific Research under grant number FA9550-17-1-0259 on Mean-Field-Type Game Theory and the Center on Stability, Instability and Turbulence.

## Conclusion

Understanding the transmission characteristics of infectious diseases in communities, counties, provinces, regions, states and countries can lead to better approaches to decreasing the transmission of these diseases. In this paper, a context-aware data-driven MFTG model. The proposed epidemiological and economic model can track and captures the country-specific COVID-19 data in 66+ countries. The context-aware data-driven MFTG model can be used in visualizing, comparing, planning, implementing, evaluating, and optimizing various detection, testing, prevention and control programs.

Data-Driven MFTG modeling can contribute to the design and analysis of the pandemic COVID-19 surveys, suggest crucial data that should be collected, identify trends, make short-term forecasts, and quantify the uncertainties in short-term forecasting. In addition to predictive safety measures, COVID-19 opens also other challenges such as

- Supply chain: Identifying ways to optimize supply chain among many different industries, so the impact of the COVID-19 can be minimized.
- Testing, Tracing and Tracking: Finding ways to better test, trace and track the infection propagation including those asymptomatic infected, best practices in how to test, trace and track in both rural and urban areas as early as possible.
- Community: Finding solutions to prevent social and economic disruption in all communities. Proposing innovative de-confinement strategies in how to create a smooth transition from physical distancing to guaranteeing minimum services and quality-preserving activities and back to regular activities.

Number of technical questions remain unanswered. This include (i) impact of the propagation in local economy (including informal economy), (ii) the duration of the limit cycle of active cases, (iii) the stabilization of the system by means of controls and co-opetitive decision-making, (iv) impact of COVID-19 on food insecurity. We leave these questions for future research.

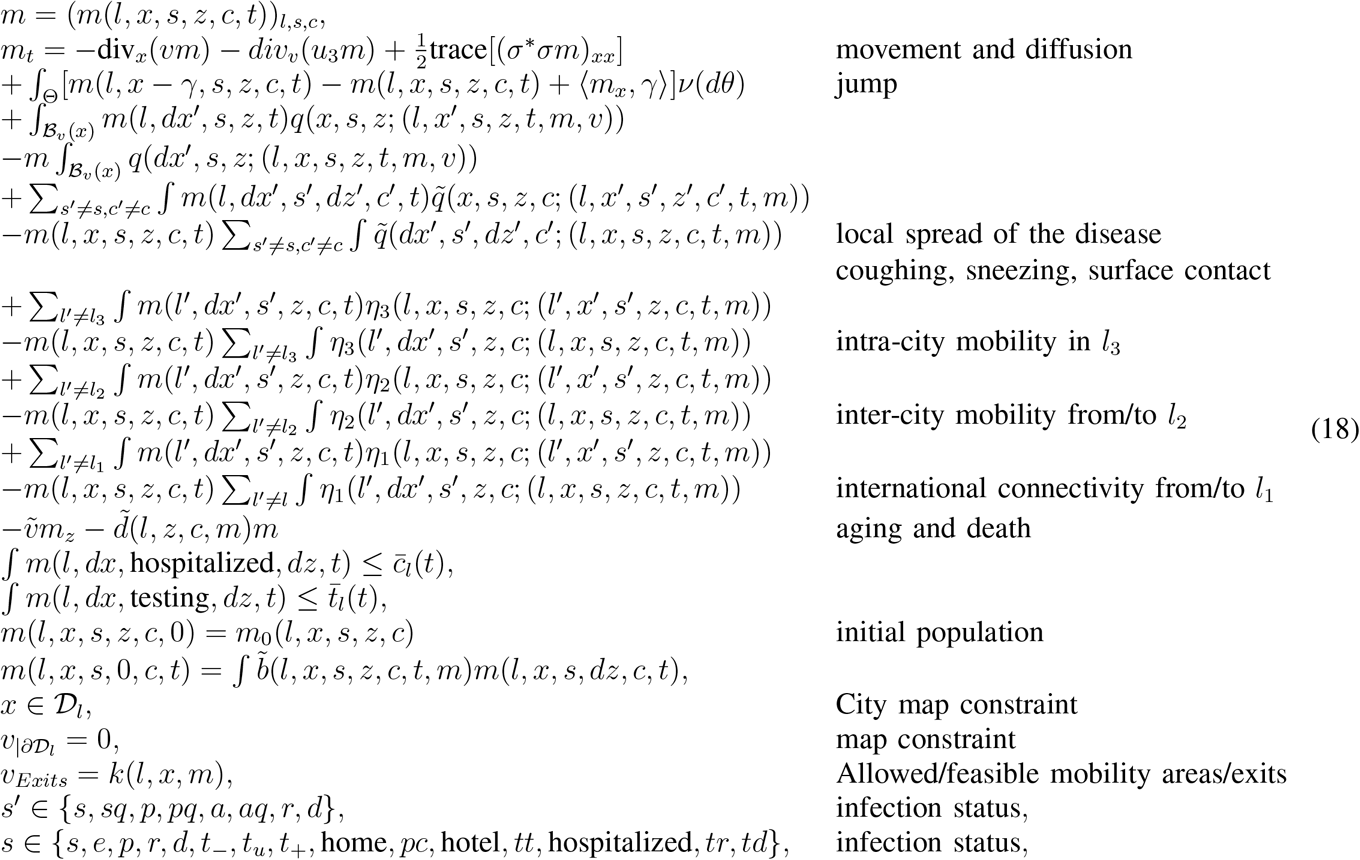

